# Differential Associations of Heat Metrics with Wellbeing across 13 Thai Provinces: The Role of Social Vulnerability in a Longitudinal Cohort of Older Adults

**DOI:** 10.64898/2026.01.06.26343526

**Authors:** Romnalin Keanjoom, Sittichai Choosumrong, Keiko Nakamura, Hoang Thuy Linh Nguyen, Nithra Kitreerawuttiwong, Sukrit Kirtsaeng, Juan Gonzalez Hijon, Jacques Wels

## Abstract

**Background:** Longitudinal evidence on the association between temperature fluctuations and psychological wellbeing remains limited, particularly in ageing populations of Southeast Asia. This study examines how heat exposure affects multiple wellbeing outcomes among older adults in Thailand.

**Methods:** We linked longitudinal data from 16,002 observations in the Health, Aging, and Retirement in Thailand (HART) study (2015-2023) with province-level meteorological data. Using a multilevel mixed-effects model, we decomposed heat indicators into within-province (monthly fluctuation) and between-province (long-term average) components to assess their independent associations with psychological distress, self-rated mental/physical health, and life quality. We additionally use negative controls to address causation.

**Results:** A 1°C within-province increase in apparent temperature was associated with higher psychological distress (β=0.10, 95%CI: 0.05, 0.14). Similar detrimental within-effects were observed for self-reported mental health (β=0.06, 95%CI: 0.03, 0.09). Between-province differences in average heat showed minimal or non-significant associations. The negative within-province effect of heat was significantly stronger for individuals with limitations in activities of daily living (ADL) (β=0.12, 95%CI: 0.08, 0.16 for physical health) and those in employment and with low socio-economic status. Results were consistent across all four heat metrics.

**Conclusions:** The wellbeing of Thai older adults is sensitive to within-province fluctuations in heat, but not to the long-term average heat of a province except for the physically frail and socioeconomically disadvantaged, who are vulnerable to both. The identified vulnerabilities highlight the need for targeted public health interventions, such as heat warnings and workplace protections, to build resilience in this ageing population.

## Background

Climate change represents a significant threat to population wellbeing, acting through both direct exposure to extreme weather and more gradual environmental degradation. Consequences for population wellbeing are acute – with events like hurricanes and wildfires linked to post-traumatic stress, depression, and anxiety [1] – and chronic, manifesting as ecological grief or anxiety [2]. These impacts are profoundly mediated by social and economic factors [3], with pre-existing health conditions [4], educational attainment [5], financial inequality [1] and deprivation [6] amplifying individual risk. Furthermore, climate crisis might disrupt health care provision for people with a mental health diagnosis [1].

Southeast Asia is a region of climate vulnerability[7], where a high frequency of extreme weather events converges with socio-economic disparities to exacerbate these wellbeing challenges.

Thailand presents a critical example within this region, where national-level vulnerability is compounded by distinct provincial climate patterns and population ageing [8,9]. The country has warmed by 1.3°C since 1970, increasing the severity of floods [10], droughts, and heat hazards [11] that will likely exacerbate in the coming decades [12]. The effect of climate change are strong amongst rural communities and has drastic effects on the agricultural sector [13] and framing communities [14,15]. But cities are not unexposed with frequent flooding in Bangkok [16] and increasing cases of seasonal influenza [17].

This creates a geographically diverse risk landscape: the North, South and West experience regular flooding [18,19], while the central and eastern regions face prolonged droughts [20]. The wellbeing impact of these changes is further shaped by social determinants. Previous single province studies have shown that factors such as age, income, marital status, and physical health are strong predictors of self-reported wellbeing and happiness [21]^,^ [22]^,^ [23].

Such a diverse climate landscape increases the risk of ecological fallacy [24]. Aggregating data at the national level obscures the substantial heterogeneity between Thai provinces in climate, economic profile, and population structure. A robust analysis must therefore account for both within-province and between-province differences.

Another issue is that previous studies mostly relied on ambient temperature. This overlooks the role of humidity and wind speed, which fundamentally shape human-perceived heat stress and its subsequent health impacts [25]. Addressing the associations between wellbeing and heat indicators that include humidity and wind speed characteristics – e.g. humidex [26], heat index [27–29] – is crucial to better frame how perceived heat and heat stress shape wellbeing outcomes^.^

The relationship between climate variability and mental health in Thailand remains inadequately researched as most research focus on high incomes countries [30,31] . A significant gap exists in the application of longitudinal study designs, which are necessary to move beyond association and establish the causal pathways through which socioeconomic and physical health factors mediate this relationship [32,33]. Against this backdrop, this study addresses the following research questions:

[R.Q.1] How are within-province fluctuations in heat indicators and between-province differences in average heat independently associated with wellbeing?

[R.Q.2] Does the association between short-term, within-province deviations in heat and wellbeing depend on the province’s long-term average climate (between-province heat)?

[R.Q.3] How consistent are the associations between heat exposure and wellbeing across different heat indicators?

[R.Q.4] To what extent do pre-existing physical health limitations and socioeconomic factors explain heterogeneity in vulnerability to the mental health associations with heat?

## Methods

### Protocol

We published a research protocol for this study on medRxiv: https://doi.org/10.1101/2025.08.12.25333382

### Data

**The Health, Ageing and Retirement in Thailand (HART)** – is biannual panel survey of adults aged 45+. It is sponsored by Fundamental Fund, Thailand Science Research and Innovation (TSRI) and is conducted by the National Institutes of Development Administration (NIDA). The sampling method typically involves stratified random sampling to ensure that different regions and socioeconomic groups are adequately represented. Data collection is carried out through Computer Assisted Personal Interviewing (CAPI). HART currently contains four waves, collected in 2015-17-20-22. It includes an original sample in 2015 and a refreshment sample in 2022, that is excluded in this study. Data were collected over 13 provinces (*Changwat*). Information on the month of data collection was provided by the NIDA. As can be seen in <u>supplementary file S1</u>, data were collected across all provinces and months in 2015 and 2017 whilst data collection in years 2020 and 2022 spanned over years 2020-21 and 2022-2023.

**Meteorological data** – We used daily meteorological data derived from observations collected at 131 weather stations across Thailand and provided by the Thai Meteorological Department as shown in **Figure 1**. Each station was georeferenced and assigned to its corresponding Thai province to generate province-level meteorological indicators. The dataset includes daily measurements of average temperature (°C), relative humidity (%), and wind speed (m/s). These variables were used to construct four heat indicators: actual Temperature, Humidex, Heat Index, and Apparent Temperature.

**Figure 1.**
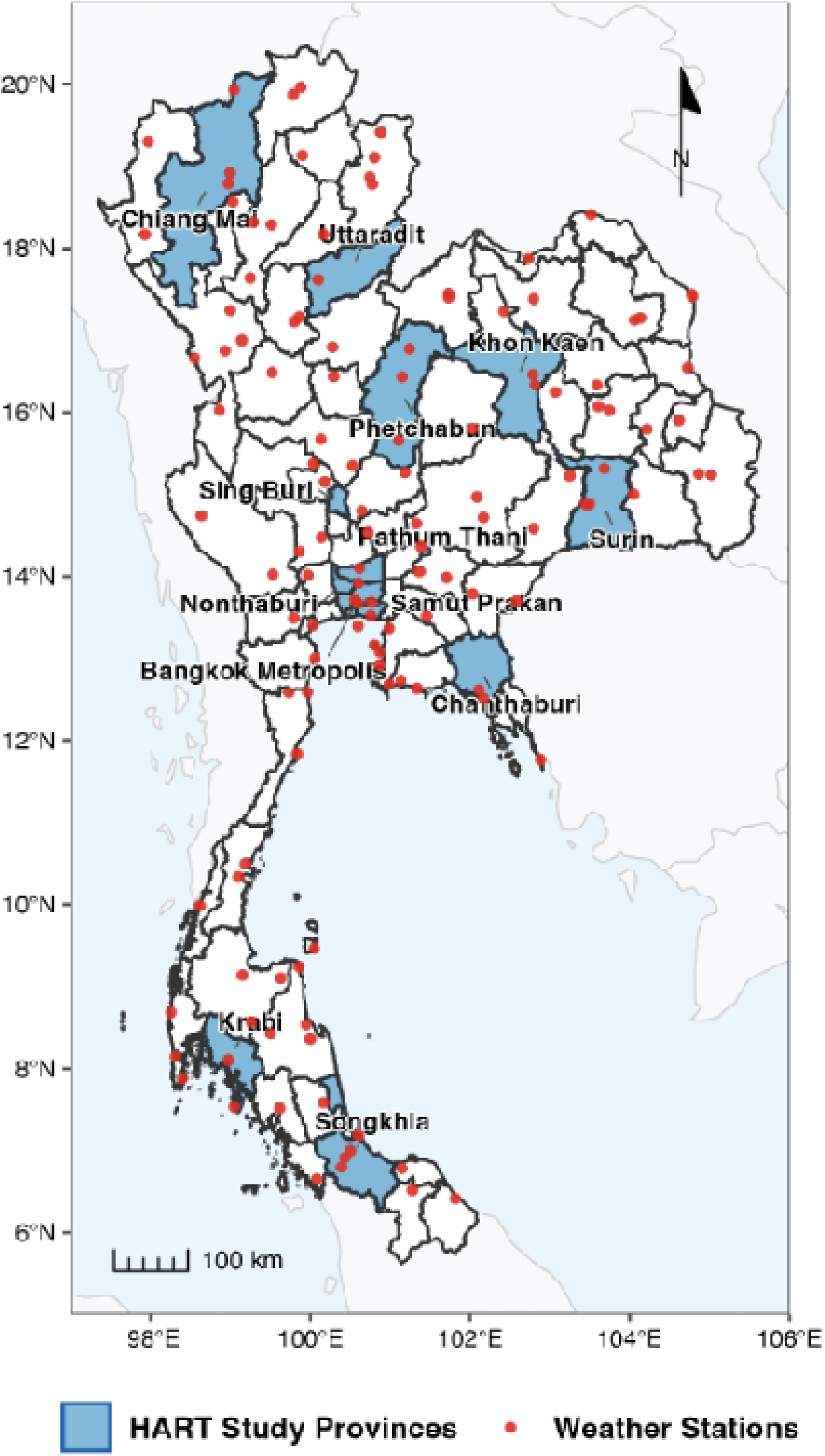
**Selected provinces and weather collection points**

We matched HART data with meteorological data based on the province, year and month of data collection. **Table 1** describes the longitudinal sample from HART, comprising a total 16,002 observations collected across four waves (2015: N=5,616; 2017: N=3,662; 2020-2021: N=4,207; 2022-2023: N=2,517). The health data from 13 provinces were matched with meteorological data from 12 weather stations. Due to the absence of weather stations in Nonthaburi and Sing Buri provinces, we implemented a geographical matching strategy: Nonthaburi was paired with adjacent Pathum Thani and Sing Buri with neighbouring Lop Buri, both pairs sharing similar climatic conditions and regional characteristics. This approach ensured comprehensive geographic coverage across Thai regions.

**Table 1.**
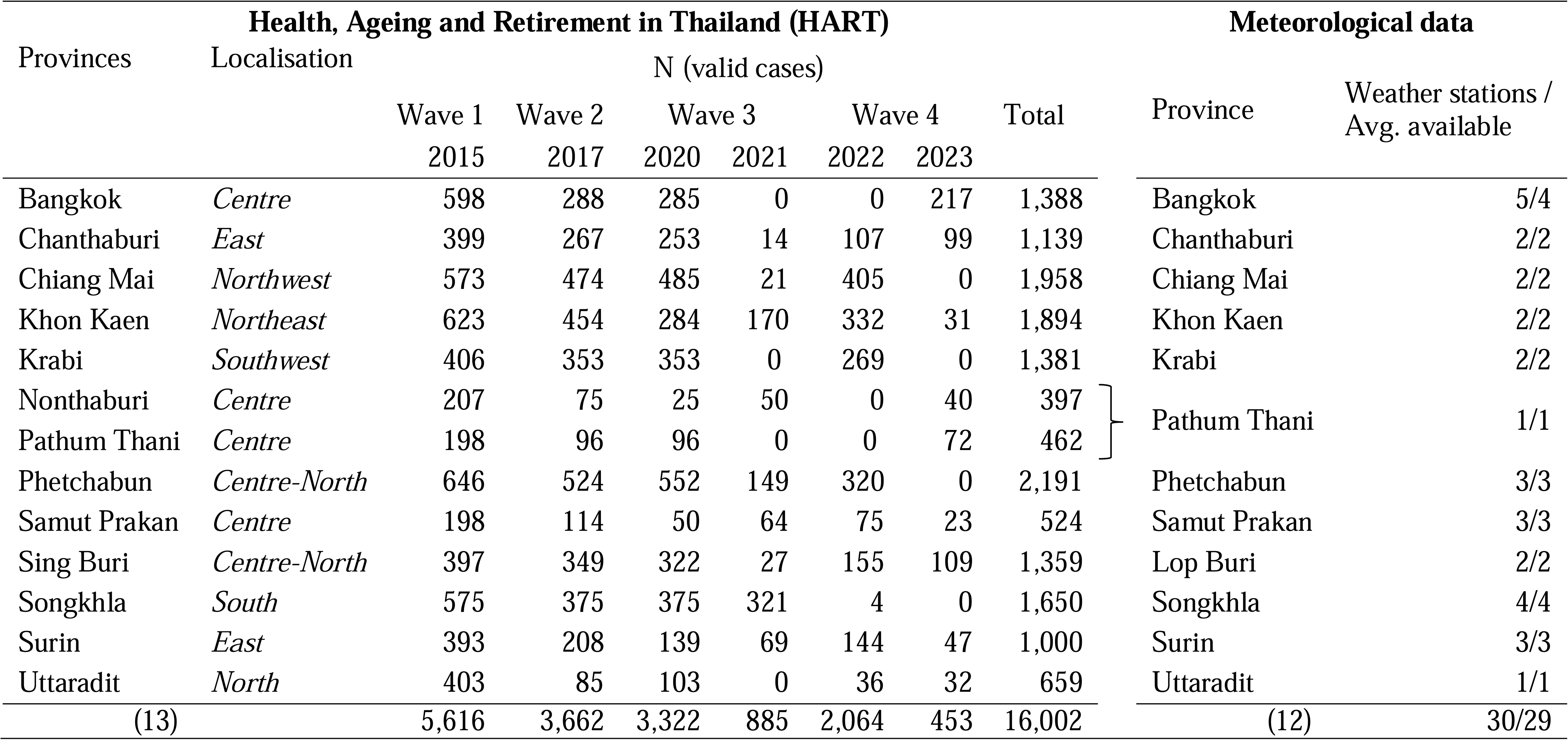
HART 13 provinces, sample size by year and available meteorological stations

### Wellbeing outcomes

#### HART includes four wellbeing-related variables

##### Psychological distress

HART includes several items related to psychological distress, specifically incorporating 10 questions similar to those used in the 12-item General Health Questionnaire (GHQ-12) [34], excluding items on feeling worthless and being able to face problems. Items include feeling bored, lack of concentration, feeling sad, feeling happy, feeling fearful, insomnia or difficulty sleeping, feeling satisfied, feeling lonely, feeling disappointed or unfulfilled and feeling bad, worthless or with no dignity. Response options are based on frequency (1: None, 2: 1-2, 3: 3-4, 4: 5-7 days/week). All items are summed (after reversing the happiness variable scale) to calculate a numeric total ranging from 1 (low psychological distress) to 31 (high psychological distress).

##### Self-reported mental health

Self-assessed mental health score coded from 1 (poor) to 10 (excellent) in all waves except 2017 where it is coded from 1 to 100. We equivalized the modalities to obtain a scale from 1 to 10 ranging from poor to excellent self-reported mental health. The variable is then rescaled from 1 (excellent) to 10 (poor).

##### Life quality

HART also includes a repeated item on satisfaction with life quality but the answer modalities changed over the waves. It was coded from 0 to 10 in 2015 and 2020, from 0 to 100 in 2017 and from 1 to 10 in 2022. The variable is equivalized from 1 (dissatisfied) to 10 (satisfied). The variable is then rescaled from 1 (satisfied) to 10 (dissatisfied).

##### Self-reported physical health

Self-assessed physical health score coded from 1 (poor) to 10 (excellent) in all waves except 2017 where it is coded from 1 to 100. We equivalized the modalities to obtain a scale from 1 to 10 ranging from poor to excellent self-reported mental health. The variable is then rescaled from 1 (excellent) to 10 (poor).

For interpretation consistency, all wellbeing metrics are scaled from high to poor wellbeing.

### Heat exposure

We compare four measures of heat commonly used to describe human-perceived heat stress. The actual temperature represents the ambient air temperature alone. Humidex combines air temperature and humidity to quantify perceived warmth, using dew point as a measure of atmospheric moisture [26]. The Heat Index also combines temperature and humidity but relies on relative humidity. It was designed to approximate the body’s reduced cooling efficiency under shaded, low-wind conditions [27–29]. The Apparent Temperature extends these measures by incorporating wind speed alongside temperature and humidity, providing a more comprehensive estimate [35–37]. Details on the calculation of these indicators are provided in <u>supplementary file S2</u> with descriptives statistics in files <u>S3 and S4</u>.

### Time varying cofounders

The study includes several cofounders that reflects social determinants of mental health wellbeing found in previous studies [23,38–43]. Activities of Daily Living (ADL) assessed self-help abilities in dressing, washing face and/or brushing teeth, bathing and/or washing hair and eating and is dichotomised, as in previous studies [44,45], distinguishing fully independent respondents (reference) from those who require any assistance. Education distinguished respondents with no formal education, with primary education or above (reference). Work status used a binary variable distinguishing respondents in employment or self-employment from non-employed respondents (reference) as complete information on non-work statuses available only in waves 2017 and 2022. Gender was coded as male or female (reference). Marital status was categorized as not married nor living together versus married or living together (reference). Household composition included four modalities: extended family (reference), living alone, living with other non-relatives, and nuclear family. Economic status satisfaction was measured on a scale from 1 to 10 (10 indicating poor satisfaction).

### Modelling

We estimated the association between apparent temperature and wellbeing outcomes using a multilevel [46] within–between mixed-effects model [47]. To avoid ecological bias – which arise when variation within units (e.g., provinces over time) is conflated with variation between units (e.g., average temperature differences across provinces) – we decomposed temperature into two components. The first component was the person-level apparent temperature at each observation, which captures *within-province* fluctuations. This allows us to estimate how deviations from typical local climatic conditions relate to changes in wellbeing outcomes. The second component was the province-level mean apparent temperature, which captures *between-province* differences in long-term climatic exposure (i.e., averages across provinces).

The model specified multiple random intercepts. A random intercept for each individual adjusts for the repeated measures and captures unobserved, time-invariant characteristics such as baseline health status, genetics, or stable lifestyle factors. A random intercept for provinces accounts for unmeasured local contextual characteristics, such as healthcare access, environmental pollution, or socioeconomic conditions [48]. Random intercepts for year and month capture temporal heterogeneity, while the month intercept adjusts for seasonal variations.

We replicated the model with a within-between heat metrics two-way interaction term, with a two-way interaction between vulnerability variables (ADL limitations, education economic and employment status) and within and between heat metrics and with a three-way interaction between heat metrics and vulnerability variables. Causal pathways were addressed using Directed Acyclic Graph as can be seen in <u>supplementary file S5</u>.

### Multiple imputations

Multiple imputation was performed to address attrition using the multilevel implementation of the Multivariate Imputation by Chained Equations framework. 20 imputed datasets were generated, and each imputation chain was iterated five times. Predictor matrices were constructed so that each variable was imputed using all relevant covariates and repeated measures, while the participant identifier served as the cluster-level variable and was not included as a predictor. The completed datasets were analysed separately and pooled according to Rubin’s rules [49].

### Software

All analyses were conducted in RStudio, Version 2024.12.1+563 using the: mice (v3.16.0)[50], lme4 (v1.1.34)[51] and broom.mixed (v0.2.9.4)[52] packages.

### Sensitivity analyses and negative control

We replicated the model using a one-month-lag, where temperature metrics are calculated the month prior the month of interview as a sensitivity check. Finally, to assess whether the observed associations are likely causal rather than attributable to unmeasured confounding, we also conducted a negative control analysis [53]. This involved substituting our primary outcome by a negative control outcome [54]. We choose perceived changes in children’s future prospects (coded from 0:positive change to 10:no change), a variable subject to similar confounders but with no plausible biological link to short-term temperature variation. The specificity of the association – present for health outcomes but absent for the negative control – strengthens the causal interpretation of our main models. To properly compare models, analyses were made on a sub-sample excluding respondents without reported children.

## Results

### Descriptive statistics

The selected provinces capture Thailand’s diverse climatic regions. Across these regions, average temperatures over the full period ranged from 23.9°C (Chiang Mai) to 29.8°C (Bangkok). However, the four heat metrics revealed distinct patterns: humidity-adjusted measures (humidex, heat index, apparent temperature) showed greater heat stress amplification in coastal provinces (e.g., Chanthaburi apparent temperature 5°C above measured temperature) compared to inland regions. This divergence demonstrates that temperature alone underestimating heat exposure in humid climates. See <u>Supplementary File 3</u> for complete data.

### Within-province and between-province differences in heat indicators

**Figure 2** shows the within and between effects for the four outcomes (life quality, physical health, mental health and psychological distress) and the four heat metrics (temperature, humidex, heat index and apparent temperature). What can be observed is that between effects tend to be negative indicating that a 1°C difference in temperature across provinces is associate with better wellbeing outcomes. However, all 95%CI overlap the null except for the model looking at psychological distress by apparent temperature [β=-0.20; 95%CI: -0.34, - 0.05]. By contrast the within effects calculating the change in temperature metrics within provinces independently of between province differences is positive in all models and significant in almost all models ranging from 0.02 [95%CI: -0.04, 0.09] in the model looking at psychological distress by temperature to 0.10 [95%CI: 0.05, 0.14] indicating that 1°C increase in heat metrics is associated with poorer wellbeing outcomes. No significant difference across health indicators is observed.

**Figure 2.**
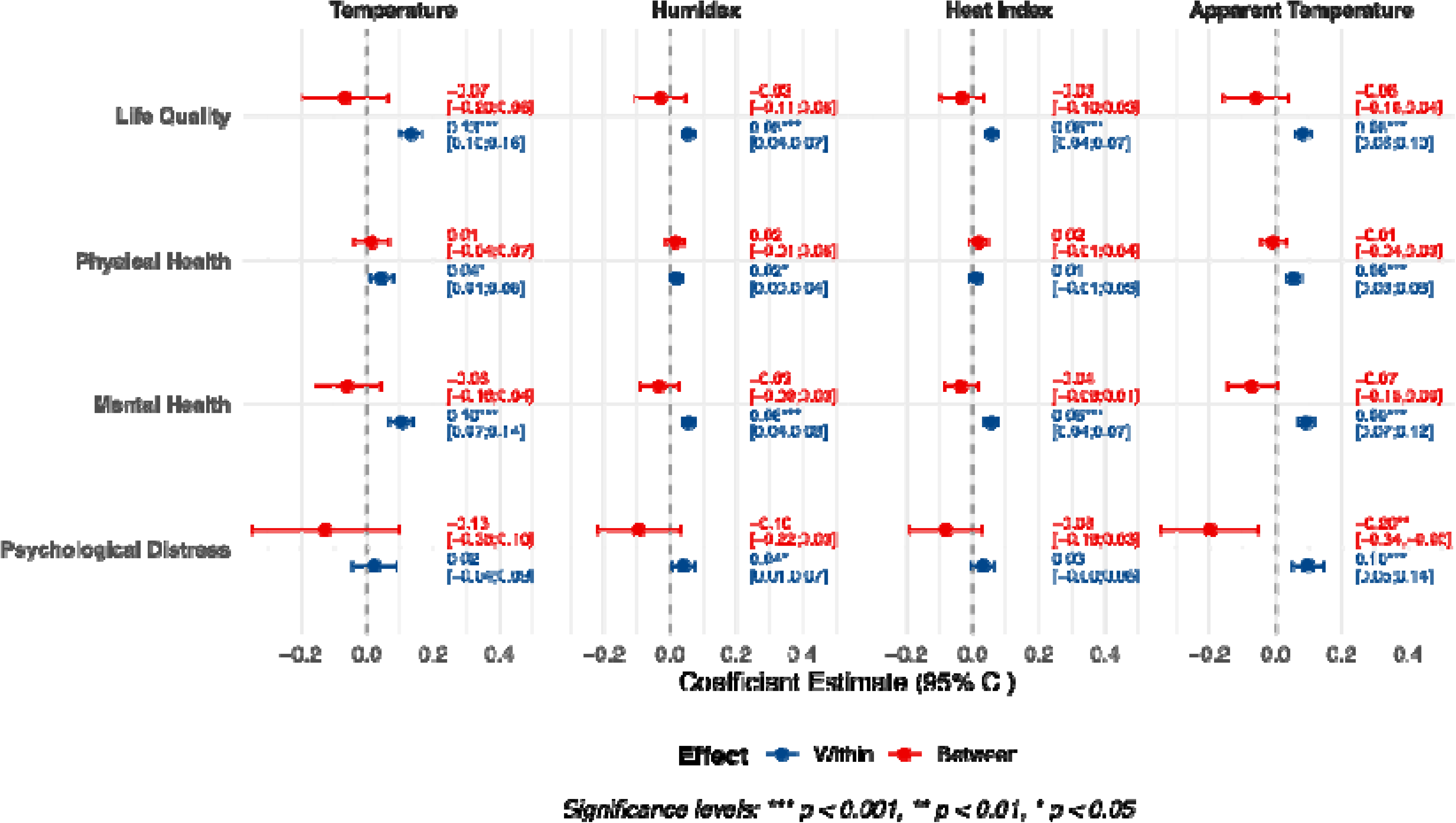
**Within and between effects of temperature indicators on wellbeing outcomes**

### Associations between within-province and between-province heat

**Figure 3** replicates the previous figure including an interaction term between the within and between effects. The interaction term allows us to estimate whether temperature increase in hot provinces is associated with poorer wellbeing scores, taking into consideration provincial heat resilience. The results indicate that both short-term temperature fluctuations and long-term climatic differences are associated with wellbeing. For instance, a 1°C increase above a province’s own long-term average monthly temperature is associated with a 0.38 increase in psychological distress, indicating a deterioration in wellbeing. Independently of these month-to-month variations, provinces that are on average 1 °C warmer tend to have psychological distress scores that are 0.26 points higher, suggesting that residents of structurally hotter provinces experience poorer wellbeing overall. The interaction term is small but statistically significant [β=-0.01; 95%CI: -0.02, -0.00]: the negative effect of warmer months is slightly weaker in provinces that are generally hotter. This pattern implies a modest degree of adaptation or resilience to chronic heat exposure in warm provinces.

**Figure 3.**
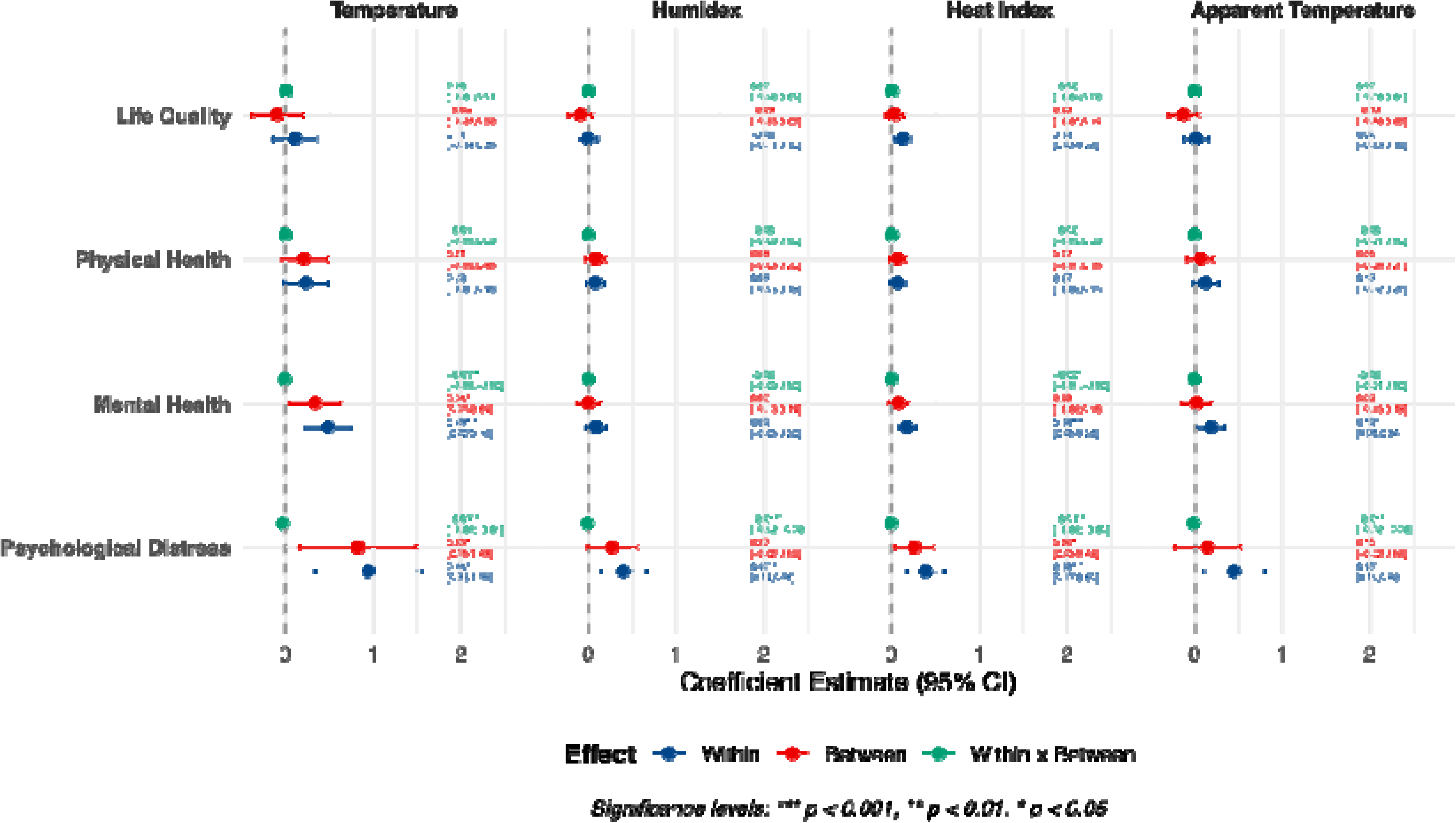
**Within, between and within*between interaction effects**

### Sub-group vulnerabilities

We look at sub-group vulnerabilities including an interaction term between the within and between effects and limitations in activities of daily living (ADL), employment status, education and financial wellbeing, respectively, as can be seen in **Figure 4**.

**Figure 4.**
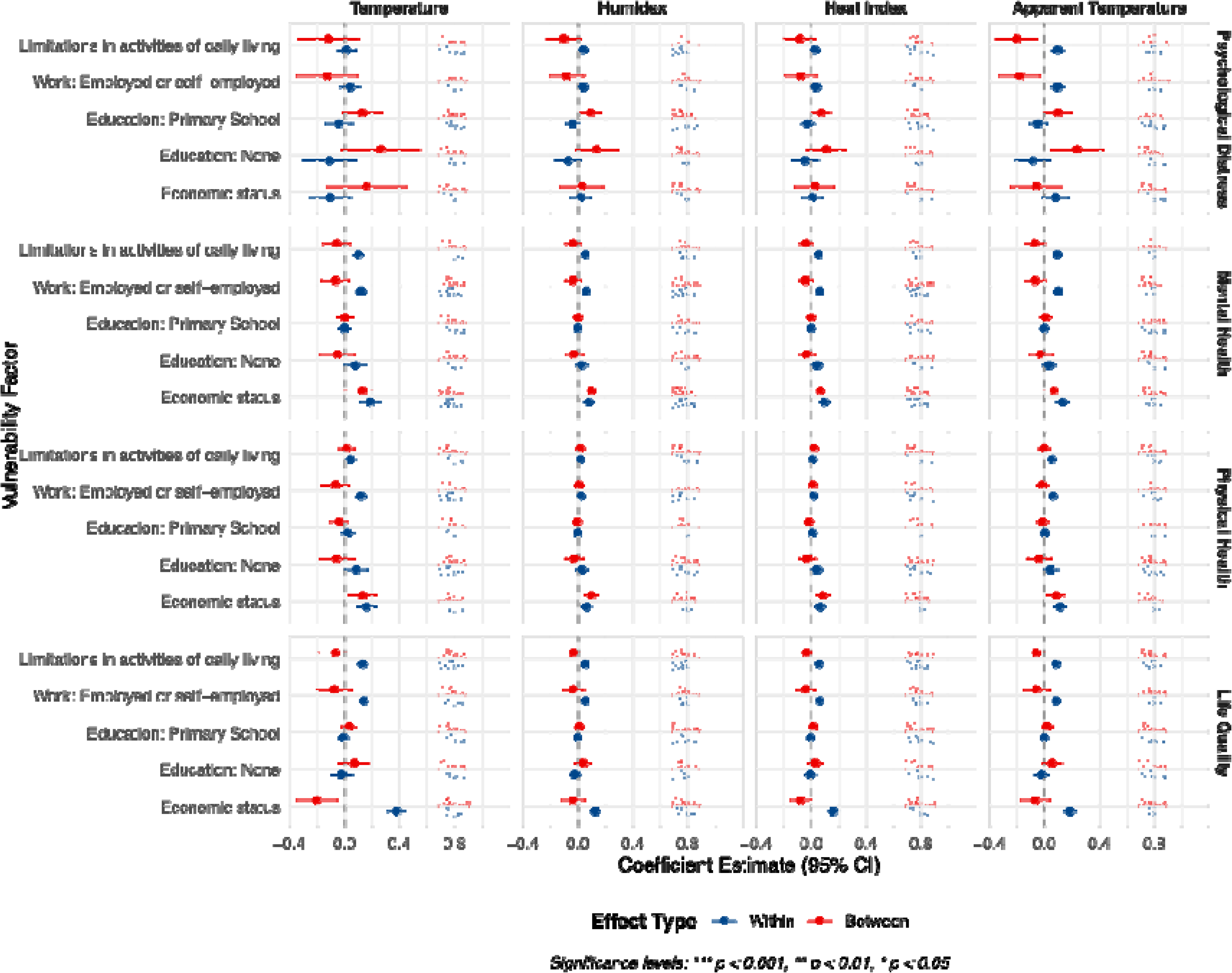
Within and between effects stratified by vulnerability factors

Looking at ADL, it can be observed that the within effects show a positive association with wellbeing outcome indicating that within provinces increase in heat indicators is associated with poorer wellbeing score. For instance, the coefficients are 0.10 [95%CI: 0.05, 0.14] in the model looking at psychological distress by apparent temperature and 0.06 [95%CI: 0.03, 0.08] when looking at physical health. All between effects overlap the null.

Similarly, reporting being in employment (versus not being in employment) is associated with poorer wellbeing within effects in all models with particularly strong associations observed for temperate/physical health [β=0.12; 95%CI:0.08,0.16] and for apparent temperature/psychological distress [β=0.10, 95%CI:0.05,0.14]. The between effects is not significant except in the apparent temperature / psychological distress model where a negative association is observed [β=-0.18, 95%CI:-0.33,-0.03].

No significant association is observed for the relationship between education and psychological distress and mental health. We observe positive coefficients between having no formal education and physical health when looking at temperatures [β=0.09; 95%CI:0.00,0.17]. Psychological distress shows a different pattern as reporting no formal education is associated with higher between effects coefficients, particularly for apparent temperatures [β=0.24, 95%CI: 0.04, 0.43].

Finally, no significant association is observed for self-reported economic status when looking at psychological distress but mental health shows both positive between and within effects. For temperature, the between effect is 0.13 [95%CI:0.00,0.26] and the within effect is 0.19 [95%CI:0.12,0.26]. The same is observed across other heat metrics. Similarly, the between and within effects are positively and significantly associated with poor physical health across all heat metrics. Life quality shows a different pattern with a positive within effects, indicating detrimental relationship with all heat metrics (e.g., β=0.38 [95%CI:0.31,0.45] for temperature), but a negative between effect temperatures [β=-0.20; 95%CI: -0.35, -0.5] and the heat index [β=-0.08; 95%CI:-0.15,-0.00].

The three-way interaction between within-, between-province heat, and vulnerability factors to test whether the association of vulnerabilities with wellbeing depends on this combined climatic context is shown in <u>Supplementary File S6</u>. For most vulnerability factors, their relationship with wellbeing is not substantially modified by the dual heat exposure. However, for individuals with physical health limitations (ADL) and those who are economically disadvantaged, increased temperatures within a chronically hot province was more detrimental to wellbeing than a temperatures increases in a cooler province.

### Sensitivity analyses and negative control

Because the monthly HART data were matched with monthly climate data, respondents who completed the survey at the beginning of a given month may have been affected by the previous month’s heat metrics. To account for this potential time lag, we replicated the analyses using a 1-month lag in which heat metrics were calculated for the month prior to HART data collection. All analyses were rerun and are presented in <u>Supplementary Files S7-S9</u>, with estimates identical to those in the non-lagged analyses.

The negative control analyses performed on a sub-sample is shown in <u>Supplementary file S10</u>. Whilst the associations between heat metrics and wellbeing indicators are similar to those observed previously, the negative outcome shows unsignificant associations for the between effects and unsignificant or negative associations for the within effects, strengthening the causal interpretation of our models.

## Discussion

This longitudinal study provides novel evidence on the relationship between heat exposure and wellbeing among older adults in Thailand, a country facing diverse and escalating climate threats. By disentangling within-province fluctuations from between-province differences in climate and employing multiple heat metrics, our analysis offers several key insights with broad relevance for climate and health policy.

First, we found that within-province increases in all heat metrics were consistently associated with poorer wellbeing outcomes (RQ1). This suggests that heat exposure, even within the normal range of a province’s climate, acts as a significant stressor. Conversely, the between-province effects indicated that residents of provinces with higher average temperatures did not consistently report poorer wellbeing. However, this adaptation appears to be incomplete and context dependent. Effect modification (RQ2) revealed that the detrimental associations of a within temperature increase with wellbeing was slightly attenuated in provinces with higher long-term average heat. This suggests that while populations in hotter provinces may develop some resilience, it does not fully buffer them from the negative effects of increased heat.

Third, our comparison of four heat indicators (RQ3) confirmed that ambient temperature alone is a sufficient metric to detect significant associations with wellbeing in this context. The more refined measures, such as Apparent Temperature and Humidex, did not yield substantially different results. This is a crucial methodological finding for public health surveillance, implying that widely available temperature data can be effectively used to monitor population-level wellbeing risks associated with heat in similar climatic zones.

However, this applies to wellbeing and could not be generalised to other health metrics, such as mortality [55].

Finally, our analysis of vulnerability (RQ4) underscores that the burden of heat-related wellbeing is not distributed equally. The stronger associations observed among individuals with limitations in Activities of Daily Living (ADL) highlight the heightened vulnerability of those in poorer physical health. Similarly, employed individuals showed stronger negative associations between heat and wellbeing than their non-employed counterparts. This may reflect the added physical strain of working in hot conditions[56]. Furthermore, both the within and between effects showed detrimental associations with wellbeing outcomes for both ADL and economic status, indicating a cumulative association of warm climate and increased temperatures.

Several limitations must be considered. Our measure of psychological distress, while robust, was not a complete clinical instrument. The reliance on self-reported economic status, though common, may introduce bias. The hierarchical design, while a strength, uses observational data; thus, we demonstrate robust associations but cannot definitively establish causation. Negative control analyses reinforce the assumption that these relationships may be causal. Furthermore, the characteristics of provinces (e.g., healthcare infrastructure, green space) may change over time, and our model did not dynamically account for such time variations. However, the model controlled from baseline characteristics and no health care reform was implemented during the period [57].

Key strengths include the use of a large, nationally representative longitudinal dataset, the application of a multilevel model that avoids ecological fallacy, and the concurrent testing of multiple heat metrics and wellbeing outcomes.

In conclusion, our findings demonstrate that heat is associated with wellbeing in Thailand’s older adults, with effects manifesting differently across temporal and geographical scales. The identification of employed individuals and those with physical health limitations as particularly vulnerable groups provides a clear target for public health interventions. Policymakers and health services must integrate mental wellbeing into heat adaptation and early-warning systems. Proactive measures, such as cooling shelters, workplace heat stress regulations, and wellness checks on vulnerable individuals, are critical.

## Data Availability

HART data are freely available at https://hart.nida.ac.th/about-hart/
Meteorological data are available upon request to the Thai Meteorological Department https://www.tmd.go.th/en/

https://hart.nida.ac.th/about-hart/

## Declarations Acknowledgments

The authors would like to thank the Thai Meteorological Department (TMD) for providing weather and climate data for Thailand, and the Health, Aging, and Retirement in Thailand (HART) project for providing access to health and retirement data.

## Funding

JW acknowledges fundings from the Belgian National Scientific Fund (FNRS), Incentive Grant for Scientific Research (MIS), Belgium (Grant number: 40021242) and Research Associate Grant (CQ), Belgium (Grant number: 40010931) as well as the United Kingdom Research and Innovation (UKRI) ‘UHealth’ (Grant number: UKRI1426). This work was partially supported by the Reinventing University Program 2024, The Ministry of Higher Education, Science, Research and Innovation (MHESI), Thailand (Grant number R2567A145), RK is the beneficiary. This work was supported by the Wallonia Brussels International (WBI) ASEM DUO 2025-2026, RK and JW are the beneficiaries. The funding sources had not role in this writing and submission of this study.

## Authors contributions

Conceptualization: JW, RK; Methodology: JW, RK; Software: JW; Validation: JGH; Formal analysis: JW; Investigation: JW, RH; Resources: RK, SC, JW; Data Curation: JW, RK, SC, SK; Writing - Original Draft: JW; Writing - Review & Editing: RK, KN, SC, HTLN, NK; Visualization: JW; Supervision: JW, RK; Project administration: RK, JW; Funding acquisition: RK, JW. Note: Romnalin Keanjoom: RK; Sittichai Choosumrong: SC; Keiko Nakamura: KN ; Hoang Thuy Linh Nguyen: HTLN ; Nithra Kitreerawuttiwong: NK; Sukrit Kirtsaeng: SK; Juan Gonzalez Hijon: JGH; Jacques Wels: JW

## Conflicts of interest

The authors report no conflict of interest

## Data availability

HART data are freely available at: https://hart.nida.ac.th/about-hart/

Meteorological data are available upon request to the Thai Meteorological Department: https://www.tmd.go.th/en/

## Use of AI

AI was not used in this study, except to assist in coding the map shown in Figure 1, where DeepSeek was used to help mapping neighbouring countries and longitudes.

## Notes

### Competing Interest Statement

The authors have declared no competing interest.

### Clinical Protocols

https://www.medrxiv.org/content/10.1101/2025.08.12.25333382v1

### Author Declarations

HART data are freely available at: https://hart.nida.ac.th/about-hart/ Meteorological data are available upon request to the Thai Meteorological Department: https://www.tmd.go.th/en/

