## Supplementary files for "Differential Associations of Heat Metrics with Wellbeing across 13 Thai Provinces: The Role of Social Vulnerability in a Longitudinal Cohort of Older Adults"

### S1. HART Data collection by province, year and month

| Province | Year | Month | | | | | | | | | | | | Total (year) | Total (all years) |
| --- | --- | --- | --- | --- | --- | --- | --- | --- | --- | --- | --- | --- | --- | --- | --- |
|  |  | 1 | 2 | 3 | 4 | 5 | 6 | 7 | 8 | 9 | 10 | 11 | 12 |  |  |
| Bangkok | 2015 | 0 | 65 | 121 | 0 | 407 | 5 | 0 | 0 | 0 | 0 | 0 | 0 | 598 | 1,388 |
|  | 2017 | 1 | 43 | 69 | 79 | 63 | 33 | 0 | 0 | 0 | 0 | 0 | 0 | 288 |  |
|  | 2020 | 0 | 0 | 0 | 0 | 0 | 0 | 0 | 1 | 2 | 137 | 82 | 63 | 285 |  |
|  | 2023 | 0 | 0 | 4 | 32 | 89 | 72 | 20 | 0 | 0 | 0 | 0 | 0 | 217 |  |
| Chanthaburi | 2015 | 0 | 0 | 399 | 0 | 0 | 0 | 0 | 0 | 0 | 0 | 0 | 0 | 399 | 1,139 |
|  | 2017 | 10 | 74 | 40 | 14 | 65 | 64 | 0 | 0 | 0 | 0 | 0 | 0 | 267 |  |
|  | 2020 | 0 | 0 | 0 | 0 | 0 | 0 | 0 | 0 | 83 | 18 | 77 | 75 | 253 |  |
|  | 2021 | 14 | 0 | 0 | 0 | 0 | 0 | 0 | 0 | 0 | 0 | 0 | 0 | 14 |  |
|  | 2022 | 0 | 0 | 0 | 0 | 0 | 0 | 0 | 0 | 0 | 0 | 10 | 97 | 107 |  |
|  | 2023 | 99 | 0 | 0 | 0 | 0 | 0 | 0 | 0 | 0 | 0 | 0 | 0 | 99 |  |
| Chiang mai | 2015 | 0 | 2 | 1 | 1 | 555 | 9 | 0 | 0 | 0 | 0 | 0 | 5 | 573 | 1,958 |
|  | 2017 | 5 | 124 | 145 | 107 | 38 | 55 | 0 | 0 | 0 | 0 | 0 | 0 | 474 |  |
|  | 2020 | 0 | 0 | 0 | 0 | 0 | 0 | 0 | 29 | 299 | 157 | 0 | 0 | 485 |  |
|  | 2021 | 0 | 21 | 0 | 0 | 0 | 0 | 0 | 0 | 0 | 0 | 0 | 0 | 21 |  |
|  | 2022 | 0 | 0 | 0 | 0 | 0 | 0 | 50 | 69 | 87 | 152 | 47 | 0 | 405 |  |
| Khon kaen | 2015 | 0 | 2 | 490 | 0 | 1 | 130 | 0 | 0 | 0 | 0 | 0 | 0 | 623 | 1,894 |
|  | 2017 | 4 | 120 | 73 | 10 | 35 | 212 | 0 | 0 | 0 | 0 | 0 | 0 | 454 |  |
|  | 2020 | 0 | 0 | 0 | 0 | 0 | 0 | 0 | 30 | 190 | 56 | 8 | 0 | 284 |  |
|  | 2021 | 0 | 30 | 0 | 140 | 0 | 0 | 0 | 0 | 0 | 0 | 0 | 0 | 170 |  |
|  | 2022 | 0 | 0 | 0 | 0 | 0 | 0 | 93 | 51 | 78 | 68 | 0 | 42 | 332 |  |
|  | 2023 | 0 | 31 | 0 | 0 | 0 | 0 | 0 | 0 | 0 | 0 | 0 | 0 | 31 |  |
| Krabi | 2015 | 1 | 0 | 0 | 0 | 53 | 262 | 90 | 0 | 0 | 0 | 0 | 0 | 406 | 1,381 |
|  | 2017 | 0 | 160 | 84 | 40 | 59 | 10 | 0 | 0 | 0 | 0 | 0 | 0 | 353 |  |
|  | 2020 | 0 | 0 | 0 | 0 | 0 | 0 | 0 | 0 | 0 | 54 | 276 | 23 | 353 |  |
|  | 2022 | 0 | 0 | 0 | 0 | 0 | 0 | 119 | 132 | 18 | 0 | 0 | 0 | 269 |  |
| Nonthaburi | 2015 | 0 | 0 | 0 | 0 | 52 | 136 | 19 | 0 | 0 | 0 | 0 | 0 | 207 | 397 |
|  | 2017 | 1 | 4 | 19 | 25 | 20 | 6 | 0 | 0 | 0 | 0 | 0 | 0 | 75 |  |
|  | 2020 | 0 | 0 | 0 | 0 | 0 | 0 | 0 | 0 | 0 | 4 | 21 | 0 | 25 |  |
|  | 2021 | 0 | 0 | 0 | 0 | 0 | 0 | 0 | 0 | 0 | 0 | 0 | 50 | 50 |  |
|  | 2023 | 0 | 0 | 0 | 0 | 0 | 0 | 36 | 4 | 0 | 0 | 0 | 0 | 40 |  |
| Pathum thani | 2015 | 0 | 0 | 0 | 1 | 195 | 2 | 0 | 0 | 0 | 0 | 0 | 0 | 198 | 462 |
|  | 2017 | 6 | 1 | 40 | 46 | 3 | 0 | 0 | 0 | 0 | 0 | 0 | 0 | 96 |  |
|  | 2020 | 0 | 0 | 0 | 0 | 0 | 0 | 0 | 0 | 0 | 18 | 78 | 0 | 96 |  |
|  | 2023 | 0 | 0 | 0 | 0 | 0 | 0 | 0 | 72 | 0 | 0 | 0 | 0 | 72 |  |
| Phetchabun | 2015 | 5 | 624 | 5 | 1 | 3 | 8 | 0 | 0 | 0 | 0 | 0 | 0 | 646 | 2,191 |
|  | 2017 | 0 | 9 | 0 | 161 | 325 | 29 | 0 | 0 | 0 | 0 | 0 | 0 | 524 |  |
|  | 2020 | 0 | 0 | 0 | 0 | 0 | 0 | 0 | 13 | 74 | 158 | 293 | 14 | 552 |  |
|  | 2022 | 0 | 0 | 0 | 0 | 0 | 0 | 0 | 0 | 0 | 0 | 95 | 54 | 149 |  |
|  | 2023 | 0 | 101 | 202 | 0 | 0 | 16 | 1 | 0 | 0 | 0 | 0 | 0 | 320 |  |
| Samut Prakan | 2015 | 0 | 1 | 86 | 111 | 0 | 0 | 0 | 0 | 0 | 0 | 0 | 0 | 198 | 524 |
|  | 2017 | 0 | 0 | 0 | 1 | 33 | 80 | 0 | 0 | 0 | 0 | 0 | 0 | 114 |  |
|  | 2020 | 0 | 0 | 0 | 0 | 0 | 0 | 0 | 0 | 1 | 15 | 33 | 1 | 50 |  |
|  | 2021 | 0 | 0 | 0 | 0 | 0 | 0 | 0 | 0 | 0 | 0 | 0 | 64 | 64 |  |
|  | 2022 | 0 | 0 | 0 | 0 | 0 | 0 | 0 | 47 | 1 | 27 | 0 | 0 | 75 |  |
|  | 2023 | 0 | 0 | 0 | 0 | 0 | 23 | 0 | 0 | 0 | 0 | 0 | 0 | 23 |  |
| Sing buri | 2015 | 2 | 393 | 0 | 0 | 0 | 2 | 0 | 0 | 0 | 0 | 0 | 0 | 397 | 1,359 |
|  | 2017 | 0 | 327 | 2 | 7 | 2 | 11 | 0 | 0 | 0 | 0 | 0 | 0 | 349 |  |
|  | 2020 | 0 | 0 | 0 | 0 | 0 | 0 | 0 | 0 | 235 | 46 | 41 | 0 | 322 |  |
|  | 2021 | 27 | 0 | 0 | 0 | 0 | 0 | 0 | 0 | 0 | 0 | 0 | 0 | 27 |  |
|  | 2022 | 0 | 0 | 0 | 0 | 0 | 0 | 0 | 0 | 0 | 27 | 0 | 128 | 155 |  |
|  | 2023 | 68 | 0 | 27 | 2 | 0 | 12 | 0 | 0 | 0 | 0 | 0 | 0 | 109 |  |
| Songkhla | 2015 | 1 | 0 | 0 | 4 | 25 | 508 | 37 | 0 | 0 | 0 | 0 | 0 | 575 | 1,650 |
|  | 2017 | 1 | 40 | 63 | 66 | 81 | 124 | 0 | 0 | 0 | 0 | 0 | 0 | 375 |  |
|  | 2020 | 0 | 0 | 0 | 0 | 0 | 0 | 0 | 49 | 166 | 140 | 20 | 0 | 375 |  |
|  | 2022 | 0 | 0 | 0 | 0 | 0 | 0 | 11 | 88 | 25 | 32 | 67 | 98 | 321 |  |
|  | 2023 | 2 | 2 | 0 | 0 | 0 | 0 | 0 | 0 | 0 | 0 | 0 | 0 | 4 |  |
| Surin | 2015 | 0 | 13 | 119 | 130 | 95 | 32 | 2 | 0 | 2 | 0 | 0 | 0 | 393 | 1,000 |
|  | 2017 | 6 | 11 | 5 | 24 | 46 | 116 | 0 | 0 | 0 | 0 | 0 | 0 | 208 |  |
|  | 2020 | 0 | 0 | 0 | 0 | 0 | 0 | 0 | 0 | 0 | 25 | 41 | 73 | 139 |  |
|  | 2021 | 1 | 0 | 7 | 61 | 0 | 0 | 0 | 0 | 0 | 0 | 0 | 0 | 69 |  |
|  | 2022 | 0 | 0 | 0 | 0 | 0 | 0 | 0 | 0 | 0 | 85 | 59 | 0 | 144 |  |
|  | 2023 | 0 | 8 | 39 | 0 | 0 | 0 | 0 | 0 | 0 | 0 | 0 | 0 | 47 |  |
| Uttaradit | 2015 | 0 | 0 | 0 | 235 | 156 | 12 | 0 | 0 | 0 | 0 | 0 | 0 | 403 | 659 |
|  | 2017 | 0 | 34 | 4 | 13 | 25 | 9 | 0 | 0 | 0 | 0 | 0 | 0 | 85 |  |
|  | 2020 | 0 | 0 | 0 | 0 | 0 | 0 | 0 | 14 | 61 | 28 | 0 | 0 | 103 |  |
|  | 2022 | 0 | 0 | 0 | 0 | 0 | 0 | 0 | 36 | 0 | 0 | 0 | 0 | 36 |  |
|  | 2023 | 0 | 32 | 0 | 0 | 0 | 0 | 0 | 0 | 0 | 0 | 0 | 0 | 32 |  |
| Total | | 254 | 2272 | 2044 | 1311 | 2426 | 1978 | 478 | 635 | 1322 | 1247 | 1248 | 787 | 16,002 | 16,002 |

### S2. Heat indicators

**Humidex** (short for "humidity index") is a metric developed by Canadian meteorologists to quantify the perceived temperature by humans, accounting for the combined effects of ambient temperature and humidity. Unlike relative humidity, the Humidex uses dew point temperature to provide an absolute measure of atmospheric moisture content, making it a more consistent indicator of mugginess throughout the day. The Humidex is calculated as follows ^26^:

Humidex = T + h, where h = (0.5555) × (e - 10.0) and e = 6.112 × 10^(7.5 × Td / (237.7 + Td))

Here, T is the air temperature (°C), Td is the dew point temperature (°C), and e is the vapour pressure in hectopascals (hPa). The dew point (Td) is itself derived from air temperature and relative humidity using the Magnus formula ^58,59^. The resulting Humidex value is a dimensionless number interpreted as an "equivalent feel-like" temperature in degrees Celsius.

The **Heat Index** (HI), also known as the "apparent temperature" in some contexts, is a measure developed by Steadman (1979) and adopted by the U.S. National Weather Service (NWS) that represents the human-perceived equivalent temperature based on the combined effects of ambient temperature and relative humidity. It is derived from a physiological model of human heat balance and approximates the body's inability to cool itself effectively through sweat evaporation when humidity is high. The Heat Index is calculated using a complex polynomial regression formula:

HI = c₁ + c₂T + c₃RH + c₄T*RH + c₅T² + c₆RH² + c₇T²*RH + c₈T*RH² + c₉T²*RH²

where T is the air temperature in degrees Fahrenheit (°F), RH is the relative humidity (%), and c₁–c₉ are empirically derived coefficients (c₁ = -42.379, c₂ = 2.04901523, c₃ = 10.14333127, c₄ = -0.22475541, c₅ = -0.00683783, c₆ = -0.05481717, c₇ = 0.00122874, c₈ = 0.00085282, c₉ = -0.00000199). The result is converted back to degrees Celsius for interpretation. The Heat Index is designed for use in shaded, light-wind conditions and provides critical thresholds for heat-related health risks ^28^.

The **Apparent temperature** (AT)**,** represents the perceived air temperature by humans, accounting for the combined effects of ambient temperature, relative humidity, and wind speed. While traditional heat indices rely solely on temperature and humidity, Steadman’s extended formula incorporates wind speed to provide a more realistic estimate of thermal comfort in warm conditions^35^. The apparent temperature (AT) in degrees Celsius (°C) is calculated as follow:

AT = T + (0.33 × e) - (0.70 × WS) - 4.00

where T is the air temperature (°C), WS is the wind speed (m/s), and e is the water vapor pressure in hPa. The vapor pressure e is derived from relative humidity (RH) (in %) and temperature using the modified Magnus formula:

e = (RH / 100) × 6.105 × exp((17.27 × T) / (237.7 + T))

The coefficients **17.27** and **237.7** in the Magnus formula are empirically derived and validated for temperatures between 0–50°C ^58,59^. This formulation reflects how AT increases with humidity (via vapor pressure) and decreases with wind speed, capturing the dual role of moisture and ventilation in perceived heat. This formulation allows apparent temperature to increase with humidity (via vapor pressure) and decrease with higher wind speeds, reflecting the dual influence of moisture and ventilation on perceived heat. Although daily rainfall does not enter the equation directly, it often affects apparent temperature indirectly by increasing near-surface humidity and altering wind patterns following precipitation events. For analytical purposes, rainfall may be included as a contextual variable rather than as an input in the heat index formula itself.

### S3. Average temperatures over the selected periods (2015, 2017, 2020-2023)

| **Province** | **Temperature** | **Humidex** | **Heat Index** | **Apparent**  **Temperature** |
| --- | --- | --- | --- | --- |
| Amnat Charoen | 27.14 | 37.04 | 30.95 | 30.57 |
| **Bangkok** | **29.84** | **41.07** | **35.58** | **33.52** |
| Bueng Kan | 27.14 | 37.38 | 31.51 | 30.73 |
| Buriram | 27.90 | 38.26 | 32.27 | 32.04 |
| Chachoengsao | 28.54 | 40.17 | 33.99 | 33.80 |
| Chai Nat | 29.11 | 40.08 | 34.34 | 33.73 |
| Chaiyaphum | 28.39 | 37.67 | 32.16 | 31.44 |
| **Chanthaburi** | **28.31** | **39.95** | **33.33** | **33.33** |
| **Chiang Mai** | **23.92** | **30.65** | **25.06** | **25.42** |
| Chiang Rai | 26.08 | 35.14 | 28.91 | 29.78 |
| Chon Buri | 29.09 | 40.53 | 34.48 | 33.12 |
| Chumphon | 28.06 | 39.68 | 32.63 | 33.21 |
| Kalasin | 27.69 | 38.38 | 32.54 | 31.68 |
| Kamphaeng Phet | 28.82 | 39.61 | 33.69 | 33.66 |
| Kanchanaburi | 28.63 | 39.00 | 32.89 | 33.30 |
| **Khon Kaen** | **27.92** | **37.79** | **32.04** | **31.64** |
| **Krabi** | **28.10** | **39.95** | **32.75** | **32.18** |
| Lampang | 27.87 | 37.75 | 31.62 | 32.50 |
| Lamphun | 28.06 | 37.73 | 31.83 | 32.39 |
| Loei | 27.11 | 37.12 | 30.96 | 31.35 |
| **Lopburi** | **29.38** | **40.33** | **34.89** | **33.72** |
| Mae Hong Son | 27.42 | 37.30 | 30.96 | 32.38 |
| Maha Sarakham | 28.31 | 38.54 | 32.86 | 32.18 |
| Mukdahan | 28.05 | 38.09 | 32.54 | 32.18 |
| Nakhon Nayok | 21.16 | 27.99 | 20.24 | 22.91 |
| Nakhon Pathom | 28.71 | 40.31 | 34.36 | 33.76 |
| Nakhon Phanom | 27.03 | 36.29 | 30.81 | 30.85 |
| Nakhon Ratchasima | 27.87 | 37.65 | 31.70 | 31.00 |
| Nakhon Sawan | 29.35 | 40.11 | 34.59 | 33.97 |
| Nakhon Si Thammarat | 28.30 | 40.77 | 33.74 | 34.03 |
| Nan | 27.03 | 36.99 | 30.62 | 31.91 |
| Narathiwat | 27.86 | 38.95 | 31.77 | 32.14 |
| Nong Bua Lamphu | 27.66 | 37.58 | 31.83 | 31.53 |
| Nong Khai | 27.98 | 37.73 | 32.17 | 31.58 |
| **Pathum Thani** | **29.96** | **41.86** | **36.54** | **34.44** |
| Pattani | 28.44 | 40.33 | 33.45 | 33.87 |
| Phang Nga | 28.01 | 39.90 | 32.69 | 32.90 |
| Phatthalung | 28.45 | 40.96 | 33.99 | 33.66 |
| Phayao | 26.79 | 36.15 | 29.78 | 30.59 |
| **Phetchabun** | **28.81** | **39.13** | **33.42** | **33.66** |
| Phetchaburi | 28.63 | 39.81 | 33.56 | 32.58 |
| Phichit | 29.08 | 40.38 | 34.50 | 33.30 |
| Phitsanulok | 28.72 | 39.43 | 33.51 | 32.87 |
| Phra Nakhon Si Ayutthaya | 29.10 | 40.36 | 34.56 | 32.73 |
| Phrae | 28.22 | 38.47 | 32.52 | 32.64 |
| Phuket | 28.80 | 40.30 | 33.71 | 33.34 |
| Prachin Buri | 29.25 | 40.72 | 35.08 | 34.78 |
| Prachuap Khiri Khan | 28.51 | 39.41 | 33.09 | 32.69 |
| Ranong | 28.06 | 39.10 | 32.11 | 32.47 |
| Ratchaburi | 28.81 | 40.38 | 34.35 | 33.17 |
| Rayong | 28.85 | 40.43 | 34.28 | 33.35 |
| Roi Et | 27.93 | 37.95 | 32.25 | 31.07 |
| Sa Kaeo | 29.03 | 40.56 | 34.78 | 34.17 |
| Sakon Nakhon | 27.28 | 37.17 | 31.46 | 31.23 |
| **Samut Prakan** | **29.12** | **40.31** | **34.42** | **31.19** |
| Samut Songkhram | 28.57 | 39.63 | 33.40 | 31.15 |
| Satun | 28.62 | 40.73 | 33.97 | 34.34 |
| Si Sa Ket | 28.31 | 38.69 | 32.99 | 31.90 |
| **Songkhla** | **28.29** | **40.11** | **33.13** | **32.73** |
| Sukhothai | 28.91 | 40.39 | 34.38 | 34.26 |
| Suphan Buri | 29.23 | 40.38 | 34.62 | 34.15 |
| Surat Thani | 28.17 | 40.26 | 33.15 | 33.48 |
| **Surin** | **27.99** | **37.98** | **32.04** | **32.15** |
| Tak | 26.69 | 35.66 | 29.21 | 30.02 |
| Trang | 28.57 | 40.49 | 33.76 | 33.27 |
| Trat | 28.19 | 39.90 | 33.02 | 32.52 |
| Ubon Ratchathani | 28.35 | 38.68 | 32.86 | 32.07 |
| Udon Thani | 27.99 | 37.78 | 32.28 | 31.42 |
| Uthai Thani | 29.46 | 40.61 | 35.12 | 33.91 |
| **Uttaradit** | **29.41** | **39.95** | **34.49** | **34.64** |
| Yala | 28.37 | 40.27 | 33.35 | 33.49 |
| Yasothon | 27.69 | 38.04 | 32.12 | 31.22 |

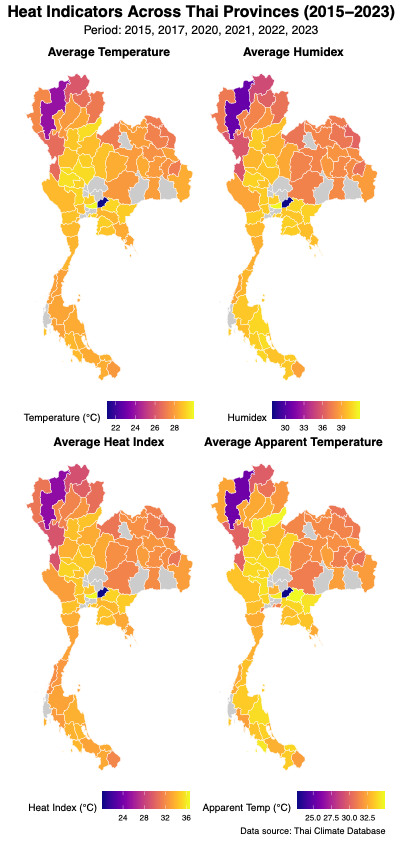

### S4. Mean monthly temperature indicators by province, year and month

| **Province** | **year** | **month** | **Temperature** | **Humidex** | **Heat index** | **Apparent**  **temperature** |
| --- | --- | --- | --- | --- | --- | --- |
| Bangkok | 2015 | 1 | 26.65 | 33.76 | 28.77 | 28.21 |
| Bangkok | 2015 | 2 | 28.75 | 37.77 | 32.04 | 30.83 |
| Bangkok | 2015 | 3 | 30.37 | 42.44 | 36.92 | 33.70 |
| Bangkok | 2015 | 4 | 31.04 | 42.71 | 37.54 | 34.65 |
| Bangkok | 2015 | 5 | 31.96 | 44.68 | 40.17 | 36.07 |
| Bangkok | 2015 | 6 | 30.96 | 42.69 | 37.35 | 34.61 |
| Bangkok | 2015 | 7 | 30.66 | 42.11 | 36.52 | 33.94 |
| Bangkok | 2015 | 8 | 30.62 | 42.02 | 36.45 | 34.01 |
| Bangkok | 2015 | 9 | 29.75 | 42.11 | 36.06 | 34.23 |
| Bangkok | 2015 | 10 | 29.26 | 41.49 | 35.13 | 34.32 |
| Bangkok | 2015 | 11 | 30.12 | 41.63 | 35.96 | 34.41 |
| Bangkok | 2015 | 12 | 29.03 | 38.71 | 32.96 | 32.19 |
| Bangkok | 2017 | 1 | 28.31 | 37.58 | 31.25 | 31.15 |
| Bangkok | 2017 | 2 | 29.08 | 37.63 | 32.17 | 31.53 |
| Bangkok | 2017 | 3 | 30.90 | 43.15 | 38.00 | 35.00 |
| Bangkok | 2017 | 4 | 31.53 | 44.26 | 39.48 | 36.09 |
| Bangkok | 2017 | 5 | 30.49 | 44.42 | 39.31 | 36.26 |
| Bangkok | 2017 | 6 | 30.44 | 42.99 | 37.56 | 35.13 |
| Bangkok | 2017 | 7 | 29.69 | 41.81 | 35.82 | 33.99 |
| Bangkok | 2017 | 8 | 30.16 | 42.68 | 37.01 | 34.88 |
| Bangkok | 2017 | 9 | 30.11 | 42.90 | 37.28 | 35.22 |
| Bangkok | 2017 | 10 | 29.06 | 41.65 | 35.31 | 34.32 |
| Bangkok | 2017 | 11 | 28.74 | 38.99 | 32.94 | 31.96 |
| Bangkok | 2017 | 12 | 26.89 | 34.67 | 29.56 | 28.49 |
| Bangkok | 2020 | 1 | 29.68 | 40.12 | 34.31 | 33.08 |
| Bangkok | 2020 | 2 | 29.81 | 39.67 | 33.98 | 32.57 |
| Bangkok | 2020 | 3 | 31.26 | 43.78 | 38.80 | 34.92 |
| Bangkok | 2020 | 4 | 31.80 | 44.87 | 40.38 | 35.94 |
| Bangkok | 2020 | 5 | 32.54 | 46.11 | 42.19 | 37.21 |
| Bangkok | 2020 | 6 | 30.69 | 43.35 | 38.04 | 35.24 |
| Bangkok | 2020 | 7 | 30.65 | 43.05 | 37.68 | 35.19 |
| Bangkok | 2020 | 8 | 30.19 | 42.31 | 36.58 | 34.26 |
| Bangkok | 2020 | 9 | 29.92 | 42.60 | 36.79 | 34.78 |
| Bangkok | 2020 | 10 | 27.96 | 39.21 | 31.97 | 32.11 |
| Bangkok | 2020 | 11 | 28.97 | 38.33 | 32.48 | 31.73 |
| Bangkok | 2020 | 12 | 27.66 | 35.20 | 29.96 | 29.59 |
| Bangkok | 2021 | 1 | 26.47 | 32.66 | 28.31 | 27.52 |
| Bangkok | 2021 | 2 | 28.84 | 37.46 | 31.71 | 31.15 |
| Bangkok | 2021 | 3 | 30.74 | 43.00 | 37.74 | 34.64 |
| Bangkok | 2021 | 4 | 30.59 | 43.60 | 38.29 | 35.46 |
| Bangkok | 2021 | 5 | 31.26 | 43.79 | 38.79 | 35.67 |
| Bangkok | 2021 | 6 | 31.16 | 43.06 | 37.87 | 34.82 |
| Bangkok | 2021 | 7 | 30.15 | 42.61 | 36.90 | 34.40 |
| Bangkok | 2021 | 8 | 30.22 | 43.09 | 37.49 | 34.70 |
| Bangkok | 2021 | 9 | 29.11 | 41.64 | 35.16 | 33.96 |
| Bangkok | 2021 | 10 | 29.26 | 41.51 | 35.26 | 33.65 |
| Bangkok | 2021 | 11 | 29.05 | 39.55 | 33.51 | 32.39 |
| Bangkok | 2021 | 12 | 27.51 | 34.37 | 29.11 | 28.87 |
| Bangkok | 2022 | 1 | 28.79 | 37.36 | 31.50 | 31.16 |
| Bangkok | 2022 | 2 | 28.88 | 39.01 | 32.98 | 32.00 |
| Bangkok | 2022 | 3 | 30.90 | 43.74 | 38.65 | 35.17 |
| Bangkok | 2022 | 4 | 31.16 | 42.58 | 37.88 | 34.61 |
| Bangkok | 2022 | 5 | 30.25 | 42.79 | 37.44 | 34.35 |
| Bangkok | 2022 | 6 | 30.77 | 43.34 | 38.08 | 35.16 |
| Bangkok | 2022 | 7 | 30.18 | 42.85 | 37.25 | 34.52 |
| Bangkok | 2022 | 8 | 29.57 | 42.20 | 36.16 | 34.10 |
| Bangkok | 2022 | 9 | 28.79 | 41.48 | 34.72 | 33.68 |
| Bangkok | 2022 | 10 | 28.83 | 39.75 | 33.51 | 32.73 |
| Bangkok | 2022 | 11 | 29.35 | 39.85 | 33.98 | 33.19 |
| Bangkok | 2022 | 12 | 27.40 | 34.82 | 29.72 | 29.08 |
| Bangkok | 2023 | 1 | 27.42 | 34.31 | 29.15 | 28.67 |
| Bangkok | 2023 | 2 | 29.09 | 38.62 | 32.96 | 31.86 |
| Bangkok | 2023 | 3 | 30.51 | 41.55 | 36.34 | 33.71 |
| Bangkok | 2023 | 4 | 32.43 | 45.60 | 41.44 | 36.96 |
| Bangkok | 2023 | 5 | 32.81 | 45.56 | 41.40 | 36.92 |
| Bangkok | 2023 | 6 | 31.68 | 43.79 | 38.89 | 35.21 |
| Bangkok | 2023 | 7 | 31.03 | 42.99 | 37.76 | 34.20 |
| Bangkok | 2023 | 8 | 30.92 | 43.31 | 38.12 | 34.66 |
| Bangkok | 2023 | 9 | 29.99 | 42.49 | 36.72 | 34.34 |
| Bangkok | 2023 | 10 | 29.74 | 42.23 | 36.30 | 34.54 |
| Bangkok | 2023 | 11 | 29.19 | 39.11 | 33.49 | 32.26 |
| Bangkok | 2023 | 12 | 29.05 | 37.87 | 32.61 | 31.40 |
| Chanthaburi | 2015 | 1 | 26.18 | 33.49 | 28.11 | 28.31 |
| Chanthaburi | 2015 | 2 | 26.89 | 36.14 | 29.36 | 30.58 |
| Chanthaburi | 2015 | 3 | 28.21 | 39.75 | 32.73 | 33.44 |
| Chanthaburi | 2015 | 4 | 29.37 | 41.59 | 35.47 | 35.01 |
| Chanthaburi | 2015 | 5 | 29.70 | 42.99 | 37.18 | 35.94 |
| Chanthaburi | 2015 | 6 | 28.88 | 41.84 | 35.15 | 35.01 |
| Chanthaburi | 2015 | 7 | 28.79 | 41.53 | 34.86 | 34.43 |
| Chanthaburi | 2015 | 8 | 28.37 | 41.14 | 34.10 | 34.22 |
| Chanthaburi | 2015 | 9 | 28.08 | 40.99 | 33.57 | 34.30 |
| Chanthaburi | 2015 | 10 | 28.31 | 40.63 | 33.45 | 33.97 |
| Chanthaburi | 2015 | 11 | 28.88 | 40.52 | 34.03 | 33.96 |
| Chanthaburi | 2015 | 12 | 28.09 | 38.21 | 31.94 | 32.20 |
| Chanthaburi | 2017 | 1 | 28.02 | 37.47 | 31.06 | 31.44 |
| Chanthaburi | 2017 | 2 | 27.88 | 37.16 | 31.04 | 31.26 |
| Chanthaburi | 2017 | 3 | 28.69 | 40.52 | 33.87 | 34.15 |
| Chanthaburi | 2017 | 4 | 29.33 | 42.58 | 36.45 | 35.73 |
| Chanthaburi | 2017 | 5 | 29.02 | 42.71 | 36.23 | 35.89 |
| Chanthaburi | 2017 | 6 | 28.42 | 41.48 | 34.37 | 34.68 |
| Chanthaburi | 2017 | 7 | 27.86 | 40.56 | 32.92 | 33.76 |
| Chanthaburi | 2017 | 8 | 28.86 | 42.30 | 35.67 | 35.16 |
| Chanthaburi | 2017 | 9 | 28.79 | 42.26 | 35.59 | 35.38 |
| Chanthaburi | 2017 | 10 | 28.76 | 41.78 | 35.15 | 34.81 |
| Chanthaburi | 2017 | 11 | 27.58 | 38.14 | 31.35 | 31.36 |
| Chanthaburi | 2017 | 12 | 26.64 | 34.98 | 29.32 | 28.69 |
| Chanthaburi | 2020 | 1 | 28.54 | 39.11 | 32.61 | 33.16 |
| Chanthaburi | 2020 | 2 | 28.44 | 38.37 | 31.89 | 32.29 |
| Chanthaburi | 2020 | 3 | 29.43 | 42.00 | 35.90 | 35.10 |
| Chanthaburi | 2020 | 4 | 29.64 | 42.50 | 36.54 | 35.78 |
| Chanthaburi | 2020 | 5 | 29.92 | 43.79 | 38.24 | 36.83 |
| Chanthaburi | 2020 | 6 | 28.36 | 41.31 | 34.16 | 34.48 |
| Chanthaburi | 2020 | 7 | 28.67 | 41.63 | 34.86 | 34.78 |
| Chanthaburi | 2020 | 8 | 28.17 | 40.79 | 33.48 | 33.85 |
| Chanthaburi | 2020 | 9 | 28.02 | 40.74 | 33.28 | 34.12 |
| Chanthaburi | 2020 | 10 | 27.07 | 38.55 | 30.47 | 32.31 |
| Chanthaburi | 2020 | 11 | 28.01 | 38.27 | 31.71 | 31.56 |
| Chanthaburi | 2020 | 12 | 27.12 | 35.87 | 29.85 | 29.70 |
| Chanthaburi | 2021 | 1 | 26.07 | 33.50 | 28.09 | 27.70 |
| Chanthaburi | 2021 | 2 | 27.22 | 36.98 | 29.89 | 31.19 |
| Chanthaburi | 2021 | 3 | 28.75 | 40.98 | 34.27 | 34.41 |
| Chanthaburi | 2021 | 4 | 28.88 | 41.86 | 35.24 | 35.11 |
| Chanthaburi | 2021 | 5 | 29.30 | 42.35 | 36.15 | 35.44 |
| Chanthaburi | 2021 | 6 | 28.81 | 41.62 | 34.97 | 34.52 |
| Chanthaburi | 2021 | 7 | 28.35 | 41.09 | 33.99 | 34.04 |
| Chanthaburi | 2021 | 8 | 28.44 | 41.29 | 34.24 | 34.35 |
| Chanthaburi | 2021 | 9 | 27.47 | 39.99 | 31.95 | 33.44 |
| Chanthaburi | 2021 | 10 | 27.89 | 40.37 | 32.86 | 33.66 |
| Chanthaburi | 2021 | 11 | 28.05 | 39.12 | 32.35 | 32.12 |
| Chanthaburi | 2021 | 12 | 26.84 | 34.69 | 28.78 | 28.28 |
| Chanthaburi | 2022 | 1 | 27.39 | 36.45 | 29.79 | 30.75 |
| Chanthaburi | 2022 | 2 | 27.78 | 38.28 | 31.25 | 31.96 |
| Chanthaburi | 2022 | 3 | 28.67 | 41.17 | 34.45 | 34.41 |
| Chanthaburi | 2022 | 4 | 28.80 | 40.24 | 34.04 | 33.51 |
| Chanthaburi | 2022 | 5 | 28.53 | 41.19 | 34.42 | 34.03 |
| Chanthaburi | 2022 | 6 | 28.95 | 41.80 | 35.27 | 34.93 |
| Chanthaburi | 2022 | 7 | 28.13 | 41.04 | 33.68 | 33.93 |
| Chanthaburi | 2022 | 8 | 27.89 | 40.53 | 32.91 | 33.55 |
| Chanthaburi | 2022 | 9 | 27.42 | 40.01 | 31.89 | 33.48 |
| Chanthaburi | 2022 | 10 | 28.22 | 39.78 | 32.93 | 32.94 |
| Chanthaburi | 2022 | 11 | 28.46 | 39.74 | 33.14 | 33.25 |
| Chanthaburi | 2022 | 12 | 26.61 | 34.93 | 29.00 | 28.85 |
| Chanthaburi | 2023 | 1 | 26.71 | 34.46 | 28.55 | 28.69 |
| Chanthaburi | 2023 | 2 | 28.19 | 38.53 | 31.96 | 32.40 |
| Chanthaburi | 2023 | 3 | 28.83 | 40.26 | 33.88 | 33.85 |
| Chanthaburi | 2023 | 4 | 30.04 | 42.77 | 37.13 | 35.78 |
| Chanthaburi | 2023 | 5 | 29.74 | 42.85 | 36.97 | 35.86 |
| Chanthaburi | 2023 | 6 | 29.08 | 42.02 | 35.55 | 34.80 |
| Chanthaburi | 2023 | 7 | 28.48 | 41.28 | 34.29 | 34.18 |
| Chanthaburi | 2023 | 8 | 29.37 | 42.20 | 36.04 | 34.58 |
| Chanthaburi | 2023 | 9 | 28.32 | 41.25 | 34.09 | 34.19 |
| Chanthaburi | 2023 | 10 | 28.76 | 41.76 | 35.02 | 34.97 |
| Chanthaburi | 2023 | 11 | 28.40 | 39.06 | 32.71 | 32.05 |
| Chanthaburi | 2023 | 12 | 28.29 | 37.36 | 31.89 | 30.71 |
| Chiang Mai | 2015 | 1 | 19.10 | 22.21 | 23.04 | 18.45 |
| Chiang Mai | 2015 | 2 | 21.74 | 23.85 | 24.60 | 20.44 |
| Chiang Mai | 2015 | 3 | 25.55 | 29.06 | 25.93 | 24.63 |
| Chiang Mai | 2015 | 4 | 26.26 | 32.23 | 27.02 | 27.02 |
| Chiang Mai | 2015 | 5 | 27.44 | 35.57 | 29.13 | 29.30 |
| Chiang Mai | 2015 | 6 | 26.30 | 34.91 | 27.90 | 27.58 |
| Chiang Mai | 2015 | 7 | 25.10 | 34.05 | 25.92 | 26.90 |
| Chiang Mai | 2015 | 8 | 24.80 | 33.91 | 25.37 | 26.67 |
| Chiang Mai | 2015 | 9 | 25.02 | 34.36 | 25.78 | 27.42 |
| Chiang Mai | 2015 | 10 | 23.80 | 31.60 | 24.43 | 25.83 |
| Chiang Mai | 2015 | 11 | 23.47 | 30.36 | 23.87 | 25.34 |
| Chiang Mai | 2015 | 12 | 21.04 | 25.59 | 22.68 | 21.22 |
| Chiang Mai | 2017 | 1 | 20.26 | 25.35 | 20.78 | 21.26 |
| Chiang Mai | 2017 | 2 | 22.13 | 25.43 | 24.15 | 21.90 |
| Chiang Mai | 2017 | 3 | 25.60 | 29.33 | 26.34 | 25.03 |
| Chiang Mai | 2017 | 4 | 26.20 | 32.95 | 27.14 | 27.79 |
| Chiang Mai | 2017 | 5 | 25.74 | 34.15 | 26.66 | 28.22 |
| Chiang Mai | 2017 | 6 | 25.19 | 34.36 | 26.11 | 27.66 |
| Chiang Mai | 2017 | 7 | 24.47 | 33.59 | 24.65 | 27.21 |
| Chiang Mai | 2017 | 8 | 24.50 | 33.65 | 24.71 | 27.03 |
| Chiang Mai | 2017 | 9 | 24.83 | 34.16 | 25.40 | 28.01 |
| Chiang Mai | 2017 | 10 | 23.54 | 32.00 | 23.32 | 26.03 |
| Chiang Mai | 2017 | 11 | 22.22 | 28.78 | 22.57 | 23.59 |
| Chiang Mai | 2017 | 12 | 19.54 | 23.91 | 22.53 | 19.36 |
| Chiang Mai | 2020 | 1 | 21.20 | 24.19 | 24.28 | 20.24 |
| Chiang Mai | 2020 | 2 | 22.66 | 25.24 | 24.86 | 21.78 |
| Chiang Mai | 2020 | 3 | 26.58 | 29.32 | 26.78 | 25.50 |
| Chiang Mai | 2020 | 4 | 27.32 | 31.97 | 27.28 | 27.21 |
| Chiang Mai | 2020 | 5 | 27.73 | 35.97 | 29.65 | 29.80 |
| Chiang Mai | 2020 | 6 | 26.24 | 35.78 | 27.94 | 29.55 |
| Chiang Mai | 2020 | 7 | 25.69 | 35.23 | 27.06 | 28.74 |
| Chiang Mai | 2020 | 8 | 24.38 | 33.82 | 24.43 | 27.24 |
| Chiang Mai | 2020 | 9 | 25.31 | 35.20 | 26.43 | 28.88 |
| Chiang Mai | 2020 | 10 | 23.11 | 30.68 | 22.97 | 24.37 |
| Chiang Mai | 2020 | 11 | 22.91 | 29.22 | 23.62 | 24.28 |
| Chiang Mai | 2020 | 12 | 20.10 | 24.44 | 22.49 | 20.42 |
| Chiang Mai | 2021 | 1 | 19.58 | 23.40 | 22.85 | 19.58 |
| Chiang Mai | 2021 | 2 | 21.88 | 25.31 | 24.14 | 21.76 |
| Chiang Mai | 2021 | 3 | 26.12 | 29.51 | 26.80 | 25.98 |
| Chiang Mai | 2021 | 4 | 25.19 | 31.93 | 25.90 | 26.98 |
| Chiang Mai | 2021 | 5 | 26.79 | 35.65 | 28.69 | 29.81 |
| Chiang Mai | 2021 | 6 | 25.35 | 34.30 | 26.40 | 27.75 |
| Chiang Mai | 2021 | 7 | 24.76 | 33.77 | 25.26 | 27.70 |
| Chiang Mai | 2021 | 8 | 25.01 | 34.21 | 25.75 | 28.33 |
| Chiang Mai | 2021 | 9 | 24.25 | 33.41 | 24.24 | 27.45 |
| Chiang Mai | 2021 | 10 | 23.71 | 32.12 | 23.63 | 26.57 |
| Chiang Mai | 2021 | 11 | 22.97 | 30.58 | 22.75 | 25.89 |
| Chiang Mai | 2021 | 12 | 19.00 | 22.73 | 22.31 | 18.94 |
| Chiang Mai | 2022 | 1 | 20.33 | 23.79 | 23.25 | 20.34 |
| Chiang Mai | 2022 | 2 | 21.76 | 24.98 | 24.12 | 21.63 |
| Chiang Mai | 2022 | 3 | 25.34 | 31.03 | 26.39 | 26.77 |
| Chiang Mai | 2022 | 4 | 24.88 | 31.43 | 26.04 | 26.62 |
| Chiang Mai | 2022 | 5 | 24.30 | 32.23 | 24.70 | 26.71 |
| Chiang Mai | 2022 | 6 | 25.03 | 33.17 | 25.90 | 26.94 |
| Chiang Mai | 2022 | 7 | 25.08 | 34.29 | 25.76 | 28.39 |
| Chiang Mai | 2022 | 8 | 24.45 | 33.59 | 24.62 | 27.53 |
| Chiang Mai | 2022 | 9 | 23.89 | 32.54 | 23.58 | 27.00 |
| Chiang Mai | 2022 | 10 | 23.01 | 30.41 | 22.88 | 25.19 |
| Chiang Mai | 2022 | 11 | 22.64 | 28.91 | 23.29 | 24.62 |
| Chiang Mai | 2022 | 12 | 20.63 | 25.89 | 21.97 | 21.57 |
| Chiang Mai | 2023 | 1 | 19.15 | 21.96 | 23.45 | 18.41 |
| Chiang Mai | 2023 | 2 | 21.59 | 24.35 | 24.31 | 21.17 |
| Chiang Mai | 2023 | 3 | 24.08 | 28.03 | 25.16 | 23.90 |
| Chiang Mai | 2023 | 4 | 28.17 | 33.06 | 28.67 | 28.14 |
| Chiang Mai | 2023 | 5 | 26.91 | 35.15 | 28.51 | 29.11 |
| Chiang Mai | 2023 | 6 | 25.80 | 34.84 | 27.12 | 28.62 |
| Chiang Mai | 2023 | 7 | 25.68 | 35.02 | 27.01 | 28.32 |
| Chiang Mai | 2023 | 8 | 24.72 | 33.88 | 25.14 | 27.16 |
| Chiang Mai | 2023 | 9 | 24.67 | 34.12 | 24.92 | 28.17 |
| Chiang Mai | 2023 | 10 | 24.00 | 32.96 | 23.75 | 27.51 |
| Chiang Mai | 2023 | 11 | 23.09 | 30.10 | 23.97 | 25.09 |
| Chiang Mai | 2023 | 12 | 21.61 | 27.41 | 22.98 | 23.02 |
| Khon Kaen | 2015 | 1 | 23.04 | 27.21 | 25.26 | 22.91 |
| Khon Kaen | 2015 | 2 | 26.05 | 33.30 | 27.98 | 28.10 |
| Khon Kaen | 2015 | 3 | 29.89 | 39.73 | 33.88 | 33.48 |
| Khon Kaen | 2015 | 4 | 30.63 | 39.49 | 34.19 | 33.41 |
| Khon Kaen | 2015 | 5 | 31.56 | 43.39 | 38.37 | 36.18 |
| Khon Kaen | 2015 | 6 | 30.26 | 42.10 | 36.43 | 34.70 |
| Khon Kaen | 2015 | 7 | 28.95 | 40.46 | 33.68 | 32.96 |
| Khon Kaen | 2015 | 8 | 28.62 | 40.95 | 34.12 | 33.87 |
| Khon Kaen | 2015 | 9 | 28.52 | 40.94 | 34.05 | 33.83 |
| Khon Kaen | 2015 | 10 | 27.43 | 37.85 | 30.62 | 31.54 |
| Khon Kaen | 2015 | 11 | 28.07 | 37.99 | 31.55 | 31.61 |
| Khon Kaen | 2015 | 12 | 26.10 | 33.31 | 27.98 | 27.97 |
| Khon Kaen | 2017 | 1 | 25.58 | 32.51 | 27.11 | 27.01 |
| Khon Kaen | 2017 | 2 | 25.61 | 31.18 | 26.98 | 26.20 |
| Khon Kaen | 2017 | 3 | 28.87 | 37.91 | 32.19 | 31.86 |
| Khon Kaen | 2017 | 4 | 30.33 | 40.40 | 34.85 | 33.84 |
| Khon Kaen | 2017 | 5 | 29.50 | 41.80 | 35.65 | 34.84 |
| Khon Kaen | 2017 | 6 | 29.24 | 41.79 | 35.46 | 34.41 |
| Khon Kaen | 2017 | 7 | 27.90 | 39.66 | 32.23 | 32.42 |
| Khon Kaen | 2017 | 8 | 28.56 | 40.99 | 34.04 | 33.80 |
| Khon Kaen | 2017 | 9 | 28.79 | 41.87 | 35.20 | 34.75 |
| Khon Kaen | 2017 | 10 | 27.25 | 38.39 | 31.08 | 31.79 |
| Khon Kaen | 2017 | 11 | 26.50 | 34.78 | 28.88 | 28.83 |
| Khon Kaen | 2017 | 12 | 23.68 | 29.73 | 26.02 | 24.49 |
| Khon Kaen | 2020 | 1 | 26.46 | 32.84 | 27.77 | 28.01 |
| Khon Kaen | 2020 | 2 | 26.53 | 32.55 | 27.74 | 27.50 |
| Khon Kaen | 2020 | 3 | 30.07 | 39.97 | 33.98 | 33.82 |
| Khon Kaen | 2020 | 4 | 30.07 | 40.54 | 34.89 | 34.09 |
| Khon Kaen | 2020 | 5 | 31.98 | 43.79 | 38.89 | 36.72 |
| Khon Kaen | 2020 | 6 | 30.10 | 42.16 | 36.36 | 35.01 |
| Khon Kaen | 2020 | 7 | 29.90 | 42.13 | 36.24 | 35.06 |
| Khon Kaen | 2020 | 8 | 28.62 | 41.14 | 34.25 | 34.03 |
| Khon Kaen | 2020 | 9 | 28.71 | 41.58 | 34.81 | 34.50 |
| Khon Kaen | 2020 | 10 | 26.21 | 36.54 | 28.77 | 30.44 |
| Khon Kaen | 2020 | 11 | 26.18 | 34.56 | 28.17 | 29.19 |
| Khon Kaen | 2020 | 12 | 23.95 | 30.01 | 25.80 | 25.35 |
| Khon Kaen | 2021 | 1 | 22.31 | 26.30 | 25.16 | 22.48 |
| Khon Kaen | 2021 | 2 | 25.46 | 30.99 | 26.41 | 26.77 |
| Khon Kaen | 2021 | 3 | 29.08 | 38.91 | 32.88 | 33.17 |
| Khon Kaen | 2021 | 4 | 29.82 | 40.99 | 34.87 | 34.46 |
| Khon Kaen | 2021 | 5 | 30.62 | 43.34 | 38.02 | 36.33 |
| Khon Kaen | 2021 | 6 | 29.98 | 41.98 | 36.04 | 34.76 |
| Khon Kaen | 2021 | 7 | 29.25 | 41.56 | 35.11 | 33.99 |
| Khon Kaen | 2021 | 8 | 29.59 | 41.89 | 35.80 | 34.53 |
| Khon Kaen | 2021 | 9 | 28.14 | 40.84 | 33.49 | 33.91 |
| Khon Kaen | 2021 | 10 | 27.53 | 38.66 | 31.46 | 32.20 |
| Khon Kaen | 2021 | 11 | 26.38 | 34.59 | 28.59 | 29.21 |
| Khon Kaen | 2021 | 12 | 23.57 | 28.60 | 25.26 | 24.21 |
| Khon Kaen | 2022 | 1 | 25.14 | 31.30 | 26.17 | 26.93 |
| Khon Kaen | 2022 | 2 | 24.99 | 32.31 | 26.83 | 27.28 |
| Khon Kaen | 2022 | 3 | 29.05 | 39.37 | 33.27 | 33.29 |
| Khon Kaen | 2022 | 4 | 29.04 | 38.20 | 33.08 | 32.42 |
| Khon Kaen | 2022 | 5 | 28.29 | 39.84 | 33.33 | 32.99 |
| Khon Kaen | 2022 | 6 | 30.19 | 42.20 | 36.51 | 35.16 |
| Khon Kaen | 2022 | 7 | 29.24 | 41.84 | 35.44 | 34.92 |
| Khon Kaen | 2022 | 8 | 28.48 | 40.76 | 33.88 | 33.79 |
| Khon Kaen | 2022 | 9 | 28.13 | 40.61 | 33.30 | 33.76 |
| Khon Kaen | 2022 | 10 | 26.61 | 35.96 | 29.08 | 30.32 |
| Khon Kaen | 2022 | 11 | 27.11 | 36.30 | 29.83 | 30.83 |
| Khon Kaen | 2022 | 12 | 23.28 | 28.78 | 25.25 | 24.46 |
| Khon Kaen | 2023 | 1 | 22.67 | 27.40 | 24.64 | 23.38 |
| Khon Kaen | 2023 | 2 | 26.59 | 33.47 | 28.67 | 28.54 |
| Khon Kaen | 2023 | 3 | 28.62 | 35.94 | 30.83 | 31.01 |
| Khon Kaen | 2023 | 4 | 32.47 | 42.82 | 37.79 | 36.56 |
| Khon Kaen | 2023 | 5 | 31.28 | 43.30 | 37.94 | 36.61 |
| Khon Kaen | 2023 | 6 | 30.19 | 42.98 | 37.35 | 35.89 |
| Khon Kaen | 2023 | 7 | 29.56 | 41.96 | 35.92 | 35.08 |
| Khon Kaen | 2023 | 8 | 29.19 | 41.81 | 35.44 | 34.88 |
| Khon Kaen | 2023 | 9 | 27.94 | 40.63 | 33.00 | 34.00 |
| Khon Kaen | 2023 | 10 | 28.34 | 40.30 | 33.44 | 33.73 |
| Khon Kaen | 2023 | 11 | 26.80 | 35.89 | 29.88 | 30.32 |
| Khon Kaen | 2023 | 12 | 25.31 | 32.72 | 27.66 | 27.51 |
| Krabi | 2015 | 1 | 27.41 | 37.26 | 30.19 | 30.47 |
| Krabi | 2015 | 2 | 27.91 | 37.41 | 30.85 | 30.73 |
| Krabi | 2015 | 3 | 29.07 | 39.78 | 33.52 | 33.01 |
| Krabi | 2015 | 4 | 29.32 | 41.55 | 35.32 | 34.29 |
| Krabi | 2015 | 5 | 28.66 | 41.06 | 34.28 | 33.27 |
| Krabi | 2015 | 6 | 28.07 | 40.60 | 33.21 | 33.13 |
| Krabi | 2015 | 7 | 27.79 | 40.05 | 32.44 | 30.87 |
| Krabi | 2015 | 8 | 27.34 | 39.29 | 31.27 | 30.91 |
| Krabi | 2015 | 9 | 27.35 | 39.29 | 31.34 | 30.82 |
| Krabi | 2015 | 10 | 27.68 | 39.96 | 32.26 | 32.72 |
| Krabi | 2015 | 11 | 27.39 | 39.40 | 31.42 | 31.93 |
| Krabi | 2015 | 12 | 27.65 | 39.04 | 31.68 | 31.40 |
| Krabi | 2017 | 1 | 27.03 | 38.67 | 30.41 | 30.65 |
| Krabi | 2017 | 2 | 27.74 | 38.74 | 31.58 | 31.27 |
| Krabi | 2017 | 3 | 28.44 | 40.49 | 33.57 | 32.75 |
| Krabi | 2017 | 4 | 28.10 | 40.85 | 33.49 | 33.07 |
| Krabi | 2017 | 5 | 28.07 | 41.22 | 33.73 | 32.98 |
| Krabi | 2017 | 6 | 27.77 | 40.28 | 32.57 | 31.80 |
| Krabi | 2017 | 7 | 27.74 | 40.04 | 32.38 | 31.04 |
| Krabi | 2017 | 8 | 27.27 | 39.57 | 31.32 | 31.06 |
| Krabi | 2017 | 9 | 27.06 | 39.28 | 30.85 | 31.32 |
| Krabi | 2017 | 10 | 27.32 | 39.46 | 31.33 | 32.04 |
| Krabi | 2017 | 11 | 26.97 | 38.96 | 30.46 | 32.00 |
| Krabi | 2017 | 12 | 26.45 | 37.26 | 29.06 | 29.87 |
| Krabi | 2020 | 1 | 29.03 | 39.33 | 33.03 | 31.79 |
| Krabi | 2020 | 2 | 29.49 | 39.52 | 33.51 | 31.68 |
| Krabi | 2020 | 3 | 30.06 | 41.01 | 35.21 | 33.61 |
| Krabi | 2020 | 4 | 30.17 | 42.76 | 37.12 | 35.16 |
| Krabi | 2020 | 5 | 29.18 | 42.26 | 35.96 | 34.52 |
| Krabi | 2020 | 6 | 28.04 | 40.53 | 33.13 | 33.31 |
| Krabi | 2020 | 7 | 27.93 | 40.18 | 32.75 | 32.90 |
| Krabi | 2020 | 8 | 28.37 | 40.57 | 33.58 | 32.09 |
| Krabi | 2020 | 9 | 27.59 | 39.73 | 31.98 | 31.22 |
| Krabi | 2020 | 10 | 26.97 | 38.86 | 30.44 | 30.24 |
| Krabi | 2020 | 11 | 27.85 | 39.97 | 32.45 | 32.84 |
| Krabi | 2020 | 12 | 27.18 | 38.13 | 30.34 | 30.68 |
| Krabi | 2021 | 1 | 27.40 | 37.36 | 30.28 | 29.76 |
| Krabi | 2021 | 2 | 28.37 | 37.60 | 31.36 | 30.61 |
| Krabi | 2021 | 3 | 29.22 | 39.65 | 33.47 | 32.77 |
| Krabi | 2021 | 4 | 28.44 | 40.53 | 33.63 | 32.89 |
| Krabi | 2021 | 5 | 28.33 | 40.73 | 33.64 | 33.17 |
| Krabi | 2021 | 6 | 27.70 | 39.38 | 31.92 | 32.04 |
| Krabi | 2021 | 7 | 27.48 | 39.05 | 31.31 | 30.69 |
| Krabi | 2021 | 8 | 26.93 | 38.38 | 30.17 | 29.86 |
| Krabi | 2021 | 9 | 26.53 | 37.63 | 29.16 | 29.89 |
| Krabi | 2021 | 10 | 26.99 | 38.32 | 30.35 | 30.41 |
| Krabi | 2021 | 11 | 27.33 | 39.69 | 31.56 | 32.60 |
| Krabi | 2021 | 12 | 28.24 | 39.45 | 32.56 | 31.88 |
| Krabi | 2022 | 1 | 29.14 | 40.13 | 33.83 | 33.04 |
| Krabi | 2022 | 2 | 28.80 | 40.56 | 34.54 | 33.35 |
| Krabi | 2022 | 3 | 28.95 | 41.25 | 34.74 | 34.43 |
| Krabi | 2022 | 4 | 29.19 | 42.14 | 35.87 | 34.99 |
| Krabi | 2022 | 5 | 28.67 | 41.91 | 35.13 | 32.97 |
| Krabi | 2022 | 6 | 28.26 | 40.79 | 33.68 | 33.16 |
| Krabi | 2022 | 7 | 28.17 | 40.88 | 33.61 | 32.95 |
| Krabi | 2022 | 8 | 27.93 | 40.33 | 32.82 | 32.44 |
| Krabi | 2022 | 9 | 27.92 | 40.18 | 32.74 | 32.02 |
| Krabi | 2022 | 10 | 27.41 | 39.91 | 31.80 | 32.58 |
| Krabi | 2022 | 11 | 28.06 | 40.67 | 33.30 | 33.55 |
| Krabi | 2022 | 12 | 27.66 | 38.99 | 31.52 | 31.60 |
| Krabi | 2023 | 1 | 27.73 | 38.27 | 31.21 | 31.13 |
| Krabi | 2023 | 2 | 28.79 | 39.17 | 32.75 | 32.15 |
| Krabi | 2023 | 3 | 29.48 | 40.05 | 34.06 | 33.13 |
| Krabi | 2023 | 4 | 30.13 | 42.37 | 36.76 | 34.42 |
| Krabi | 2023 | 5 | 29.33 | 42.18 | 35.97 | 34.02 |
| Krabi | 2023 | 6 | 28.59 | 40.96 | 34.15 | 32.51 |
| Krabi | 2023 | 7 | 28.33 | 40.92 | 33.92 | 30.88 |
| Krabi | 2023 | 8 | 28.22 | 40.79 | 33.58 | 31.01 |
| Krabi | 2023 | 9 | 27.88 | 40.98 | 33.32 | 32.10 |
| Krabi | 2023 | 10 | 28.05 | 40.65 | 33.29 | 32.71 |
| Krabi | 2023 | 11 | 27.79 | 40.86 | 33.08 | 32.81 |
| Krabi | 2023 | 12 | 28.61 | 41.45 | 34.68 | 33.17 |
| Lopburi | 2015 | 1 | 25.04 | 30.28 | 26.45 | 25.76 |
| Lopburi | 2015 | 2 | 28.24 | 36.03 | 30.87 | 30.46 |
| Lopburi | 2015 | 3 | 30.43 | 41.97 | 36.39 | 34.87 |
| Lopburi | 2015 | 4 | 30.98 | 41.56 | 36.31 | 35.00 |
| Lopburi | 2015 | 5 | 32.03 | 44.73 | 40.25 | 36.72 |
| Lopburi | 2015 | 6 | 31.71 | 43.91 | 39.07 | 36.24 |
| Lopburi | 2015 | 7 | 30.76 | 42.82 | 37.41 | 35.26 |
| Lopburi | 2015 | 8 | 29.71 | 42.39 | 36.47 | 35.12 |
| Lopburi | 2015 | 9 | 29.15 | 42.26 | 35.89 | 35.13 |
| Lopburi | 2015 | 10 | 28.63 | 40.91 | 34.22 | 34.33 |
| Lopburi | 2015 | 11 | 29.19 | 40.60 | 34.53 | 34.11 |
| Lopburi | 2015 | 12 | 27.50 | 36.61 | 30.47 | 30.93 |
| Lopburi | 2017 | 1 | 27.46 | 35.63 | 29.57 | 29.56 |
| Lopburi | 2017 | 2 | 27.89 | 34.82 | 29.79 | 29.35 |
| Lopburi | 2017 | 3 | 30.81 | 41.69 | 36.48 | 34.63 |
| Lopburi | 2017 | 4 | 31.60 | 42.97 | 38.01 | 35.69 |
| Lopburi | 2017 | 5 | 30.67 | 44.22 | 39.13 | 36.59 |
| Lopburi | 2017 | 6 | 29.99 | 42.89 | 37.23 | 35.50 |
| Lopburi | 2017 | 7 | 28.98 | 41.71 | 35.18 | 34.40 |
| Lopburi | 2017 | 8 | 29.73 | 42.97 | 37.13 | 35.72 |
| Lopburi | 2017 | 9 | 29.77 | 43.39 | 37.67 | 36.26 |
| Lopburi | 2017 | 10 | 28.80 | 41.04 | 34.69 | 34.32 |
| Lopburi | 2017 | 11 | 27.84 | 37.32 | 31.15 | 31.26 |
| Lopburi | 2017 | 12 | 25.92 | 32.67 | 28.03 | 27.14 |
| Lopburi | 2020 | 1 | 28.71 | 36.92 | 31.19 | 31.51 |
| Lopburi | 2020 | 2 | 29.11 | 36.23 | 31.02 | 30.55 |
| Lopburi | 2020 | 3 | 31.77 | 42.84 | 37.66 | 35.50 |
| Lopburi | 2020 | 4 | 32.66 | 43.95 | 39.32 | 36.67 |
| Lopburi | 2020 | 5 | 32.88 | 46.15 | 42.30 | 38.22 |
| Lopburi | 2020 | 6 | 31.10 | 43.93 | 38.87 | 36.43 |
| Lopburi | 2020 | 7 | 30.91 | 43.83 | 38.78 | 36.62 |
| Lopburi | 2020 | 8 | 29.76 | 42.61 | 36.72 | 35.40 |
| Lopburi | 2020 | 9 | 29.64 | 43.20 | 37.33 | 36.08 |
| Lopburi | 2020 | 10 | 27.10 | 38.37 | 30.74 | 32.21 |
| Lopburi | 2020 | 11 | 27.76 | 36.90 | 30.70 | 31.01 |
| Lopburi | 2020 | 12 | 26.41 | 33.21 | 28.32 | 27.77 |
| Lopburi | 2021 | 1 | 25.07 | 29.78 | 26.77 | 25.11 |
| Lopburi | 2021 | 2 | 28.01 | 34.92 | 29.62 | 29.87 |
| Lopburi | 2021 | 3 | 31.45 | 42.19 | 37.05 | 35.55 |
| Lopburi | 2021 | 4 | 30.58 | 43.58 | 38.22 | 36.43 |
| Lopburi | 2021 | 5 | 31.49 | 44.58 | 39.89 | 37.30 |
| Lopburi | 2021 | 6 | 31.13 | 43.32 | 38.13 | 35.93 |
| Lopburi | 2021 | 7 | 29.95 | 42.78 | 36.98 | 35.41 |
| Lopburi | 2021 | 8 | 30.20 | 43.43 | 37.96 | 36.04 |
| Lopburi | 2021 | 9 | 28.87 | 42.26 | 35.66 | 35.43 |
| Lopburi | 2021 | 10 | 28.88 | 41.55 | 35.14 | 34.99 |
| Lopburi | 2021 | 11 | 28.39 | 38.51 | 32.44 | 32.17 |
| Lopburi | 2021 | 12 | 25.95 | 32.22 | 27.34 | 27.22 |
| Lopburi | 2022 | 1 | 27.15 | 35.03 | 29.07 | 30.17 |
| Lopburi | 2022 | 2 | 27.89 | 36.97 | 31.15 | 31.25 |
| Lopburi | 2022 | 3 | 30.72 | 43.14 | 37.90 | 36.28 |
| Lopburi | 2022 | 4 | 30.58 | 41.59 | 36.81 | 35.07 |
| Lopburi | 2022 | 5 | 30.04 | 42.66 | 37.31 | 35.23 |
| Lopburi | 2022 | 6 | 30.70 | 43.58 | 38.42 | 36.24 |
| Lopburi | 2022 | 7 | 30.11 | 43.59 | 38.08 | 36.13 |
| Lopburi | 2022 | 8 | 29.30 | 42.35 | 36.14 | 35.16 |
| Lopburi | 2022 | 9 | 28.92 | 42.31 | 35.73 | 35.45 |
| Lopburi | 2022 | 10 | 28.08 | 38.65 | 32.07 | 32.56 |
| Lopburi | 2022 | 11 | 28.30 | 38.91 | 32.63 | 33.18 |
| Lopburi | 2022 | 12 | 25.59 | 32.40 | 27.51 | 27.37 |
| Lopburi | 2023 | 1 | 25.73 | 31.53 | 27.33 | 26.63 |
| Lopburi | 2023 | 2 | 28.36 | 36.73 | 31.36 | 31.15 |
| Lopburi | 2023 | 3 | 30.48 | 40.05 | 35.05 | 33.90 |
| Lopburi | 2023 | 4 | 32.84 | 45.25 | 40.99 | 37.89 |
| Lopburi | 2023 | 5 | 32.21 | 45.18 | 40.79 | 37.87 |
| Lopburi | 2023 | 6 | 31.33 | 44.14 | 39.28 | 36.41 |
| Lopburi | 2023 | 7 | 30.72 | 43.52 | 38.32 | 36.19 |
| Lopburi | 2023 | 8 | 30.78 | 43.77 | 38.70 | 36.13 |
| Lopburi | 2023 | 9 | 29.52 | 43.14 | 37.21 | 35.87 |
| Lopburi | 2023 | 10 | 29.53 | 42.68 | 36.69 | 35.80 |
| Lopburi | 2023 | 11 | 28.23 | 38.24 | 32.59 | 32.26 |
| Lopburi | 2023 | 12 | 27.73 | 35.80 | 30.60 | 30.22 |
| Pathum Thani | 2015 | 1 | 26.39 | 34.45 | 28.73 | 28.57 |
| Pathum Thani | 2015 | 2 | 28.53 | 38.05 | 32.06 | 31.35 |
| Pathum Thani | 2015 | 3 | 30.22 | 42.34 | 36.74 | 34.09 |
| Pathum Thani | 2015 | 4 | 31.05 | 42.81 | 37.66 | 34.96 |
| Pathum Thani | 2015 | 5 | 32.23 | 45.39 | 41.19 | 36.24 |
| Pathum Thani | 2015 | 6 | 31.24 | 43.45 | 38.43 | 34.54 |
| Pathum Thani | 2015 | 7 | 30.98 | 43.59 | 38.40 | 34.53 |
| Pathum Thani | 2015 | 8 | 30.93 | 43.55 | 38.40 | 34.80 |
| Pathum Thani | 2015 | 9 | 29.71 | 42.35 | 36.38 | 33.96 |
| Pathum Thani | 2015 | 10 | 29.86 | 43.00 | 37.23 | 35.44 |
| Pathum Thani | 2015 | 11 | 30.33 | 42.69 | 37.26 | 35.35 |
| Pathum Thani | 2015 | 12 | 29.09 | 39.24 | 33.51 | 32.52 |
| Pathum Thani | 2017 | 1 | 28.26 | 37.62 | 31.28 | 31.42 |
| Pathum Thani | 2017 | 2 | 28.99 | 37.83 | 32.18 | 31.81 |
| Pathum Thani | 2017 | 3 | 30.84 | 43.01 | 37.84 | 34.99 |
| Pathum Thani | 2017 | 4 | 31.70 | 44.35 | 39.67 | 36.24 |
| Pathum Thani | 2017 | 5 | 30.74 | 45.63 | 41.14 | 36.86 |
| Pathum Thani | 2017 | 6 | 30.87 | 44.06 | 39.07 | 35.66 |
| Pathum Thani | 2017 | 7 | 30.08 | 43.11 | 37.53 | 34.60 |
| Pathum Thani | 2017 | 8 | 30.45 | 43.61 | 38.29 | 35.43 |
| Pathum Thani | 2017 | 9 | 30.57 | 44.13 | 39.06 | 35.79 |
| Pathum Thani | 2017 | 10 | 29.74 | 42.67 | 36.94 | 35.29 |
| Pathum Thani | 2017 | 11 | 28.88 | 39.42 | 33.63 | 32.33 |
| Pathum Thani | 2017 | 12 | 26.81 | 34.65 | 29.54 | 28.68 |
| Pathum Thani | 2020 | 1 | 29.65 | 40.49 | 34.59 | 33.93 |
| Pathum Thani | 2020 | 2 | 29.78 | 39.78 | 34.05 | 33.06 |
| Pathum Thani | 2020 | 3 | 31.85 | 44.16 | 39.40 | 35.90 |
| Pathum Thani | 2020 | 4 | 32.24 | 45.16 | 40.80 | 36.98 |
| Pathum Thani | 2020 | 5 | 32.93 | 47.68 | 44.61 | 38.83 |
| Pathum Thani | 2020 | 6 | 30.99 | 45.22 | 40.61 | 36.75 |
| Pathum Thani | 2020 | 7 | 30.89 | 45.02 | 40.43 | 36.84 |
| Pathum Thani | 2020 | 8 | 30.59 | 43.33 | 38.01 | 35.48 |
| Pathum Thani | 2020 | 9 | 30.13 | 43.79 | 38.34 | 36.06 |
| Pathum Thani | 2020 | 10 | 28.10 | 40.00 | 32.85 | 32.97 |
| Pathum Thani | 2020 | 11 | 29.00 | 39.22 | 33.30 | 32.87 |
| Pathum Thani | 2020 | 12 | 27.79 | 35.88 | 30.47 | 30.21 |
| Pathum Thani | 2021 | 1 | 26.37 | 33.92 | 28.72 | 28.99 |
| Pathum Thani | 2021 | 2 | 28.84 | 39.01 | 32.80 | 32.92 |
| Pathum Thani | 2021 | 3 | 31.35 | 45.34 | 40.99 | 37.01 |
| Pathum Thani | 2021 | 4 | 30.77 | 44.75 | 39.91 | 36.62 |
| Pathum Thani | 2021 | 5 | 31.62 | 45.33 | 40.96 | 37.04 |
| Pathum Thani | 2021 | 6 | 31.59 | 43.74 | 38.83 | 35.81 |
| Pathum Thani | 2021 | 7 | 30.43 | 42.88 | 37.32 | 35.08 |
| Pathum Thani | 2021 | 8 | 30.37 | 43.14 | 37.62 | 35.46 |
| Pathum Thani | 2021 | 9 | 29.14 | 42.20 | 35.82 | 35.12 |
| Pathum Thani | 2021 | 10 | 29.41 | 42.14 | 36.06 | 34.75 |
| Pathum Thani | 2021 | 11 | 29.11 | 40.28 | 34.44 | 33.50 |
| Pathum Thani | 2021 | 12 | 27.18 | 34.48 | 29.05 | 28.85 |
| Pathum Thani | 2022 | 1 | 28.31 | 37.63 | 31.34 | 32.01 |
| Pathum Thani | 2022 | 2 | 28.69 | 39.37 | 33.18 | 32.85 |
| Pathum Thani | 2022 | 3 | 30.81 | 44.40 | 39.48 | 36.43 |
| Pathum Thani | 2022 | 4 | 30.95 | 43.21 | 38.47 | 35.61 |
| Pathum Thani | 2022 | 5 | 30.57 | 43.38 | 38.26 | 35.30 |
| Pathum Thani | 2022 | 6 | 30.95 | 44.91 | 40.26 | 36.66 |
| Pathum Thani | 2022 | 7 | 30.43 | 44.78 | 39.85 | 36.46 |
| Pathum Thani | 2022 | 8 | 29.73 | 43.10 | 37.24 | 35.02 |
| Pathum Thani | 2022 | 9 | 29.14 | 41.89 | 35.48 | 34.56 |
| Pathum Thani | 2022 | 10 | 28.93 | 40.30 | 34.08 | 33.55 |
| Pathum Thani | 2022 | 11 | 29.20 | 40.58 | 34.58 | 33.92 |
| Pathum Thani | 2022 | 12 | 26.93 | 34.89 | 29.38 | 29.44 |
| Pathum Thani | 2023 | 1 | 26.82 | 34.64 | 28.92 | 29.29 |
| Pathum Thani | 2023 | 2 | 28.80 | 38.63 | 32.69 | 32.51 |
| Pathum Thani | 2023 | 3 | 30.63 | 42.55 | 37.37 | 35.67 |
| Pathum Thani | 2023 | 4 | 32.70 | 45.84 | 41.77 | 38.30 |
| Pathum Thani | 2023 | 5 | 32.53 | 46.85 | 43.28 | 38.79 |
| Pathum Thani | 2023 | 6 | 31.58 | 44.65 | 40.01 | 36.65 |
| Pathum Thani | 2023 | 7 | 30.91 | 43.25 | 38.02 | 35.32 |
| Pathum Thani | 2023 | 8 | 31.40 | 44.49 | 39.82 | 36.30 |
| Pathum Thani | 2023 | 9 | 30.38 | 44.03 | 38.76 | 36.05 |
| Pathum Thani | 2023 | 10 | 30.08 | 44.04 | 38.64 | 36.40 |
| Pathum Thani | 2023 | 11 | 29.03 | 40.19 | 34.46 | 33.46 |
| Pathum Thani | 2023 | 12 | 28.69 | 38.62 | 33.18 | 32.48 |
| Phetchabun | 2015 | 1 | 24.06 | 28.87 | 25.52 | 25.85 |
| Phetchabun | 2015 | 2 | 26.92 | 34.19 | 28.76 | 30.22 |
| Phetchabun | 2015 | 3 | 30.05 | 40.34 | 34.55 | 34.99 |
| Phetchabun | 2015 | 4 | 30.48 | 40.07 | 34.47 | 34.95 |
| Phetchabun | 2015 | 5 | 31.38 | 43.28 | 38.19 | 37.08 |
| Phetchabun | 2015 | 6 | 31.18 | 42.53 | 37.24 | 36.44 |
| Phetchabun | 2015 | 7 | 29.59 | 41.16 | 34.89 | 35.09 |
| Phetchabun | 2015 | 8 | 28.96 | 40.98 | 34.43 | 34.98 |
| Phetchabun | 2015 | 9 | 28.96 | 41.60 | 35.09 | 35.46 |
| Phetchabun | 2015 | 10 | 28.50 | 40.20 | 33.49 | 34.45 |
| Phetchabun | 2015 | 11 | 28.80 | 39.02 | 32.77 | 33.83 |
| Phetchabun | 2015 | 12 | 26.93 | 35.19 | 29.16 | 30.81 |
| Phetchabun | 2017 | 1 | 26.89 | 34.23 | 28.59 | 30.05 |
| Phetchabun | 2017 | 2 | 27.06 | 32.82 | 28.29 | 29.33 |
| Phetchabun | 2017 | 3 | 30.06 | 39.09 | 33.57 | 34.22 |
| Phetchabun | 2017 | 4 | 31.32 | 41.62 | 36.34 | 36.16 |
| Phetchabun | 2017 | 5 | 30.28 | 42.94 | 37.38 | 36.56 |
| Phetchabun | 2017 | 6 | 29.48 | 41.97 | 35.86 | 35.53 |
| Phetchabun | 2017 | 7 | 28.06 | 40.14 | 32.83 | 33.81 |
| Phetchabun | 2017 | 8 | 28.81 | 41.25 | 34.57 | 34.89 |
| Phetchabun | 2017 | 9 | 29.11 | 41.90 | 35.52 | 35.59 |
| Phetchabun | 2017 | 10 | 28.12 | 39.49 | 32.60 | 33.68 |
| Phetchabun | 2017 | 11 | 27.61 | 36.75 | 30.56 | 31.80 |
| Phetchabun | 2017 | 12 | 25.08 | 31.80 | 27.16 | 27.76 |
| Phetchabun | 2020 | 1 | 27.72 | 34.73 | 29.31 | 30.86 |
| Phetchabun | 2020 | 2 | 28.15 | 34.20 | 29.25 | 30.43 |
| Phetchabun | 2020 | 3 | 30.73 | 40.65 | 35.02 | 35.33 |
| Phetchabun | 2020 | 4 | 31.79 | 42.42 | 37.29 | 36.64 |
| Phetchabun | 2020 | 5 | 32.37 | 44.88 | 40.43 | 38.40 |
| Phetchabun | 2020 | 6 | 30.37 | 42.85 | 37.28 | 36.56 |
| Phetchabun | 2020 | 7 | 30.27 | 42.97 | 37.45 | 36.66 |
| Phetchabun | 2020 | 8 | 28.88 | 41.38 | 34.73 | 35.15 |
| Phetchabun | 2020 | 9 | 29.54 | 42.61 | 36.58 | 36.24 |
| Phetchabun | 2020 | 10 | 27.24 | 38.08 | 30.69 | 32.45 |
| Phetchabun | 2020 | 11 | 27.77 | 36.48 | 30.40 | 31.42 |
| Phetchabun | 2020 | 12 | 25.82 | 32.13 | 27.41 | 27.95 |
| Phetchabun | 2021 | 1 | 24.45 | 29.23 | 26.16 | 25.72 |
| Phetchabun | 2021 | 2 | 27.07 | 33.37 | 28.19 | 29.50 |
| Phetchabun | 2021 | 3 | 30.95 | 40.76 | 35.31 | 35.43 |
| Phetchabun | 2021 | 4 | 29.82 | 41.92 | 35.91 | 35.73 |
| Phetchabun | 2021 | 5 | 30.95 | 43.97 | 38.96 | 37.32 |
| Phetchabun | 2021 | 6 | 30.39 | 42.61 | 36.97 | 36.21 |
| Phetchabun | 2021 | 7 | 29.37 | 41.97 | 35.67 | 35.49 |
| Phetchabun | 2021 | 8 | 29.53 | 42.30 | 36.21 | 35.89 |
| Phetchabun | 2021 | 9 | 28.67 | 41.70 | 34.92 | 35.24 |
| Phetchabun | 2021 | 10 | 28.31 | 40.21 | 33.32 | 34.21 |
| Phetchabun | 2021 | 11 | 28.20 | 37.78 | 31.63 | 32.51 |
| Phetchabun | 2021 | 12 | 24.98 | 30.66 | 26.31 | 26.82 |
| Phetchabun | 2022 | 1 | 26.35 | 33.26 | 27.53 | 29.25 |
| Phetchabun | 2022 | 2 | 27.13 | 35.52 | 29.61 | 30.79 |
| Phetchabun | 2022 | 3 | 30.23 | 41.67 | 36.06 | 35.68 |
| Phetchabun | 2022 | 4 | 30.00 | 40.65 | 35.40 | 34.90 |
| Phetchabun | 2022 | 5 | 29.92 | 41.78 | 36.14 | 35.56 |
| Phetchabun | 2022 | 6 | 30.14 | 42.24 | 36.57 | 36.00 |
| Phetchabun | 2022 | 7 | 29.55 | 42.43 | 36.35 | 36.02 |
| Phetchabun | 2022 | 8 | 28.79 | 41.31 | 34.66 | 34.88 |
| Phetchabun | 2022 | 9 | 28.55 | 41.51 | 34.58 | 35.16 |
| Phetchabun | 2022 | 10 | 27.92 | 37.97 | 31.39 | 32.59 |
| Phetchabun | 2022 | 11 | 28.12 | 38.39 | 31.98 | 33.15 |
| Phetchabun | 2022 | 12 | 25.36 | 32.22 | 27.20 | 28.16 |
| Phetchabun | 2023 | 1 | 24.97 | 30.85 | 26.42 | 27.17 |
| Phetchabun | 2023 | 2 | 27.59 | 35.17 | 29.62 | 30.86 |
| Phetchabun | 2023 | 3 | 30.13 | 38.15 | 32.97 | 33.62 |
| Phetchabun | 2023 | 4 | 32.77 | 43.32 | 38.52 | 37.62 |
| Phetchabun | 2023 | 5 | 31.84 | 44.48 | 39.76 | 38.01 |
| Phetchabun | 2023 | 6 | 30.71 | 43.33 | 38.03 | 36.71 |
| Phetchabun | 2023 | 7 | 30.48 | 43.19 | 37.81 | 36.63 |
| Phetchabun | 2023 | 8 | 29.59 | 42.33 | 36.27 | 35.82 |
| Phetchabun | 2023 | 9 | 28.82 | 41.97 | 35.31 | 35.46 |
| Phetchabun | 2023 | 10 | 29.04 | 41.78 | 35.37 | 35.35 |
| Phetchabun | 2023 | 11 | 27.94 | 37.57 | 31.70 | 32.05 |
| Phetchabun | 2023 | 12 | 27.22 | 35.03 | 29.71 | 30.28 |
| Phetchaburi | 2015 | 1 | 25.09 | 32.51 | 26.56 | 24.97 |
| Phetchaburi | 2015 | 2 | 27.30 | 36.74 | 30.28 | 27.87 |
| Phetchaburi | 2015 | 3 | 29.45 | 41.73 | 35.69 | 30.03 |
| Phetchaburi | 2015 | 4 | 29.19 | 40.42 | 34.30 | 31.19 |
| Phetchaburi | 2015 | 5 | 31.18 | 44.10 | 39.21 | 34.19 |
| Phetchaburi | 2015 | 6 | 30.50 | 43.30 | 37.94 | 34.28 |
| Phetchaburi | 2015 | 7 | 30.49 | 43.44 | 38.11 | 33.63 |
| Phetchaburi | 2015 | 8 | 30.30 | 42.65 | 37.11 | 34.47 |
| Phetchaburi | 2015 | 9 | 29.53 | 42.13 | 36.04 | 33.46 |
| Phetchaburi | 2015 | 10 | 28.98 | 41.28 | 34.80 | 32.78 |
| Phetchaburi | 2015 | 11 | 29.17 | 41.15 | 34.79 | 32.61 |
| Phetchaburi | 2015 | 12 | 28.02 | 38.48 | 31.94 | 30.02 |
| Phetchaburi | 2017 | 1 | 26.88 | 36.18 | 29.11 | 28.54 |
| Phetchaburi | 2017 | 2 | 27.48 | 36.43 | 30.34 | 29.44 |
| Phetchaburi | 2017 | 3 | 29.67 | 41.34 | 35.45 | 31.51 |
| Phetchaburi | 2017 | 4 | 30.02 | 42.02 | 36.27 | 32.82 |
| Phetchaburi | 2017 | 5 | 29.99 | 43.24 | 37.61 | 34.33 |
| Phetchaburi | 2017 | 6 | 30.62 | 42.73 | 37.37 | 34.40 |
| Phetchaburi | 2017 | 7 | 29.43 | 41.50 | 35.36 | 33.20 |
| Phetchaburi | 2017 | 8 | 29.98 | 42.48 | 36.77 | 34.39 |
| Phetchaburi | 2017 | 9 | 30.15 | 43.23 | 37.70 | 34.54 |
| Phetchaburi | 2017 | 10 | 29.07 | 41.73 | 35.30 | 32.81 |
| Phetchaburi | 2017 | 11 | 27.58 | 38.72 | 31.38 | 29.40 |
| Phetchaburi | 2017 | 12 | 25.90 | 34.67 | 28.44 | 25.48 |
| Phetchaburi | 2020 | 1 | 28.38 | 38.79 | 32.39 | 30.81 |
| Phetchaburi | 2020 | 2 | 28.76 | 38.91 | 32.71 | 30.14 |
| Phetchaburi | 2020 | 3 | 30.88 | 44.16 | 39.31 | 31.64 |
| Phetchaburi | 2020 | 4 | 31.05 | 44.33 | 39.52 | 33.07 |
| Phetchaburi | 2020 | 5 | 31.91 | 45.57 | 41.39 | 34.84 |
| Phetchaburi | 2020 | 6 | 30.91 | 43.80 | 38.76 | 34.66 |
| Phetchaburi | 2020 | 7 | 30.75 | 43.26 | 38.02 | 34.51 |
| Phetchaburi | 2020 | 8 | 30.17 | 42.78 | 37.22 | 33.95 |
| Phetchaburi | 2020 | 9 | 29.74 | 42.85 | 37.06 | 33.71 |
| Phetchaburi | 2020 | 10 | 27.22 | 38.81 | 30.79 | 29.99 |
| Phetchaburi | 2020 | 11 | 27.85 | 38.87 | 31.90 | 29.83 |
| Phetchaburi | 2020 | 12 | 26.31 | 35.74 | 28.64 | 27.07 |
| Phetchaburi | 2021 | 1 | 24.36 | 31.62 | 25.67 | 24.44 |
| Phetchaburi | 2021 | 2 | 26.16 | 34.66 | 27.80 | 26.88 |
| Phetchaburi | 2021 | 3 | 27.93 | 38.72 | 31.68 | 28.05 |
| Phetchaburi | 2021 | 4 | 28.33 | 39.55 | 32.64 | 33.70 |
| Phetchaburi | 2021 | 5 | 29.17 | 40.60 | 34.25 | 35.25 |
| Phetchaburi | 2021 | 6 | 28.97 | 39.99 | 33.58 | 34.80 |
| Phetchaburi | 2021 | 7 | 28.64 | 40.73 | 34.00 | 35.11 |
| Phetchaburi | 2021 | 8 | 28.97 | 40.66 | 34.18 | 35.21 |
| Phetchaburi | 2021 | 9 | 28.12 | 40.00 | 32.80 | 34.47 |
| Phetchaburi | 2021 | 10 | 28.25 | 40.49 | 33.35 | 34.81 |
| Phetchaburi | 2021 | 11 | 27.90 | 39.22 | 32.16 | 33.92 |
| Phetchaburi | 2021 | 12 | 25.82 | 33.84 | 27.44 | 29.87 |
| Phetchaburi | 2022 | 1 | 25.95 | 34.83 | 27.56 | 30.52 |
| Phetchaburi | 2022 | 2 | 26.86 | 36.65 | 29.22 | 31.97 |
| Phetchaburi | 2022 | 3 | 28.42 | 40.33 | 33.46 | 34.79 |
| Phetchaburi | 2022 | 4 | 28.79 | 39.79 | 33.56 | 34.62 |
| Phetchaburi | 2022 | 5 | 28.39 | 40.30 | 33.44 | 34.75 |
| Phetchaburi | 2022 | 6 | 29.13 | 41.16 | 34.80 | 35.57 |
| Phetchaburi | 2022 | 7 | 28.84 | 39.95 | 33.46 | 34.73 |
| Phetchaburi | 2022 | 8 | 28.35 | 39.33 | 32.55 | 34.16 |
| Phetchaburi | 2022 | 9 | 27.70 | 38.69 | 31.39 | 33.52 |
| Phetchaburi | 2022 | 10 | 27.75 | 38.76 | 31.42 | 33.58 |
| Phetchaburi | 2022 | 11 | 27.96 | 38.66 | 31.76 | 33.61 |
| Phetchaburi | 2022 | 12 | 26.14 | 34.26 | 28.42 | 30.26 |
| Phetchaburi | 2023 | 1 | 25.75 | 32.92 | 27.23 | 29.30 |
| Phetchaburi | 2023 | 2 | 26.95 | 36.50 | 29.51 | 31.91 |
| Phetchaburi | 2023 | 3 | 28.40 | 39.88 | 33.45 | 34.51 |
| Phetchaburi | 2023 | 4 | 30.43 | 43.05 | 37.64 | 37.22 |
| Phetchaburi | 2023 | 5 | 31.10 | 43.28 | 38.14 | 37.62 |
| Phetchaburi | 2023 | 6 | 30.55 | 42.34 | 36.85 | 36.84 |
| Phetchaburi | 2023 | 7 | 30.06 | 41.47 | 35.63 | 36.13 |
| Phetchaburi | 2023 | 8 | 29.91 | 41.20 | 35.30 | 35.91 |
| Phetchaburi | 2023 | 9 | 29.16 | 40.84 | 34.46 | 35.39 |
| Phetchaburi | 2023 | 10 | 28.21 | 39.94 | 32.86 | 34.46 |
| Phetchaburi | 2023 | 11 | 27.14 | 37.06 | 30.31 | 32.32 |
| Phetchaburi | 2023 | 12 | 26.60 | 35.15 | 29.12 | 30.97 |
| Samut Prakan | 2015 | 1 | 26.17 | 33.47 | 28.17 | 26.41 |
| Samut Prakan | 2015 | 2 | 27.94 | 36.91 | 30.75 | 27.79 |
| Samut Prakan | 2015 | 3 | 29.29 | 41.06 | 34.85 | 29.61 |
| Samut Prakan | 2015 | 4 | 30.26 | 41.83 | 36.28 | 31.87 |
| Samut Prakan | 2015 | 5 | 31.06 | 43.06 | 37.91 | 32.60 |
| Samut Prakan | 2015 | 6 | 30.14 | 41.35 | 35.61 | 31.80 |
| Samut Prakan | 2015 | 7 | 29.94 | 41.00 | 35.09 | 30.74 |
| Samut Prakan | 2015 | 8 | 29.88 | 40.62 | 34.67 | 31.73 |
| Samut Prakan | 2015 | 9 | 29.24 | 40.65 | 34.33 | 31.74 |
| Samut Prakan | 2015 | 10 | 29.12 | 40.54 | 34.16 | 32.55 |
| Samut Prakan | 2015 | 11 | 29.80 | 40.88 | 35.00 | 32.86 |
| Samut Prakan | 2015 | 12 | 28.65 | 38.39 | 32.50 | 30.82 |
| Samut Prakan | 2017 | 1 | 27.80 | 36.35 | 30.08 | 28.35 |
| Samut Prakan | 2017 | 2 | 28.24 | 36.61 | 30.91 | 28.15 |
| Samut Prakan | 2017 | 3 | 29.72 | 40.95 | 35.01 | 30.18 |
| Samut Prakan | 2017 | 4 | 30.58 | 41.91 | 36.36 | 32.00 |
| Samut Prakan | 2017 | 5 | 29.98 | 42.44 | 36.71 | 32.35 |
| Samut Prakan | 2017 | 6 | 29.94 | 41.06 | 35.17 | 31.47 |
| Samut Prakan | 2017 | 7 | 28.95 | 39.79 | 33.38 | 30.11 |
| Samut Prakan | 2017 | 8 | 29.40 | 40.68 | 34.53 | 30.59 |
| Samut Prakan | 2017 | 9 | 29.74 | 41.51 | 35.54 | 32.23 |
| Samut Prakan | 2017 | 10 | 29.12 | 40.64 | 34.35 | 32.07 |
| Samut Prakan | 2017 | 11 | 28.25 | 38.09 | 31.85 | 29.34 |
| Samut Prakan | 2017 | 12 | 26.47 | 33.82 | 28.81 | 25.58 |
| Samut Prakan | 2020 | 1 | 29.03 | 39.80 | 33.68 | 31.02 |
| Samut Prakan | 2020 | 2 | 29.05 | 38.46 | 32.49 | 29.16 |
| Samut Prakan | 2020 | 3 | 30.10 | 42.51 | 36.85 | 30.77 |
| Samut Prakan | 2020 | 4 | 30.73 | 43.71 | 38.59 | 32.66 |
| Samut Prakan | 2020 | 5 | 31.51 | 45.39 | 41.08 | 33.79 |
| Samut Prakan | 2020 | 6 | 29.95 | 42.52 | 36.75 | 32.50 |
| Samut Prakan | 2020 | 7 | 29.82 | 42.42 | 36.59 | 32.94 |
| Samut Prakan | 2020 | 8 | 29.59 | 41.78 | 35.73 | 31.83 |
| Samut Prakan | 2020 | 9 | 29.33 | 42.51 | 36.34 | 33.26 |
| Samut Prakan | 2020 | 10 | 27.67 | 39.08 | 31.63 | 31.22 |
| Samut Prakan | 2020 | 11 | 28.28 | 38.66 | 32.24 | 30.08 |
| Samut Prakan | 2020 | 12 | 27.06 | 35.21 | 29.49 | 27.47 |
| Samut Prakan | 2021 | 1 | 25.75 | 32.73 | 27.83 | 26.09 |
| Samut Prakan | 2021 | 2 | 28.05 | 37.83 | 31.39 | 29.75 |
| Samut Prakan | 2021 | 3 | 29.67 | 42.09 | 36.22 | 31.55 |
| Samut Prakan | 2021 | 4 | 29.79 | 43.10 | 37.36 | 34.18 |
| Samut Prakan | 2021 | 5 | 30.43 | 43.57 | 38.32 | 35.12 |
| Samut Prakan | 2021 | 6 | 30.27 | 42.16 | 36.53 | 33.61 |
| Samut Prakan | 2021 | 7 | 29.47 | 41.87 | 35.76 | 31.92 |
| Samut Prakan | 2021 | 8 | 29.59 | 42.39 | 36.36 | 32.54 |
| Samut Prakan | 2021 | 9 | 28.67 | 41.20 | 34.42 | 32.27 |
| Samut Prakan | 2021 | 10 | 28.78 | 40.94 | 34.37 | 31.99 |
| Samut Prakan | 2021 | 11 | 28.54 | 39.76 | 33.43 | 31.11 |
| Samut Prakan | 2021 | 12 | 26.55 | 34.27 | 28.46 | 26.84 |
| Samut Prakan | 2022 | 1 | 28.08 | 37.47 | 31.10 | 30.18 |
| Samut Prakan | 2022 | 2 | 28.34 | 38.76 | 32.36 | 30.53 |
| Samut Prakan | 2022 | 3 | 29.65 | 43.08 | 37.32 | 32.97 |
| Samut Prakan | 2022 | 4 | 29.97 | 42.29 | 37.18 | 32.55 |
| Samut Prakan | 2022 | 5 | 29.48 | 41.99 | 36.17 | 31.70 |
| Samut Prakan | 2022 | 6 | 29.83 | 42.63 | 36.83 | 33.26 |
| Samut Prakan | 2022 | 7 | 29.46 | 42.49 | 36.42 | 32.35 |
| Samut Prakan | 2022 | 8 | 29.07 | 41.40 | 34.99 | 31.97 |
| Samut Prakan | 2022 | 9 | 28.44 | 40.68 | 33.66 | 32.16 |
| Samut Prakan | 2022 | 10 | 28.34 | 38.72 | 32.30 | 30.74 |
| Samut Prakan | 2022 | 11 | 28.65 | 39.54 | 33.26 | 32.03 |
| Samut Prakan | 2022 | 12 | 26.56 | 34.67 | 28.85 | 27.31 |
| Samut Prakan | 2023 | 1 | 26.44 | 33.87 | 28.24 | 26.97 |
| Samut Prakan | 2023 | 2 | 28.03 | 37.49 | 31.34 | 29.48 |
| Samut Prakan | 2023 | 3 | 29.26 | 40.27 | 34.38 | 31.15 |
| Samut Prakan | 2023 | 4 | 31.06 | 44.66 | 39.94 | 34.34 |
| Samut Prakan | 2023 | 5 | 31.27 | 44.56 | 39.83 | 34.60 |
| Samut Prakan | 2023 | 6 | 30.66 | 44.07 | 39.00 | 33.27 |
| Samut Prakan | 2023 | 7 | 30.23 | 43.04 | 37.54 | 32.68 |
| Samut Prakan | 2023 | 8 | 30.20 | 42.40 | 36.76 | 31.85 |
| Samut Prakan | 2023 | 9 | 29.47 | 41.78 | 35.64 | 32.22 |
| Samut Prakan | 2023 | 10 | 29.57 | 42.03 | 35.98 | 33.63 |
| Samut Prakan | 2023 | 11 | 28.65 | 39.07 | 33.08 | 30.65 |
| Samut Prakan | 2023 | 12 | 28.35 | 38.06 | 32.36 | 30.23 |
| Samut Songkhram | 2020 | 1 | 27.79 | 37.77 | 31.09 | 30.32 |
| Samut Songkhram | 2020 | 2 | 27.98 | 38.00 | 31.42 | 29.17 |
| Samut Songkhram | 2020 | 3 | 29.25 | 41.94 | 35.75 | 29.75 |
| Samut Songkhram | 2020 | 4 | 29.91 | 43.77 | 38.29 | 32.18 |
| Samut Songkhram | 2020 | 5 | 30.67 | 46.21 | 42.07 | 34.95 |
| Samut Songkhram | 2020 | 6 | 29.74 | 42.22 | 36.31 | 33.06 |
| Samut Songkhram | 2020 | 7 | 29.75 | 41.67 | 35.72 | 32.62 |
| Samut Songkhram | 2020 | 8 | 29.30 | 41.03 | 34.74 | 32.24 |
| Samut Songkhram | 2020 | 9 | 29.19 | 40.96 | 34.61 | 32.16 |
| Samut Songkhram | 2020 | 10 | 27.02 | 37.92 | 30.05 | 30.21 |
| Samut Songkhram | 2020 | 11 | 27.67 | 37.24 | 30.70 | 29.70 |
| Samut Songkhram | 2020 | 12 | 26.56 | 34.33 | 28.79 | 27.42 |
| Samut Songkhram | 2021 | 1 | 24.96 | 31.57 | 26.77 | 25.29 |
| Samut Songkhram | 2021 | 2 | 27.00 | 35.87 | 29.27 | 28.29 |
| Samut Songkhram | 2021 | 3 | 28.97 | 40.15 | 33.73 | 29.69 |
| Samut Songkhram | 2021 | 4 | 29.31 | 41.07 | 34.81 | 31.99 |
| Samut Songkhram | 2021 | 5 | 30.25 | 42.30 | 36.71 | 32.81 |
| Samut Songkhram | 2021 | 6 | 30.54 | 42.11 | 36.59 | 33.02 |
| Samut Songkhram | 2021 | 7 | 29.35 | 41.03 | 34.81 | 31.99 |
| Samut Songkhram | 2021 | 8 | 29.69 | 41.73 | 35.73 | 32.72 |
| Samut Songkhram | 2021 | 9 | 28.56 | 40.53 | 33.64 | 32.04 |
| Samut Songkhram | 2021 | 10 | 28.45 | 40.31 | 33.44 | 31.76 |
| Samut Songkhram | 2021 | 11 | 27.97 | 38.67 | 31.97 | 30.98 |
| Samut Songkhram | 2021 | 12 | 26.08 | 33.19 | 27.63 | 26.73 |
| Samut Songkhram | 2022 | 1 | 27.00 | 35.80 | 29.23 | 28.74 |
| Samut Songkhram | 2022 | 2 | 27.78 | 37.86 | 31.12 | 29.73 |
| Samut Songkhram | 2022 | 3 | 29.25 | 41.21 | 34.96 | 31.37 |
| Samut Songkhram | 2022 | 4 | 29.57 | 40.90 | 35.25 | 31.74 |
| Samut Songkhram | 2022 | 5 | 29.15 | 41.20 | 35.01 | 32.27 |
| Samut Songkhram | 2022 | 6 | 29.79 | 41.99 | 36.09 | 33.24 |
| Samut Songkhram | 2022 | 7 | 29.33 | 41.26 | 35.06 | 32.48 |
| Samut Songkhram | 2022 | 8 | 28.64 | 40.58 | 33.87 | 32.47 |
| Samut Songkhram | 2022 | 9 | 28.92 | 41.40 | 34.84 | 34.27 |
| Samut Songkhram | 2022 | 10 | 27.67 | 38.91 | 31.67 | 32.51 |
| Samut Songkhram | 2022 | 11 | 28.02 | 39.05 | 32.14 | 33.14 |
| Samut Songkhram | 2022 | 12 | 25.73 | 33.96 | 27.76 | 28.44 |
| Samut Songkhram | 2023 | 1 | 25.64 | 33.32 | 27.26 | 27.51 |
| Samut Songkhram | 2023 | 2 | 27.04 | 37.25 | 30.06 | 29.99 |
| Samut Songkhram | 2023 | 3 | 27.99 | 39.23 | 32.36 | 29.89 |
| Samut Songkhram | 2023 | 4 | 29.94 | 41.93 | 36.12 | 32.32 |
| Samut Songkhram | 2023 | 5 | 30.51 | 42.21 | 36.66 | 32.57 |
| Samut Songkhram | 2023 | 6 | 30.18 | 41.76 | 36.01 | 32.05 |
| Samut Songkhram | 2023 | 7 | 29.50 | 40.51 | 34.36 | 31.62 |
| Samut Songkhram | 2023 | 8 | 29.55 | 41.18 | 35.08 | 32.50 |
| Samut Songkhram | 2023 | 9 | 28.68 | 40.13 | 33.45 | 31.72 |
| Songkhla | 2015 | 1 | 26.93 | 36.90 | 29.52 | 29.58 |
| Songkhla | 2015 | 2 | 27.29 | 36.92 | 29.98 | 29.49 |
| Songkhla | 2015 | 3 | 28.68 | 39.19 | 32.75 | 31.80 |
| Songkhla | 2015 | 4 | 29.38 | 41.09 | 34.90 | 33.70 |
| Songkhla | 2015 | 5 | 29.57 | 42.16 | 36.16 | 35.07 |
| Songkhla | 2015 | 6 | 29.01 | 41.28 | 34.82 | 34.22 |
| Songkhla | 2015 | 7 | 28.75 | 40.76 | 34.08 | 33.37 |
| Songkhla | 2015 | 8 | 28.51 | 40.79 | 33.91 | 33.62 |
| Songkhla | 2015 | 9 | 28.42 | 40.48 | 33.54 | 33.29 |
| Songkhla | 2015 | 10 | 28.15 | 40.32 | 33.12 | 33.28 |
| Songkhla | 2015 | 11 | 27.58 | 39.85 | 32.01 | 32.48 |
| Songkhla | 2015 | 12 | 27.78 | 39.31 | 31.97 | 31.72 |
| Songkhla | 2017 | 1 | 26.87 | 38.20 | 29.94 | 30.53 |
| Songkhla | 2017 | 2 | 27.54 | 38.10 | 30.90 | 30.49 |
| Songkhla | 2017 | 3 | 28.32 | 39.65 | 32.80 | 32.15 |
| Songkhla | 2017 | 4 | 28.58 | 41.22 | 34.34 | 33.96 |
| Songkhla | 2017 | 5 | 29.10 | 41.83 | 35.42 | 34.45 |
| Songkhla | 2017 | 6 | 28.93 | 41.25 | 34.75 | 33.95 |
| Songkhla | 2017 | 7 | 29.20 | 41.02 | 34.72 | 33.70 |
| Songkhla | 2017 | 8 | 28.65 | 40.69 | 33.92 | 33.35 |
| Songkhla | 2017 | 9 | 28.38 | 40.68 | 33.69 | 33.35 |
| Songkhla | 2017 | 10 | 28.10 | 39.93 | 32.74 | 32.81 |
| Songkhla | 2017 | 11 | 26.72 | 38.34 | 29.68 | 31.45 |
| Songkhla | 2017 | 12 | 26.67 | 37.60 | 29.42 | 30.43 |
| Songkhla | 2020 | 1 | 27.64 | 37.95 | 30.91 | 30.20 |
| Songkhla | 2020 | 2 | 27.82 | 38.25 | 31.29 | 30.06 |
| Songkhla | 2020 | 3 | 29.12 | 40.33 | 34.07 | 32.66 |
| Songkhla | 2020 | 4 | 29.84 | 42.15 | 36.30 | 34.13 |
| Songkhla | 2020 | 5 | 29.78 | 43.31 | 37.60 | 35.38 |
| Songkhla | 2020 | 6 | 28.87 | 41.95 | 35.40 | 34.39 |
| Songkhla | 2020 | 7 | 28.63 | 41.56 | 34.77 | 34.07 |
| Songkhla | 2020 | 8 | 28.88 | 41.61 | 35.05 | 34.08 |
| Songkhla | 2020 | 9 | 28.49 | 41.21 | 34.28 | 33.63 |
| Songkhla | 2020 | 10 | 27.83 | 40.31 | 32.71 | 33.12 |
| Songkhla | 2020 | 11 | 27.30 | 39.52 | 31.36 | 32.41 |
| Songkhla | 2020 | 12 | 26.54 | 37.89 | 29.23 | 30.93 |
| Songkhla | 2021 | 1 | 26.64 | 37.01 | 29.11 | 29.53 |
| Songkhla | 2021 | 2 | 27.13 | 36.57 | 29.66 | 29.47 |
| Songkhla | 2021 | 3 | 28.43 | 39.59 | 32.91 | 31.97 |
| Songkhla | 2021 | 4 | 28.99 | 41.63 | 35.20 | 34.14 |
| Songkhla | 2021 | 5 | 29.27 | 42.44 | 36.21 | 34.80 |
| Songkhla | 2021 | 6 | 28.96 | 41.37 | 34.91 | 34.06 |
| Songkhla | 2021 | 7 | 29.01 | 41.24 | 34.71 | 33.86 |
| Songkhla | 2021 | 8 | 28.60 | 40.49 | 33.68 | 33.13 |
| Songkhla | 2021 | 9 | 28.44 | 40.59 | 33.65 | 33.43 |
| Songkhla | 2021 | 10 | 28.55 | 40.85 | 33.95 | 33.54 |
| Songkhla | 2021 | 11 | 26.99 | 39.23 | 30.69 | 32.37 |
| Songkhla | 2021 | 12 | 26.93 | 37.93 | 29.93 | 30.50 |
| Songkhla | 2022 | 1 | 27.30 | 37.56 | 30.29 | 30.43 |
| Songkhla | 2022 | 2 | 27.80 | 39.04 | 31.69 | 31.33 |
| Songkhla | 2022 | 3 | 28.63 | 40.66 | 33.93 | 33.45 |
| Songkhla | 2022 | 4 | 28.96 | 41.34 | 34.80 | 34.21 |
| Songkhla | 2022 | 5 | 29.15 | 41.92 | 35.54 | 34.57 |
| Songkhla | 2022 | 6 | 28.88 | 41.12 | 34.57 | 33.97 |
| Songkhla | 2022 | 7 | 29.02 | 41.42 | 34.98 | 34.10 |
| Songkhla | 2022 | 8 | 28.61 | 40.41 | 33.66 | 33.36 |
| Songkhla | 2022 | 9 | 28.37 | 40.04 | 33.11 | 32.76 |
| Songkhla | 2022 | 10 | 27.67 | 39.66 | 32.04 | 32.64 |
| Songkhla | 2022 | 11 | 27.59 | 39.68 | 31.92 | 32.67 |
| Songkhla | 2022 | 12 | 26.35 | 37.24 | 28.68 | 30.04 |
| Songkhla | 2023 | 1 | 26.66 | 37.47 | 29.34 | 30.28 |
| Songkhla | 2023 | 2 | 27.48 | 37.98 | 30.71 | 30.44 |
| Songkhla | 2023 | 3 | 28.03 | 38.50 | 31.71 | 30.98 |
| Songkhla | 2023 | 4 | 29.88 | 42.36 | 36.61 | 35.06 |
| Songkhla | 2023 | 5 | 30.02 | 42.97 | 37.36 | 35.17 |
| Songkhla | 2023 | 6 | 29.58 | 42.06 | 36.05 | 34.43 |
| Songkhla | 2023 | 7 | 29.36 | 41.32 | 35.09 | 33.58 |
| Songkhla | 2023 | 8 | 29.13 | 41.10 | 34.73 | 33.39 |
| Songkhla | 2023 | 9 | 28.50 | 40.67 | 33.76 | 33.30 |
| Songkhla | 2023 | 10 | 28.90 | 41.52 | 34.97 | 34.16 |
| Songkhla | 2023 | 11 | 27.24 | 39.42 | 31.22 | 32.16 |
| Songkhla | 2023 | 12 | 27.79 | 39.85 | 32.29 | 31.76 |
| Surin | 2015 | 1 | 23.50 | 28.44 | 25.52 | 24.16 |
| Surin | 2015 | 2 | 26.39 | 33.85 | 28.51 | 29.06 |
| Surin | 2015 | 3 | 29.73 | 39.95 | 34.00 | 34.18 |
| Surin | 2015 | 4 | 30.37 | 39.76 | 34.29 | 34.07 |
| Surin | 2015 | 5 | 31.29 | 42.59 | 37.33 | 35.96 |
| Surin | 2015 | 6 | 30.24 | 41.81 | 36.05 | 34.97 |
| Surin | 2015 | 7 | 28.80 | 40.24 | 33.40 | 33.57 |
| Surin | 2015 | 8 | 28.59 | 40.21 | 33.45 | 33.68 |
| Surin | 2015 | 9 | 28.19 | 39.71 | 32.58 | 33.05 |
| Surin | 2015 | 10 | 27.07 | 37.37 | 30.02 | 31.34 |
| Surin | 2015 | 11 | 27.72 | 37.82 | 31.08 | 31.72 |
| Surin | 2015 | 12 | 26.21 | 33.79 | 28.14 | 28.51 |
| Surin | 2017 | 1 | 26.20 | 34.05 | 27.97 | 28.56 |
| Surin | 2017 | 2 | 25.94 | 32.80 | 27.56 | 27.88 |
| Surin | 2017 | 3 | 29.22 | 39.30 | 33.43 | 33.34 |
| Surin | 2017 | 4 | 30.13 | 41.19 | 35.48 | 34.66 |
| Surin | 2017 | 5 | 29.62 | 42.04 | 35.97 | 35.24 |
| Surin | 2017 | 6 | 29.61 | 42.04 | 36.04 | 34.99 |
| Surin | 2017 | 7 | 28.41 | 40.49 | 33.52 | 33.44 |
| Surin | 2017 | 8 | 29.03 | 41.25 | 34.75 | 34.29 |
| Surin | 2017 | 9 | 29.02 | 41.51 | 35.03 | 34.64 |
| Surin | 2017 | 10 | 28.00 | 39.24 | 32.32 | 32.59 |
| Surin | 2017 | 11 | 26.71 | 35.47 | 29.15 | 29.31 |
| Surin | 2017 | 12 | 24.34 | 30.66 | 26.58 | 25.28 |
| Surin | 2020 | 1 | 26.08 | 33.17 | 27.52 | 28.80 |
| Surin | 2020 | 2 | 26.49 | 32.81 | 27.83 | 28.45 |
| Surin | 2020 | 3 | 30.65 | 40.27 | 34.69 | 34.90 |
| Surin | 2020 | 4 | 30.24 | 40.65 | 35.00 | 34.68 |
| Surin | 2020 | 5 | 31.72 | 43.74 | 38.80 | 37.19 |
| Surin | 2020 | 6 | 29.60 | 41.46 | 35.32 | 35.11 |
| Surin | 2020 | 7 | 29.45 | 41.05 | 34.91 | 34.85 |
| Surin | 2020 | 8 | 28.36 | 39.97 | 33.02 | 33.58 |
| Surin | 2020 | 9 | 28.49 | 40.36 | 33.45 | 34.14 |
| Surin | 2020 | 10 | 25.16 | 34.69 | 26.36 | 29.22 |
| Surin | 2020 | 11 | 25.65 | 33.41 | 27.22 | 28.24 |
| Surin | 2020 | 12 | 23.65 | 29.63 | 25.47 | 25.20 |
| Surin | 2021 | 1 | 22.94 | 27.86 | 25.17 | 24.30 |
| Surin | 2021 | 2 | 25.83 | 32.29 | 27.14 | 28.35 |
| Surin | 2021 | 3 | 29.97 | 39.61 | 33.92 | 34.25 |
| Surin | 2021 | 4 | 30.33 | 41.83 | 36.14 | 35.73 |
| Surin | 2021 | 5 | 30.34 | 42.48 | 36.85 | 36.09 |
| Surin | 2021 | 6 | 29.69 | 41.74 | 35.66 | 35.12 |
| Surin | 2021 | 7 | 28.97 | 40.55 | 33.99 | 34.12 |
| Surin | 2021 | 8 | 29.26 | 41.18 | 34.86 | 34.86 |
| Surin | 2021 | 9 | 27.81 | 39.74 | 32.20 | 33.68 |
| Surin | 2021 | 10 | 27.23 | 38.11 | 30.65 | 32.53 |
| Surin | 2021 | 11 | 26.10 | 34.69 | 28.26 | 29.50 |
| Surin | 2021 | 12 | 24.02 | 30.05 | 25.36 | 25.81 |
| Surin | 2022 | 1 | 25.73 | 32.80 | 27.02 | 28.76 |
| Surin | 2022 | 2 | 26.11 | 34.11 | 28.48 | 29.31 |
| Surin | 2022 | 3 | 29.46 | 40.15 | 34.15 | 34.45 |
| Surin | 2022 | 4 | 28.46 | 38.62 | 32.82 | 32.99 |
| Surin | 2022 | 5 | 28.38 | 40.05 | 33.44 | 33.77 |
| Surin | 2022 | 6 | 29.93 | 41.81 | 35.97 | 35.41 |
| Surin | 2022 | 7 | 29.05 | 41.34 | 34.88 | 34.82 |
| Surin | 2022 | 8 | 28.58 | 40.63 | 33.82 | 34.03 |
| Surin | 2022 | 9 | 27.80 | 39.87 | 32.30 | 33.45 |
| Surin | 2022 | 10 | 26.43 | 35.71 | 28.74 | 30.13 |
| Surin | 2022 | 11 | 27.44 | 37.19 | 30.65 | 31.59 |
| Surin | 2022 | 12 | 23.96 | 30.16 | 25.75 | 25.67 |
| Surin | 2023 | 1 | 23.29 | 28.79 | 24.95 | 24.43 |
| Surin | 2023 | 2 | 27.27 | 35.15 | 29.71 | 30.17 |
| Surin | 2023 | 3 | 29.00 | 37.41 | 31.98 | 32.58 |
| Surin | 2023 | 4 | 31.99 | 43.03 | 37.95 | 36.97 |
| Surin | 2023 | 5 | 31.04 | 43.39 | 38.09 | 36.77 |
| Surin | 2023 | 6 | 29.93 | 42.47 | 36.64 | 35.48 |
| Surin | 2023 | 7 | 29.52 | 41.81 | 35.68 | 35.12 |
| Surin | 2023 | 8 | 29.36 | 41.88 | 35.65 | 35.03 |
| Surin | 2023 | 9 | 28.37 | 40.88 | 33.81 | 34.22 |
| Surin | 2023 | 10 | 28.47 | 40.36 | 33.58 | 33.90 |
| Surin | 2023 | 11 | 27.09 | 36.48 | 30.38 | 30.36 |
| Surin | 2023 | 12 | 26.10 | 33.81 | 28.60 | 28.50 |
| Uttaradit | 2015 | 1 | 23.97 | 29.16 | 25.16 | 26.33 |
| Uttaradit | 2015 | 2 | 26.93 | 33.65 | 28.37 | 30.06 |
| Uttaradit | 2015 | 3 | 30.40 | 40.06 | 34.39 | 35.18 |
| Uttaradit | 2015 | 4 | 31.30 | 40.13 | 34.96 | 35.53 |
| Uttaradit | 2015 | 5 | 32.35 | 43.95 | 39.18 | 38.10 |
| Uttaradit | 2015 | 6 | 31.25 | 42.42 | 37.08 | 36.92 |
| Uttaradit | 2015 | 7 | 29.71 | 41.56 | 35.37 | 35.88 |
| Uttaradit | 2015 | 8 | 29.76 | 42.26 | 36.33 | 36.24 |
| Uttaradit | 2015 | 9 | 29.97 | 42.56 | 36.82 | 36.69 |
| Uttaradit | 2015 | 10 | 29.19 | 40.81 | 34.65 | 35.13 |
| Uttaradit | 2015 | 11 | 29.46 | 39.46 | 33.52 | 34.48 |
| Uttaradit | 2015 | 12 | 26.62 | 34.57 | 28.49 | 30.25 |
| Uttaradit | 2017 | 1 | 26.45 | 34.35 | 28.02 | 30.10 |
| Uttaradit | 2017 | 2 | 27.31 | 33.72 | 28.71 | 30.01 |
| Uttaradit | 2017 | 3 | 30.40 | 39.37 | 33.83 | 34.50 |
| Uttaradit | 2017 | 4 | 31.17 | 42.21 | 36.94 | 36.69 |
| Uttaradit | 2017 | 5 | 31.24 | 44.45 | 39.65 | 37.95 |
| Uttaradit | 2017 | 6 | 30.52 | 43.64 | 38.38 | 37.28 |
| Uttaradit | 2017 | 7 | 29.18 | 42.34 | 35.99 | 36.04 |
| Uttaradit | 2017 | 8 | 29.77 | 42.92 | 37.08 | 36.55 |
| Uttaradit | 2017 | 9 | 30.05 | 43.69 | 38.22 | 37.11 |
| Uttaradit | 2017 | 10 | 29.15 | 41.59 | 35.36 | 35.63 |
| Uttaradit | 2017 | 11 | 28.44 | 38.46 | 32.38 | 33.12 |
| Uttaradit | 2017 | 12 | 25.74 | 33.27 | 27.96 | 29.09 |
| Uttaradit | 2020 | 1 | 27.41 | 34.41 | 29.26 | 30.54 |
| Uttaradit | 2020 | 2 | 28.22 | 34.39 | 29.30 | 30.58 |
| Uttaradit | 2020 | 3 | 31.35 | 40.21 | 34.85 | 35.43 |
| Uttaradit | 2020 | 4 | 32.63 | 42.58 | 37.47 | 37.27 |
| Uttaradit | 2020 | 5 | 33.38 | 45.82 | 41.80 | 39.43 |
| Uttaradit | 2020 | 6 | 31.73 | 44.20 | 39.26 | 37.96 |
| Uttaradit | 2020 | 7 | 31.41 | 44.14 | 39.28 | 37.82 |
| Uttaradit | 2020 | 8 | 29.48 | 42.43 | 36.24 | 35.78 |
| Uttaradit | 2020 | 9 | 30.09 | 43.50 | 38.00 | 37.04 |
| Uttaradit | 2020 | 10 | 28.56 | 40.21 | 33.55 | 34.45 |
| Uttaradit | 2020 | 11 | 28.51 | 37.80 | 31.74 | 32.72 |
| Uttaradit | 2020 | 12 | 26.09 | 32.59 | 27.66 | 28.75 |
| Uttaradit | 2021 | 1 | 24.79 | 30.00 | 26.34 | 26.77 |
| Uttaradit | 2021 | 2 | 27.14 | 33.34 | 28.28 | 30.01 |
| Uttaradit | 2021 | 3 | 31.14 | 40.73 | 35.42 | 36.06 |
| Uttaradit | 2021 | 4 | 30.56 | 42.35 | 36.79 | 36.66 |
| Uttaradit | 2021 | 5 | 32.50 | 44.78 | 40.29 | 38.95 |
| Uttaradit | 2021 | 6 | 30.79 | 42.89 | 37.45 | 37.06 |
| Uttaradit | 2021 | 7 | 30.16 | 43.29 | 37.69 | 37.16 |
| Uttaradit | 2021 | 8 | 30.16 | 43.00 | 37.38 | 36.96 |
| Uttaradit | 2021 | 9 | 29.41 | 42.48 | 36.43 | 36.39 |
| Uttaradit | 2021 | 10 | 29.07 | 41.59 | 35.24 | 35.71 |
| Uttaradit | 2021 | 11 | 28.71 | 39.27 | 32.99 | 34.13 |
| Uttaradit | 2021 | 12 | 25.31 | 31.62 | 26.59 | 28.12 |
| Uttaradit | 2022 | 1 | 26.16 | 33.14 | 27.38 | 29.58 |
| Uttaradit | 2022 | 2 | 26.85 | 35.31 | 28.89 | 30.92 |
| Uttaradit | 2022 | 3 | 30.77 | 41.64 | 36.19 | 36.29 |
| Uttaradit | 2022 | 4 | 31.30 | 41.58 | 36.42 | 36.30 |
| Uttaradit | 2022 | 5 | 30.48 | 42.98 | 37.62 | 36.51 |
| Uttaradit | 2022 | 6 | 31.02 | 43.33 | 38.17 | 37.20 |
| Uttaradit | 2022 | 7 | 30.66 | 43.85 | 38.67 | 37.49 |
| Uttaradit | 2022 | 8 | 29.55 | 42.64 | 36.61 | 36.23 |
| Uttaradit | 2022 | 9 | 29.54 | 42.76 | 36.77 | 36.28 |
| Uttaradit | 2022 | 10 | 28.75 | 40.00 | 33.61 | 34.59 |
| Uttaradit | 2022 | 11 | 28.57 | 38.85 | 32.64 | 33.66 |
| Uttaradit | 2022 | 12 | 26.29 | 33.93 | 28.38 | 29.76 |
| Uttaradit | 2023 | 1 | 25.34 | 31.03 | 26.63 | 27.45 |
| Uttaradit | 2023 | 2 | 27.79 | 35.02 | 29.69 | 31.10 |
| Uttaradit | 2023 | 3 | 29.86 | 38.47 | 32.91 | 34.04 |
| Uttaradit | 2023 | 4 | 33.19 | 44.23 | 39.71 | 38.83 |
| Uttaradit | 2023 | 5 | 32.78 | 45.16 | 40.79 | 39.06 |
| Uttaradit | 2023 | 6 | 32.01 | 44.55 | 39.94 | 37.96 |
| Uttaradit | 2023 | 7 | 31.31 | 44.58 | 39.85 | 37.89 |
| Uttaradit | 2023 | 8 | 30.55 | 43.38 | 37.95 | 36.85 |
| Uttaradit | 2023 | 9 | 29.65 | 43.30 | 37.42 | 36.79 |
| Uttaradit | 2023 | 10 | 29.72 | 42.96 | 37.12 | 36.55 |
| Uttaradit | 2023 | 11 | 28.69 | 38.96 | 33.13 | 33.97 |
| Uttaradit | 2023 | 12 | 27.78 | 36.33 | 30.88 | 31.97 |

### S5. Directed Acyclic Graphs

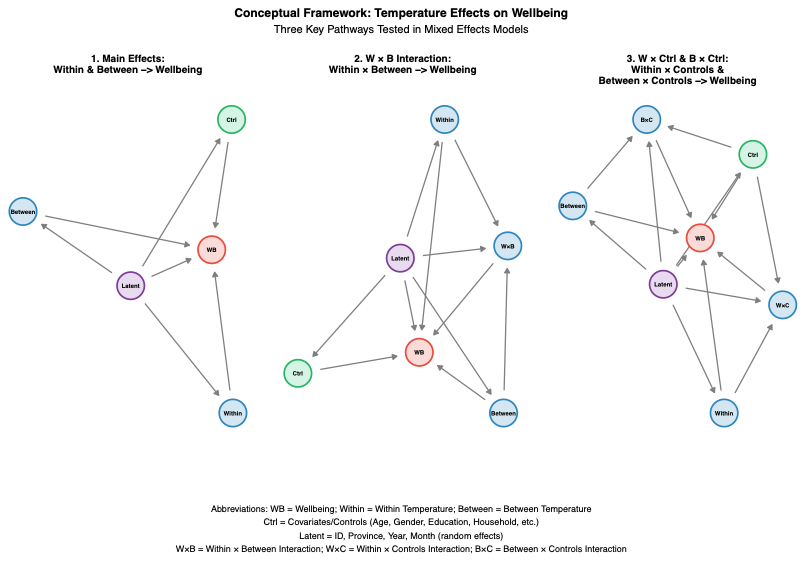

### S6. Three way interactions between vulnerabilities, within and between variables

| **Heat metric** | **Outcome** | **Vulnerability** | **Term** | **Coefficient** | **95% CI** | **Significance** |
| --- | --- | --- | --- | --- | --- | --- |
| Temperature | Psychological Distress | Economic | Within (main) | 0.60660231 | [-0.7284; 1.9416] |  |
|  | Psychological Distress | Economic | Between (main) | 0.84985748 | [-0.4917; 2.1914] |  |
|  | Psychological Distress | Economic | Vulnerability (main) | 0.00383216 | [-4.9849; 4.9925] |  |
|  | Psychological Distress | Economic | Within × Vulnerability | 0.02749346 | [-0.153; 0.208] |  |
|  | Psychological Distress | Economic | Within × Between | -0.0248546 | [-0.0721; 0.0224] |  |
|  | Psychological Distress | Economic | Vulnerability × Between | -0.0273731 | [-0.2071; 0.1523] |  |
|  | Psychological Distress | Economic | Three-way interaction | -0.0003792 | [-0.0068; 0.006] |  |
|  | Psychological Distress | Education | Within (main) | 0.12467215 | [-0.8114; 1.0608] |  |
|  | Psychological Distress | Education | Between (main) | -0.1313332 | [-1.0961; 0.8334] |  |
|  | Psychological Distress | Education | Vulnerability (main) | -33.215056 | [-83.6319; 17.2018] |  |
|  | Psychological Distress | Education | Within × Vulnerability | 0.95488763 | [-0.8479; 2.7577] |  |
|  | Psychological Distress | Education | Within × Between | -0.0024175 | [-0.0354; 0.0306] |  |
|  | Psychological Distress | Education | Vulnerability × Between | 1.31457375 | [-0.4792; 3.1083] |  |
|  | Psychological Distress | Education | Three-way interaction | -0.0376478 | [-0.1008; 0.0255] |  |
|  | Psychological Distress | Work | Within (main) | 1.19244511 | [0.5151; 1.8698] | *** |
|  | Psychological Distress | Work | Between (main) | 1.04820968 | [0.3277; 1.7688] | ** |
|  | Psychological Distress | Work | Vulnerability (main) | 21.5681278 | [-0.7421; 43.8784] |  |
|  | Psychological Distress | Work | Within × Vulnerability | -0.791726 | [-1.5905; 0.0071] |  |
|  | Psychological Distress | Work | Within × Between | -0.0409721 | [-0.065; -0.0169] | *** |
|  | Psychological Distress | Work | Vulnerability × Between | -0.7224414 | [-1.5179; 0.0731] |  |
|  | Psychological Distress | Work | Three-way interaction | 0.02623007 | [-0.0019; 0.0544] |  |
|  | Psychological Distress | ADL | Within (main) | 0.92927496 | [0.3123; 1.5463] | ** |
|  | Psychological Distress | ADL | Between (main) | 0.82912968 | [0.1612; 1.497] | * |
|  | Psychological Distress | ADL | Vulnerability (main) | -6.9958973 | [-55.9315; 41.9397] |  |
|  | Psychological Distress | ADL | Within × Vulnerability | 0.48239585 | [-1.2833; 2.2481] |  |
|  | Psychological Distress | ADL | Within × Between | -0.032584 | [-0.0545; -0.0107] | ** |
|  | Psychological Distress | ADL | Vulnerability × Between | 0.13835256 | [-1.5948; 1.8715] |  |
|  | Psychological Distress | ADL | Three-way interaction | -0.010823 | [-0.0726; 0.051] |  |
|  | Mental Health | Economic | Within (main) | -0.7622951 | [-1.3422; -0.1824] | ** |
|  | Mental Health | Economic | Between (main) | -0.8062794 | [-1.3922; -0.2203] | ** |
|  | Mental Health | Economic | Vulnerability (main) | -4.0601705 | [-6.2347; -1.8856] | *** |
|  | Mental Health | Economic | Within × Vulnerability | 0.16631162 | [0.0877; 0.2449] | *** |
|  | Mental Health | Economic | Within × Between | 0.03390405 | [0.0134; 0.0544] | ** |
|  | Mental Health | Economic | Vulnerability × Between | 0.15232145 | [0.0739; 0.2307] | *** |
|  | Mental Health | Economic | Three-way interaction | -0.0064536 | [-0.0092; -0.0037] | *** |
|  | Mental Health | Education | Within (main) | 0.31240548 | [-0.0937; 0.7185] |  |
|  | Mental Health | Education | Between (main) | 0.17274822 | [-0.2519; 0.5974] |  |
|  | Mental Health | Education | Vulnerability (main) | 1.8341383 | [-19.3808; 23.0491] |  |
|  | Mental Health | Education | Within × Vulnerability | -0.0011024 | [-0.7592; 0.757] |  |
|  | Mental Health | Education | Within × Between | -0.0076804 | [-0.0221; 0.0067] |  |
|  | Mental Health | Education | Vulnerability × Between | -0.1334699 | [-0.8906; 0.6237] |  |
|  | Mental Health | Education | Three-way interaction | 0.00282697 | [-0.0238; 0.0294] |  |
|  | Mental Health | Work | Within (main) | 0.50104441 | [0.2034; 0.7987] | *** |
|  | Mental Health | Work | Between (main) | 0.32809444 | [0.009; 0.6472] | * |
|  | Mental Health | Work | Vulnerability (main) | 1.61494706 | [-7.8867; 11.1166] |  |
|  | Mental Health | Work | Within × Vulnerability | -0.0859005 | [-0.4262; 0.2544] |  |
|  | Mental Health | Work | Within × Between | -0.0135808 | [-0.0242; -0.003] | * |
|  | Mental Health | Work | Vulnerability × Between | -0.0143155 | [-0.3532; 0.3246] |  |
|  | Mental Health | Work | Three-way interaction | 0.0012759 | [-0.0107; 0.0133] |  |
|  | Mental Health | ADL | Within (main) | 0.5239805 | [0.2514; 0.7966] | *** |
|  | Mental Health | ADL | Between (main) | 0.38076811 | [0.083; 0.6785] | * |
|  | Mental Health | ADL | Vulnerability (main) | 24.4541511 | [2.9532; 45.9551] | * |
|  | Mental Health | ADL | Within × Vulnerability | -0.806692 | [-1.5819; -0.0315] | * |
|  | Mental Health | ADL | Within × Between | -0.015099 | [-0.0248; -0.0054] | ** |
|  | Mental Health | ADL | Vulnerability × Between | -0.8786473 | [-1.6416; -0.1157] | * |
|  | Mental Health | ADL | Three-way interaction | 0.02978211 | [0.0026; 0.057] | * |
|  | Physical Health | Economic | Within (main) | -0.2362201 | [-0.8087; 0.3362] |  |
|  | Physical Health | Economic | Between (main) | -0.259512 | [-0.8303; 0.3113] |  |
|  | Physical Health | Economic | Vulnerability (main) | -1.1729391 | [-3.3735; 1.0276] |  |
|  | Physical Health | Economic | Within × Vulnerability | 0.05152335 | [-0.0282; 0.1312] |  |
|  | Physical Health | Economic | Within × Between | 0.0141521 | [-0.0061; 0.0344] |  |
|  | Physical Health | Economic | Vulnerability × Between | 0.05253573 | [-0.0267; 0.1317] |  |
|  | Physical Health | Economic | Three-way interaction | -0.0025131 | [-0.0053; 3e-04] |  |
|  | Physical Health | Education | Within (main) | 0.21304065 | [-0.1862; 0.6123] |  |
|  | Physical Health | Education | Between (main) | 0.24809141 | [-0.1591; 0.6553] |  |
|  | Physical Health | Education | Vulnerability (main) | -8.2060941 | [-30.4084; 13.9962] |  |
|  | Physical Health | Education | Within × Vulnerability | 0.357762 | [-0.4349; 1.1504] |  |
|  | Physical Health | Education | Within × Between | -0.0069301 | [-0.021; 0.0072] |  |
|  | Physical Health | Education | Vulnerability × Between | 0.2099037 | [-0.5806; 1.0004] |  |
|  | Physical Health | Education | Three-way interaction | -0.0095198 | [-0.0373; 0.0183] |  |
|  | Physical Health | Work | Within (main) | 0.30629337 | [0.0276; 0.585] | * |
|  | Physical Health | Work | Between (main) | 0.24528558 | [-0.046; 0.5366] |  |
|  | Physical Health | Work | Vulnerability (main) | 3.40557918 | [-6.4362; 13.2474] |  |
|  | Physical Health | Work | Within × Vulnerability | -0.189624 | [-0.5418; 0.1625] |  |
|  | Physical Health | Work | Within × Between | -0.0087515 | [-0.0187; 0.0012] |  |
|  | Physical Health | Work | Vulnerability × Between | -0.0828309 | [-0.4336; 0.2679] |  |
|  | Physical Health | Work | Three-way interaction | 0.0049166 | [-0.0075; 0.0173] |  |
|  | Physical Health | ADL | Within (main) | 0.3290075 | [0.0796; 0.5784] | ** |
|  | Physical Health | ADL | Between (main) | 0.30978843 | [0.0444; 0.5752] | * |
|  | Physical Health | ADL | Vulnerability (main) | 61.4546456 | [39.5523; 83.357] | *** |
|  | Physical Health | ADL | Within × Vulnerability | -2.1242914 | [-2.9148; -1.3338] | *** |
|  | Physical Health | ADL | Within × Between | -0.0102372 | [-0.0192; -0.0013] | * |
|  | Physical Health | ADL | Vulnerability × Between | -2.1657462 | [-2.9429; -1.3886] | *** |
|  | Physical Health | ADL | Three-way interaction | 0.07574584 | [0.048; 0.1035] | *** |
|  | Life Quality | Economic | Within (main) | 1.3218918 | [0.7883; 1.8555] | *** |
|  | Life Quality | Economic | Between (main) | 0.73104669 | [0.1843; 1.2777] | ** |
|  | Life Quality | Economic | Vulnerability (main) | 4.36249309 | [2.3594; 6.3656] | *** |
|  | Life Quality | Economic | Within × Vulnerability | -0.1891326 | [-0.2615; -0.1167] | *** |
|  | Life Quality | Economic | Within × Between | -0.0335835 | [-0.0525; -0.0147] | *** |
|  | Life Quality | Economic | Vulnerability × Between | -0.1294835 | [-0.2017; -0.0572] | *** |
|  | Life Quality | Economic | Three-way interaction | 0.00538612 | [0.0028; 0.008] | *** |
|  | Life Quality | Education | Within (main) | 0.14144568 | [0.0946; 0.1883] | *** |
|  | Life Quality | Education | Between (main) | -0.0939645 | [-0.231; 0.0431] |  |
|  | Life Quality | Education | Vulnerability (main) | -1.2856919 | [-3.4002; 0.8288] |  |
|  | Life Quality | Education | Within × Vulnerability | -0.0183774 | [-0.0947; 0.058] |  |
|  | Life Quality | Education | Vulnerability × Between | 0.07298633 | [-0.0388; 0.1848] |  |
|  | Life Quality | Work | Within (main) | 0.15134524 | [-0.1228; 0.4255] |  |
|  | Life Quality | Work | Between (main) | -0.0663533 | [-0.3756; 0.2429] |  |
|  | Life Quality | Work | Vulnerability (main) | 2.30336425 | [-6.4321; 11.0388] |  |
|  | Life Quality | Work | Within × Vulnerability | -0.1097428 | [-0.4224; 0.2029] |  |
|  | Life Quality | Work | Within × Between | -0.0003504 | [-0.0101; 0.0094] |  |
|  | Life Quality | Work | Vulnerability × Between | -0.0623493 | [-0.3741; 0.2494] |  |
|  | Life Quality | Work | Three-way interaction | 0.0030906 | [-0.0079; 0.0141] |  |
|  | Life Quality | ADL | Within (main) | 0.13694862 | [-0.1148; 0.3887] |  |
|  | Life Quality | ADL | Between (main) | -0.0617687 | [-0.3526; 0.229] |  |
|  | Life Quality | ADL | Vulnerability (main) | 17.9211239 | [-1.226; 37.0683] |  |
|  | Life Quality | ADL | Within × Vulnerability | -0.5867149 | [-1.2772; 0.1037] |  |
|  | Life Quality | ADL | Within × Between | -0.0001143 | [-0.0091; 0.0088] |  |
|  | Life Quality | ADL | Vulnerability × Between | -0.604085 | [-1.2836; 0.0754] |  |
|  | Life Quality | ADL | Three-way interaction | 0.02031824 | [-0.0039; 0.0445] |  |
| Humidex | Psychological Distress | Economic | Within (main) | -0.0093609 | [-0.5866; 0.5679] |  |
|  | Psychological Distress | Economic | Between (main) | 0.00350488 | [-0.5723; 0.5793] |  |
|  | Psychological Distress | Economic | Vulnerability (main) | -1.7480278 | [-4.6836; 1.1876] |  |
|  | Psychological Distress | Economic | Within × Vulnerability | 0.05781718 | [-0.0222; 0.1378] |  |
|  | Psychological Distress | Economic | Within × Between | 0.00087353 | [-0.0141; 0.0159] |  |
|  | Psychological Distress | Economic | Vulnerability × Between | 0.03634822 | [-0.0419; 0.1146] |  |
|  | Psychological Distress | Economic | Three-way interaction | -0.0014547 | [-0.0035; 6e-04] |  |
|  | Psychological Distress | Education | Within (main) | 0.22603732 | [-0.1862; 0.6382] |  |
|  | Psychological Distress | Education | Between (main) | -0.0041133 | [-0.43; 0.4218] |  |
|  | Psychological Distress | Education | Vulnerability (main) | -10.512156 | [-39.4802; 18.4559] |  |
|  | Psychological Distress | Education | Within × Vulnerability | 0.16576357 | [-0.6093; 0.9408] |  |
|  | Psychological Distress | Education | Within × Between | -0.004061 | [-0.0148; 0.0067] |  |
|  | Psychological Distress | Education | Vulnerability × Between | 0.36916166 | [-0.4031; 1.1415] |  |
|  | Psychological Distress | Education | Three-way interaction | -0.0062556 | [-0.0264; 0.0139] |  |
|  | Psychological Distress | Work | Within (main) | 0.45089903 | [0.166; 0.7358] | ** |
|  | Psychological Distress | Work | Between (main) | 0.33464648 | [0.0245; 0.6447] | * |
|  | Psychological Distress | Work | Vulnerability (main) | 8.91329122 | [-4.533; 22.3596] |  |
|  | Psychological Distress | Work | Within × Vulnerability | -0.2142347 | [-0.5753; 0.1469] |  |
|  | Psychological Distress | Work | Within × Between | -0.0108168 | [-0.0183; -0.0033] | ** |
|  | Psychological Distress | Work | Vulnerability × Between | -0.2389425 | [-0.595; 0.1171] |  |
|  | Psychological Distress | Work | Three-way interaction | 0.00558103 | [-0.0038; 0.015] |  |
|  | Psychological Distress | ADL | Within (main) | 0.42078619 | [0.1607; 0.6809] | ** |
|  | Psychological Distress | ADL | Between (main) | 0.29041691 | [0; 0.5808] | * |
|  | Psychological Distress | ADL | Vulnerability (main) | 15.3623126 | [-15.1676; 45.8923] |  |
|  | Psychological Distress | ADL | Within × Vulnerability | -0.4863783 | [-1.307; 0.3343] |  |
|  | Psychological Distress | ADL | Within × Between | -0.0100639 | [-0.0169; -0.0032] | ** |
|  | Psychological Distress | ADL | Vulnerability × Between | -0.3729006 | [-1.1729; 0.4271] |  |
|  | Psychological Distress | ADL | Three-way interaction | 0.01309279 | [-0.0081; 0.0343] |  |
|  | Mental Health | Economic | Within (main) | -0.7407401 | [-0.9928; -0.4887] | *** |
|  | Mental Health | Economic | Between (main) | -0.6940838 | [-0.9486; -0.4396] | *** |
|  | Mental Health | Economic | Vulnerability (main) | -3.9483141 | [-5.2314; -2.6652] | *** |
|  | Mental Health | Economic | Within × Vulnerability | 0.12271536 | [0.0877; 0.1577] | *** |
|  | Mental Health | Economic | Within × Between | 0.02154185 | [0.015; 0.0281] | *** |
|  | Mental Health | Economic | Vulnerability × Between | 0.10298573 | [0.0688; 0.1372] | *** |
|  | Mental Health | Economic | Three-way interaction | -0.0033273 | [-0.0042; -0.0024] | *** |
|  | Mental Health | Education | Within (main) | 0.06185957 | [-0.1171; 0.2408] |  |
|  | Mental Health | Education | Between (main) | -0.024851 | [-0.2132; 0.1635] |  |
|  | Mental Health | Education | Vulnerability (main) | 5.58366625 | [-6.7324; 17.8998] |  |
|  | Mental Health | Education | Within × Vulnerability | -0.1164734 | [-0.4468; 0.2139] |  |
|  | Mental Health | Education | Within × Between | -0.0001919 | [-0.0049; 0.0045] |  |
|  | Mental Health | Education | Vulnerability × Between | -0.1657388 | [-0.4944; 0.1629] |  |
|  | Mental Health | Education | Three-way interaction | 0.00373405 | [-0.0049; 0.0123] |  |
|  | Mental Health | Work | Within (main) | 0.1066631 | [-0.0202; 0.2335] |  |
|  | Mental Health | Work | Between (main) | 0.01267407 | [-0.1298; 0.1551] |  |
|  | Mental Health | Work | Vulnerability (main) | 2.73837149 | [-3.0217; 8.4984] |  |
|  | Mental Health | Work | Within × Vulnerability | -0.0817487 | [-0.2365; 0.073] |  |
|  | Mental Health | Work | Within × Between | -0.0011778 | [-0.0045; 0.0022] |  |
|  | Mental Health | Work | Vulnerability × Between | -0.0569448 | [-0.2094; 0.0955] |  |
|  | Mental Health | Work | Three-way interaction | 0.00161466 | [-0.0024; 0.0056] |  |
|  | Mental Health | ADL | Within (main) | 0.10711504 | [-0.0098; 0.224] |  |
|  | Mental Health | ADL | Between (main) | 0.01922113 | [-0.1154; 0.1539] |  |
|  | Mental Health | ADL | Vulnerability (main) | 17.5294092 | [4.0528; 31.006] | * |
|  | Mental Health | ADL | Within × Vulnerability | -0.4657959 | [-0.8282; -0.1034] | * |
|  | Mental Health | ADL | Within × Between | -0.0013516 | [-0.0044; 0.0017] |  |
|  | Mental Health | ADL | Vulnerability × Between | -0.4393902 | [-0.7922; -0.0866] | * |
|  | Mental Health | ADL | Three-way interaction | 0.01206748 | [0.0027; 0.0214] | * |
|  | Physical Health | Economic | Within (main) | -0.3500177 | [-0.6018; -0.0983] | ** |
|  | Physical Health | Economic | Between (main) | -0.3015534 | [-0.5507; -0.0524] | * |
|  | Physical Health | Economic | Vulnerability (main) | -2.0293782 | [-3.3327; -0.726] | ** |
|  | Physical Health | Economic | Within × Vulnerability | 0.06208953 | [0.0266; 0.0976] | *** |
|  | Physical Health | Economic | Within × Between | 0.01088681 | [0.0043; 0.0175] | ** |
|  | Physical Health | Economic | Vulnerability × Between | 0.05489232 | [0.0201; 0.0897] | ** |
|  | Physical Health | Economic | Three-way interaction | -0.0018213 | [-0.0027; -9e-04] | *** |
|  | Physical Health | Education | Within (main) | 0.14961589 | [-0.0276; 0.3269] |  |
|  | Physical Health | Education | Between (main) | 0.15308073 | [-0.0267; 0.3328] |  |
|  | Physical Health | Education | Vulnerability (main) | 2.94569111 | [-9.8638; 15.7552] |  |
|  | Physical Health | Education | Within × Vulnerability | -0.0517202 | [-0.3945; 0.291] |  |
|  | Physical Health | Education | Within × Between | -0.0035096 | [-0.0081; 0.0011] |  |
|  | Physical Health | Education | Vulnerability × Between | -0.0987634 | [-0.4403; 0.2428] |  |
|  | Physical Health | Education | Three-way interaction | 0.00205054 | [-0.0069; 0.011] |  |
|  | Physical Health | Work | Within (main) | 0.1219623 | [0.0029; 0.241] | * |
|  | Physical Health | Work | Between (main) | 0.11124694 | [-0.0145; 0.237] |  |
|  | Physical Health | Work | Vulnerability (main) | 3.27551501 | [-2.6741; 9.2251] |  |
|  | Physical Health | Work | Within × Vulnerability | -0.1165546 | [-0.2763; 0.0432] |  |
|  | Physical Health | Work | Within × Between | -0.0026465 | [-0.0058; 5e-04] |  |
|  | Physical Health | Work | Vulnerability × Between | -0.0825526 | [-0.2399; 0.0748] |  |
|  | Physical Health | Work | Three-way interaction | 0.00270512 | [-0.0014; 0.0069] |  |
|  | Physical Health | ADL | Within (main) | 0.11942221 | [0.0126; 0.2262] | * |
|  | Physical Health | ADL | Between (main) | 0.12196238 | [0.0064; 0.2375] | * |
|  | Physical Health | ADL | Vulnerability (main) | 36.8798665 | [23.2948; 50.4649] | *** |
|  | Physical Health | ADL | Within × Vulnerability | -0.9328075 | [-1.299; -0.5666] | *** |
|  | Physical Health | ADL | Within × Between | -0.0027003 | [-0.0055; 1e-04] |  |
|  | Physical Health | ADL | Vulnerability × Between | -0.9442246 | [-1.2997; -0.5887] | *** |
|  | Physical Health | ADL | Three-way interaction | 0.02433947 | [0.0149; 0.0338] | *** |
|  | Life Quality | Economic | Within (main) | 0.24653122 | [0.0168; 0.4763] | * |
|  | Life Quality | Economic | Between (main) | 0.07931525 | [-0.1595; 0.3182] |  |
|  | Life Quality | Economic | Vulnerability (main) | 1.18303471 | [0.0176; 2.3484] | * |
|  | Life Quality | Economic | Within × Vulnerability | -0.0405611 | [-0.0723; -0.0088] | * |
|  | Life Quality | Economic | Within × Between | -0.0030936 | [-0.0091; 0.0029] |  |
|  | Life Quality | Economic | Vulnerability × Between | -0.0285835 | [-0.0597; 0.0025] |  |
|  | Life Quality | Economic | Three-way interaction | 0.00077543 | [-1e-04; 0.0016] |  |
|  | Life Quality | Education | Within (main) | -0.0105461 | [-0.1762; 0.1551] |  |
|  | Life Quality | Education | Between (main) | -0.1068092 | [-0.2896; 0.076] |  |
|  | Life Quality | Education | Vulnerability (main) | 9.11312907 | [-2.2051; 20.4313] |  |
|  | Life Quality | Education | Within × Vulnerability | -0.2763633 | [-0.5801; 0.0274] |  |
|  | Life Quality | Education | Within × Between | 0.00176961 | [-0.0025; 0.0061] |  |
|  | Life Quality | Education | Vulnerability × Between | -0.2161953 | [-0.5182; 0.0859] |  |
|  | Life Quality | Education | Three-way interaction | 0.00676776 | [-0.0011; 0.0147] |  |
|  | Life Quality | Work | Within (main) | 0.00421887 | [-0.1125; 0.1209] |  |
|  | Life Quality | Work | Between (main) | -0.0857203 | [-0.2282; 0.0568] |  |
|  | Life Quality | Work | Vulnerability (main) | 0.68942001 | [-4.608; 5.9868] |  |
|  | Life Quality | Work | Within × Vulnerability | -0.0265071 | [-0.1689; 0.1159] |  |
|  | Life Quality | Work | Within × Between | 0.00139478 | [-0.0017; 0.0045] |  |
|  | Life Quality | Work | Vulnerability × Between | -0.0140307 | [-0.1542; 0.1262] |  |
|  | Life Quality | Work | Three-way interaction | 0.0005292 | [-0.0032; 0.0042] |  |
|  | Life Quality | ADL | Within (main) | 0.00911055 | [-0.099; 0.1172] |  |
|  | Life Quality | ADL | Between (main) | -0.0792777 | [-0.2159; 0.0573] |  |
|  | Life Quality | ADL | Vulnerability (main) | 13.4663013 | [1.4492; 25.4834] | * |
|  | Life Quality | ADL | Within × Vulnerability | -0.3689553 | [-0.6928; -0.0451] | * |
|  | Life Quality | ADL | Within × Between | 0.00124874 | [-0.0016; 0.0041] |  |
|  | Life Quality | ADL | Vulnerability × Between | -0.3072309 | [-0.6223; 0.0078] |  |
|  | Life Quality | ADL | Three-way interaction | 0.00879284 | [4e-04; 0.0171] | * |
| Heat Index | Psychological Distress | Economic | Within (main) | 0.19942307 | [-0.2167; 0.6156] |  |
|  | Psychological Distress | Economic | Between (main) | 0.1880089 | [-0.204; 0.58] |  |
|  | Psychological Distress | Economic | Vulnerability (main) | -0.5241863 | [-2.1641; 1.1157] |  |
|  | Psychological Distress | Economic | Within × Vulnerability | 0.02214954 | [-0.0329; 0.0772] |  |
|  | Psychological Distress | Economic | Within × Between | -0.0053186 | [-0.0175; 0.0069] |  |
|  | Psychological Distress | Economic | Vulnerability × Between | 0.00501491 | [-0.0461; 0.0561] |  |
|  | Psychological Distress | Economic | Three-way interaction | -0.0006135 | [-0.0022; 0.001] |  |
|  | Psychological Distress | Education | Within (main) | 0.28072928 | [-0.0297; 0.5911] |  |
|  | Psychological Distress | Education | Between (main) | 0.07814672 | [-0.2252; 0.3815] |  |
|  | Psychological Distress | Education | Vulnerability (main) | -6.3567541 | [-23.8373; 11.1238] |  |
|  | Psychological Distress | Education | Within × Vulnerability | 0.11472747 | [-0.4511; 0.6806] |  |
|  | Psychological Distress | Education | Within × Between | -0.0066527 | [-0.0157; 0.0024] |  |
|  | Psychological Distress | Education | Vulnerability × Between | 0.26341583 | [-0.2729; 0.7998] |  |
|  | Psychological Distress | Education | Three-way interaction | -0.0048551 | [-0.0215; 0.0118] |  |
|  | Psychological Distress | Work | Within (main) | 0.45916519 | [0.228; 0.6903] | *** |
|  | Psychological Distress | Work | Between (main) | 0.32726832 | [0.0868; 0.5678] | ** |
|  | Psychological Distress | Work | Vulnerability (main) | 7.33151453 | [-0.1988; 14.8618] |  |
|  | Psychological Distress | Work | Within × Vulnerability | -0.2366502 | [-0.4822; 0.0089] |  |
|  | Psychological Distress | Work | Within × Between | -0.0126461 | [-0.0195; -0.0058] | *** |
|  | Psychological Distress | Work | Vulnerability × Between | -0.2161412 | [-0.4491; 0.0168] |  |
|  | Psychological Distress | Work | Three-way interaction | 0.00672366 | [-6e-04; 0.014] |  |
|  | Psychological Distress | ADL | Within (main) | 0.37941136 | [0.1652; 0.5937] | *** |
|  | Psychological Distress | ADL | Between (main) | 0.2598891 | [0.0317; 0.4881] | * |
|  | Psychological Distress | ADL | Vulnerability (main) | 0.44366595 | [-15.5358; 16.4231] |  |
|  | Psychological Distress | ADL | Within × Vulnerability | 0.03896405 | [-0.4822; 0.5602] |  |
|  | Psychological Distress | ADL | Within × Between | -0.0105039 | [-0.0168; -0.0042] | ** |
|  | Psychological Distress | ADL | Vulnerability × Between | -0.0139094 | [-0.5056; 0.4777] |  |
|  | Psychological Distress | ADL | Three-way interaction | 0.0005556 | [-0.0149; 0.016] |  |
|  | Mental Health | Economic | Within (main) | -0.3166498 | [-0.5011; -0.1322] | *** |
|  | Mental Health | Economic | Between (main) | -0.3032333 | [-0.4783; -0.1282] | *** |
|  | Mental Health | Economic | Vulnerability (main) | -1.7151956 | [-2.4303; -1.0001] | *** |
|  | Mental Health | Economic | Within × Vulnerability | 0.0664569 | [0.0425; 0.0904] | *** |
|  | Mental Health | Economic | Within × Between | 0.01242261 | [0.007; 0.0178] | *** |
|  | Mental Health | Economic | Vulnerability × Between | 0.05221867 | [0.0299; 0.0745] | *** |
|  | Mental Health | Economic | Three-way interaction | -0.002219 | [-0.0029; -0.0015] | *** |
|  | Mental Health | Education | Within (main) | 0.15325036 | [0.0168; 0.2897] | * |
|  | Mental Health | Education | Between (main) | 0.05720894 | [-0.0773; 0.1917] |  |
|  | Mental Health | Education | Vulnerability (main) | 0.40826714 | [-6.8957; 7.7122] |  |
|  | Mental Health | Education | Within × Vulnerability | 0.02562672 | [-0.2118; 0.2631] |  |
|  | Mental Health | Education | Within × Between | -0.0029066 | [-0.0069; 0.001] |  |
|  | Mental Health | Education | Vulnerability × Between | -0.0456001 | [-0.2705; 0.1793] |  |
|  | Mental Health | Education | Three-way interaction | 0.00048681 | [-0.0065; 0.0075] |  |
|  | Mental Health | Work | Within (main) | 0.21025471 | [0.1042; 0.3163] | *** |
|  | Mental Health | Work | Between (main) | 0.09857365 | [-0.0108; 0.2079] |  |
|  | Mental Health | Work | Vulnerability (main) | 2.64820969 | [-0.5552; 5.8516] |  |
|  | Mental Health | Work | Within × Vulnerability | -0.1025842 | [-0.2071; 0.002] |  |
|  | Mental Health | Work | Within × Between | -0.0043443 | [-0.0074; -0.0013] | ** |
|  | Mental Health | Work | Vulnerability × Between | -0.0614038 | [-0.1606; 0.0378] |  |
|  | Mental Health | Work | Three-way interaction | 0.00233222 | [-8e-04; 0.0054] |  |
|  | Mental Health | ADL | Within (main) | 0.18733155 | [0.0884; 0.2863] | *** |
|  | Mental Health | ADL | Between (main) | 0.09027071 | [-0.014; 0.1945] |  |
|  | Mental Health | ADL | Vulnerability (main) | 6.5505149 | [-0.4344; 13.5354] |  |
|  | Mental Health | ADL | Within × Vulnerability | -0.1828904 | [-0.4106; 0.0448] |  |
|  | Mental Health | ADL | Within × Between | -0.0039332 | [-0.0068; -0.0011] | ** |
|  | Mental Health | ADL | Vulnerability × Between | -0.1890293 | [-0.4049; 0.0268] |  |
|  | Mental Health | ADL | Three-way interaction | 0.00582389 | [-0.001; 0.0126] |  |
|  | Physical Health | Economic | Within (main) | -0.1742379 | [-0.3543; 0.0058] |  |
|  | Physical Health | Economic | Between (main) | -0.1321842 | [-0.2977; 0.0333] |  |
|  | Physical Health | Economic | Vulnerability (main) | -0.8234506 | [-1.5525; -0.0944] | * |
|  | Physical Health | Economic | Within × Vulnerability | 0.03065964 | [0.0062; 0.0551] | * |
|  | Physical Health | Economic | Within × Between | 0.00722165 | [0.0019; 0.0125] | ** |
|  | Physical Health | Economic | Vulnerability × Between | 0.02637499 | [0.0037; 0.0491] | * |
|  | Physical Health | Economic | Three-way interaction | -0.0011956 | [-0.0019; -5e-04] | ** |
|  | Physical Health | Education | Within (main) | 0.08220852 | [-0.0479; 0.2123] |  |
|  | Physical Health | Education | Between (main) | 0.10319797 | [-0.0195; 0.2259] |  |
|  | Physical Health | Education | Vulnerability (main) | -1.5073159 | [-9.1845; 6.1698] |  |
|  | Physical Health | Education | Within × Vulnerability | 0.08106142 | [-0.1674; 0.3295] |  |
|  | Physical Health | Education | Within × Between | -0.0023834 | [-0.0062; 0.0014] |  |
|  | Physical Health | Education | Vulnerability × Between | 0.01178683 | [-0.2238; 0.2474] |  |
|  | Physical Health | Education | Three-way interaction | -0.0012284 | [-0.0085; 0.0061] |  |
|  | Physical Health | Work | Within (main) | 0.09887535 | [0.0061; 0.1917] | * |
|  | Physical Health | Work | Between (main) | 0.08945726 | [-0.0019; 0.1808] |  |
|  | Physical Health | Work | Vulnerability (main) | 1.5599798 | [-1.7744; 4.8944] |  |
|  | Physical Health | Work | Within × Vulnerability | -0.0818637 | [-0.1905; 0.0268] |  |
|  | Physical Health | Work | Within × Between | -0.0024943 | [-0.0052; 2e-04] |  |
|  | Physical Health | Work | Vulnerability × Between | -0.0415375 | [-0.1446; 0.0615] |  |
|  | Physical Health | Work | Three-way interaction | 0.00196535 | [-0.0013; 0.0052] |  |
|  | Physical Health | ADL | Within (main) | 0.10069428 | [0.0167; 0.1847] | * |
|  | Physical Health | ADL | Between (main) | 0.10439657 | [0.0203; 0.1885] | * |
|  | Physical Health | ADL | Vulnerability (main) | 20.6016605 | [13.4798; 27.7235] | *** |
|  | Physical Health | ADL | Within × Vulnerability | -0.6222036 | [-0.8549; -0.3895] | *** |
|  | Physical Health | ADL | Within × Between | -0.0027222 | [-0.0052; -2e-04] | * |
|  | Physical Health | ADL | Vulnerability × Between | -0.6051699 | [-0.825; -0.3853] | *** |
|  | Physical Health | ADL | Three-way interaction | 0.01892394 | [0.012; 0.0258] | *** |
|  | Life Quality | Economic | Within (main) | 0.59167458 | [-0.3373; 1.5207] |  |
|  | Life Quality | Economic | Between (main) | 0.48074693 | [-0.3767; 1.3382] |  |
|  | Life Quality | Economic | Vulnerability (main) | 2.06302597 | [-1.9756; 6.1016] |  |
|  | Life Quality | Economic | Within × Vulnerability | -0.0633068 | [-0.1983; 0.0717] |  |
|  | Life Quality | Economic | Within × Between | -0.0142965 | [-0.0416; 0.013] |  |
|  | Life Quality | Economic | Vulnerability × Between | -0.0628798 | [-0.1873; 0.0615] |  |
|  | Life Quality | Economic | Three-way interaction | 0.00167541 | [-0.0023; 0.0056] |  |
|  | Life Quality | Education | Within (main) | 0.13548563 | [0.0077; 0.2632] | * |
|  | Life Quality | Education | Between (main) | 0.02366772 | [-0.1087; 0.1561] |  |
|  | Life Quality | Education | Vulnerability (main) | 4.28113848 | [-2.4496; 11.0119] |  |
|  | Life Quality | Education | Within × Vulnerability | -0.1619847 | [-0.3811; 0.0571] |  |
|  | Life Quality | Education | Within × Between | -0.0021696 | [-0.0058; 0.0015] |  |
|  | Life Quality | Education | Vulnerability × Between | -0.1150493 | [-0.3218; 0.0917] |  |
|  | Life Quality | Education | Three-way interaction | 0.00466298 | [-0.0018; 0.0111] |  |
|  | Life Quality | Work | Within (main) | 0.15561031 | [0.0563; 0.2549] | ** |
|  | Life Quality | Work | Between (main) | 0.04893215 | [-0.063; 0.1609] |  |
|  | Life Quality | Work | Vulnerability (main) | 1.73494577 | [-1.2103; 4.6802] |  |
|  | Life Quality | Work | Within × Vulnerability | -0.0754021 | [-0.1713; 0.0205] |  |
|  | Life Quality | Work | Within × Between | -0.002757 | [-0.0056; 1e-04] |  |
|  | Life Quality | Work | Vulnerability × Between | -0.0404764 | [-0.1319; 0.051] |  |
|  | Life Quality | Work | Three-way interaction | 0.00182868 | [-0.001; 0.0047] |  |
|  | Life Quality | ADL | Within (main) | 0.13422327 | [0.041; 0.2274] | ** |
|  | Life Quality | ADL | Between (main) | 0.03917793 | [-0.0686; 0.147] |  |
|  | Life Quality | ADL | Vulnerability (main) | 5.06302574 | [-1.2482; 11.3743] |  |
|  | Life Quality | ADL | Within × Vulnerability | -0.1420195 | [-0.3492; 0.0652] |  |
|  | Life Quality | ADL | Within × Between | -0.0022422 | [-0.0049; 4e-04] |  |
|  | Life Quality | ADL | Vulnerability × Between | -0.1243012 | [-0.3182; 0.0696] |  |
|  | Life Quality | ADL | Three-way interaction | 0.00387478 | [-0.0023; 0.01] |  |
| Apparent Temperature | Psychological Distress | Economic | Within (main) | 0.15399263 | [-0.6256; 0.9336] |  |
|  | Psychological Distress | Economic | Between (main) | 0.01918816 | [-0.7282; 0.7666] |  |
|  | Psychological Distress | Economic | Vulnerability (main) | -0.7657316 | [-3.9435; 2.4121] |  |
|  | Psychological Distress | Economic | Within × Vulnerability | 0.03748095 | [-0.0702; 0.1452] |  |
|  | Psychological Distress | Economic | Within × Between | -0.0022823 | [-0.0265; 0.022] |  |
|  | Psychological Distress | Economic | Vulnerability × Between | 0.01294136 | [-0.0882; 0.1141] |  |
|  | Psychological Distress | Economic | Three-way interaction | -0.0011061 | [-0.0045; 0.0022] |  |
|  | Psychological Distress | Education | Within (main) | -0.049879 | [-0.6092; 0.5095] |  |
|  | Psychological Distress | Education | Between (main) | -0.4428498 | [-1.0006; 0.1149] |  |
|  | Psychological Distress | Education | Vulnerability (main) | -15.355693 | [-47.1119; 16.4005] |  |
|  | Psychological Distress | Education | Within × Vulnerability | 0.29440462 | [-0.7702; 1.359] |  |
|  | Psychological Distress | Education | Within × Between | 0.00586262 | [-0.0116; 0.0233] |  |
|  | Psychological Distress | Education | Vulnerability × Between | 0.59097183 | [-0.4169; 1.5988] |  |
|  | Psychological Distress | Education | Three-way interaction | -0.0118904 | [-0.045; 0.0212] |  |
|  | Psychological Distress | Work | Within (main) | 0.55867016 | [0.1751; 0.9423] | ** |
|  | Psychological Distress | Work | Between (main) | 0.26744147 | [-0.1292; 0.6641] |  |
|  | Psychological Distress | Work | Vulnerability (main) | 13.2017954 | [-0.9835; 27.3871] |  |
|  | Psychological Distress | Work | Within × Vulnerability | -0.4065208 | [-0.8797; 0.0666] |  |
|  | Psychological Distress | Work | Within × Between | -0.0145158 | [-0.0265; -0.0025] | * |
|  | Psychological Distress | Work | Vulnerability × Between | -0.4251293 | [-0.8727; 0.0224] |  |
|  | Psychological Distress | Work | Three-way interaction | 0.01281559 | [-0.0019; 0.0275] |  |
|  | Psychological Distress | ADL | Within (main) | 0.47320376 | [0.1226; 0.8238] | ** |
|  | Psychological Distress | ADL | Between (main) | 0.16989096 | [-0.2011; 0.5409] |  |
|  | Psychological Distress | ADL | Vulnerability (main) | 14.5883433 | [-17.5277; 46.7044] |  |
|  | Psychological Distress | ADL | Within × Vulnerability | -0.5207505 | [-1.5888; 0.5473] |  |
|  | Psychological Distress | ADL | Within × Between | -0.0118247 | [-0.0228; -9e-04] | * |
|  | Psychological Distress | ADL | Vulnerability × Between | -0.4160025 | [-1.4222; 0.5902] |  |
|  | Psychological Distress | ADL | Three-way interaction | 0.01672026 | [-0.0162; 0.0497] |  |
|  | Mental Health | Economic | Within (main) | -0.8316341 | [-1.171; -0.4922] | *** |
|  | Mental Health | Economic | Between (main) | -0.8261675 | [-1.1557; -0.4966] | *** |
|  | Mental Health | Economic | Vulnerability (main) | -3.8834453 | [-5.2653; -2.5016] | *** |
|  | Mental Health | Economic | Within × Vulnerability | 0.14703938 | [0.1002; 0.1939] | *** |
|  | Mental Health | Economic | Within × Between | 0.03021655 | [0.0196; 0.0408] | *** |
|  | Mental Health | Economic | Vulnerability × Between | 0.12285085 | [0.0788; 0.1669] | *** |
|  | Mental Health | Economic | Three-way interaction | -0.0048292 | [-0.0063; -0.0034] | *** |
|  | Mental Health | Education | Within (main) | 0.14326109 | [-0.1005; 0.387] |  |
|  | Mental Health | Education | Between (main) | -0.0232931 | [-0.2718; 0.2252] |  |
|  | Mental Health | Education | Vulnerability (main) | 6.6984886 | [-6.8185; 20.2155] |  |
|  | Mental Health | Education | Within × Vulnerability | -0.1900327 | [-0.6443; 0.2643] |  |
|  | Mental Health | Education | Within × Between | -0.00159 | [-0.0092; 0.006] |  |
|  | Mental Health | Education | Vulnerability × Between | -0.2339248 | [-0.6631; 0.1953] |  |
|  | Mental Health | Education | Three-way interaction | 0.0069487 | [-0.0072; 0.0211] |  |
|  | Mental Health | Work | Within (main) | 0.17666707 | [0.0054; 0.3479] | * |
|  | Mental Health | Work | Between (main) | 0.00627262 | [-0.1771; 0.1897] |  |
|  | Mental Health | Work | Vulnerability (main) | 0.18167451 | [-5.8678; 6.2312] |  |
|  | Mental Health | Work | Within × Vulnerability | -0.0119137 | [-0.2139; 0.1901] |  |
|  | Mental Health | Work | Within × Between | -0.0023966 | [-0.0077; 0.003] |  |
|  | Mental Health | Work | Vulnerability × Between | 0.01235819 | [-0.1785; 0.2032] |  |
|  | Mental Health | Work | Three-way interaction | -0.0003809 | [-0.0066; 0.0059] |  |
|  | Mental Health | ADL | Within (main) | 0.21231225 | [0.0544; 0.3702] | ** |
|  | Mental Health | ADL | Between (main) | 0.04660609 | [-0.1266; 0.2198] |  |
|  | Mental Health | ADL | Vulnerability (main) | 21.2197175 | [7.1659; 35.2736] | ** |
|  | Mental Health | ADL | Within × Vulnerability | -0.6787281 | [-1.1454; -0.2121] | ** |
|  | Mental Health | ADL | Within × Between | -0.0037524 | [-0.0087; 0.0012] |  |
|  | Mental Health | ADL | Vulnerability × Between | -0.6377678 | [-1.0783; -0.1972] | ** |
|  | Mental Health | ADL | Three-way interaction | 0.02096681 | [0.0066; 0.0354] | ** |
|  | Physical Health | Economic | Within (main) | -0.3614957 | [-0.7004; -0.0226] | * |
|  | Physical Health | Economic | Between (main) | -0.3581054 | [-0.6814; -0.0348] | * |
|  | Physical Health | Economic | Vulnerability (main) | -1.7885295 | [-3.1973; -0.3797] | * |
|  | Physical Health | Economic | Within × Vulnerability | 0.06759045 | [0.0198; 0.1153] | ** |
|  | Physical Health | Economic | Within × Between | 0.01490633 | [0.0043; 0.0255] | ** |
|  | Physical Health | Economic | Vulnerability × Between | 0.05861144 | [0.0137; 0.1035] | * |
|  | Physical Health | Economic | Three-way interaction | -0.0024188 | [-0.0039; -9e-04] | ** |
|  | Physical Health | Education | Within (main) | 0.11168146 | [-0.1297; 0.3531] |  |
|  | Physical Health | Education | Between (main) | 0.06683587 | [-0.1709; 0.3046] |  |
|  | Physical Health | Education | Vulnerability (main) | -1.5791539 | [-15.6591; 12.5008] |  |
|  | Physical Health | Education | Within × Vulnerability | 0.09769367 | [-0.3745; 0.5699] |  |
|  | Physical Health | Education | Within × Between | -0.0019178 | [-0.0095; 0.0057] |  |
|  | Physical Health | Education | Vulnerability × Between | 0.01195945 | [-0.4348; 0.4588] |  |
|  | Physical Health | Education | Three-way interaction | -0.0016672 | [-0.0163; 0.013] |  |
|  | Physical Health | Work | Within (main) | 0.17455154 | [0.0143; 0.3348] | * |
|  | Physical Health | Work | Between (main) | 0.09470866 | [-0.0677; 0.2571] |  |
|  | Physical Health | Work | Vulnerability (main) | 2.93086092 | [-3.3189; 9.1807] |  |
|  | Physical Health | Work | Within × Vulnerability | -0.1353147 | [-0.3437; 0.0731] |  |
|  | Physical Health | Work | Within × Between | -0.0035583 | [-0.0086; 0.0015] |  |
|  | Physical Health | Work | Vulnerability × Between | -0.0848576 | [-0.2819; 0.1122] |  |
|  | Physical Health | Work | Three-way interaction | 0.00364625 | [-0.0028; 0.0101] |  |
|  | Physical Health | ADL | Within (main) | 0.16572793 | [0.0217; 0.3098] | * |
|  | Physical Health | ADL | Between (main) | 0.1054628 | [-0.0439; 0.2548] |  |
|  | Physical Health | ADL | Vulnerability (main) | 34.05842 | [19.6919; 48.425] | *** |
|  | Physical Health | ADL | Within × Vulnerability | -1.0304969 | [-1.5089; -0.5521] | *** |
|  | Physical Health | ADL | Within × Between | -0.0034964 | [-0.008; 0.001] |  |
|  | Physical Health | ADL | Vulnerability × Between | -1.0396912 | [-1.4896; -0.5897] | *** |
|  | Physical Health | ADL | Three-way interaction | 0.03213905 | [0.0174; 0.0469] | *** |
|  | Life Quality | Economic | Within (main) | 0.32283638 | [0.0133; 0.6324] | * |
|  | Life Quality | Economic | Between (main) | 0.05540624 | [-0.2532; 0.364] |  |
|  | Life Quality | Economic | Vulnerability (main) | 1.24629127 | [-0.0141; 2.5066] |  |
|  | Life Quality | Economic | Within × Vulnerability | -0.0525847 | [-0.0953; -0.0099] | * |
|  | Life Quality | Economic | Within × Between | -0.0042878 | [-0.0139; 0.0054] |  |
|  | Life Quality | Economic | Vulnerability × Between | -0.0334617 | [-0.0736; 0.0067] |  |
|  | Life Quality | Economic | Three-way interaction | 0.00115076 | [-2e-04; 0.0025] |  |
|  | Life Quality | Education | Within (main) | 0.01372342 | [-0.2099; 0.2373] |  |
|  | Life Quality | Education | Between (main) | -0.1468018 | [-0.385; 0.0914] |  |
|  | Life Quality | Education | Vulnerability (main) | 13.0657743 | [0.6634; 25.4681] | * |
|  | Life Quality | Education | Within × Vulnerability | -0.4918986 | [-0.9087; -0.0751] | * |
|  | Life Quality | Education | Within × Between | 0.00224823 | [-0.0047; 0.0092] |  |
|  | Life Quality | Education | Vulnerability × Between | -0.3846245 | [-0.7785; 0.0093] |  |
|  | Life Quality | Education | Three-way interaction | 0.01478835 | [0.0018; 0.0277] | * |
|  | Life Quality | Work | Within (main) | 0.01212097 | [-0.1448; 0.169] |  |
|  | Life Quality | Work | Between (main) | -0.1314376 | [-0.3134; 0.0505] |  |
|  | Life Quality | Work | Vulnerability (main) | -0.1337309 | [-5.7005; 5.4331] |  |
|  | Life Quality | Work | Within × Vulnerability | 0.00236474 | [-0.1835; 0.1883] |  |
|  | Life Quality | Work | Within × Between | 0.00227572 | [-0.0026; 0.0072] |  |
|  | Life Quality | Work | Vulnerability × Between | 0.00823823 | [-0.1674; 0.1839] |  |
|  | Life Quality | Work | Three-way interaction | -0.0002806 | [-0.006; 0.0055] |  |
|  | Life Quality | ADL | Within (main) | 0.0290632 | [-0.1165; 0.1746] |  |
|  | Life Quality | ADL | Between (main) | -0.1161027 | [-0.2902; 0.058] |  |
|  | Life Quality | ADL | Vulnerability (main) | 10.5808585 | [-2.0074; 23.1691] |  |
|  | Life Quality | ADL | Within × Vulnerability | -0.3496743 | [-0.7683; 0.0689] |  |
|  | Life Quality | ADL | Within × Between | 0.0017341 | [-0.0028; 0.0063] |  |
|  | Life Quality | ADL | Vulnerability × Between | -0.2689376 | [-0.6643; 0.1264] |  |
|  | Life Quality | ADL | Three-way interaction | 0.00947704 | [-0.0035; 0.0224] |  |

### S7. Within and between effects of temperature indicators on wellbeing outcomes, 1-year lagged model

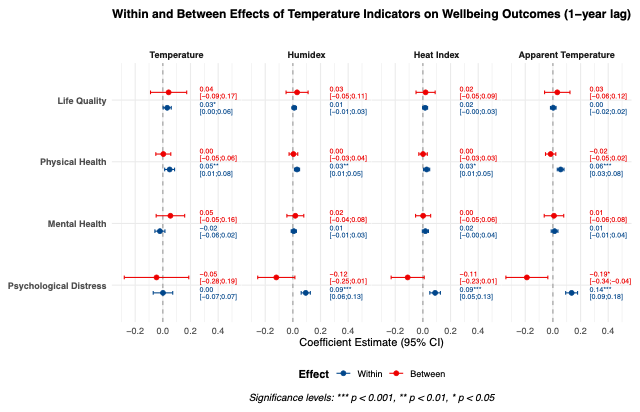

### S8. Within, between and within*between interaction effects, 1-year lagged model

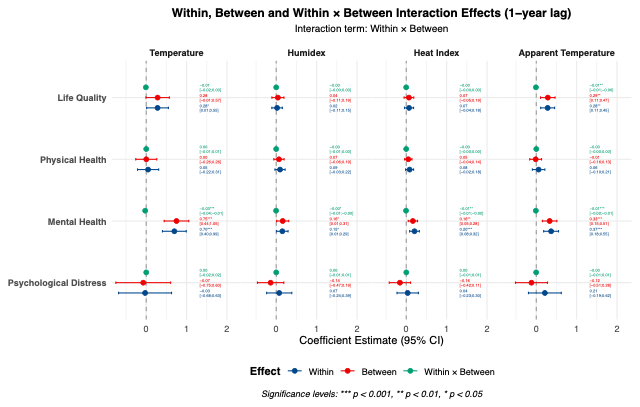

### S9. Within and between effects stratified by vulnerability factors, 1-year lagged model

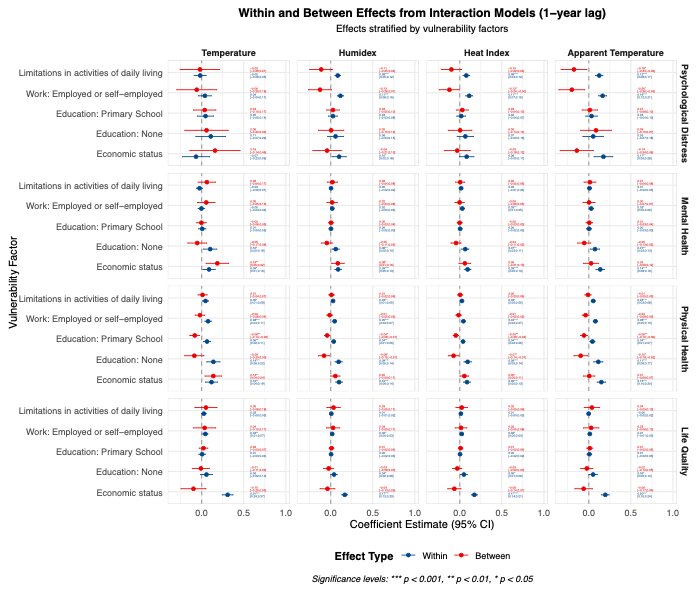

### S10. Negative control (child future prospect) modelling compared to original outcomes (population subset)

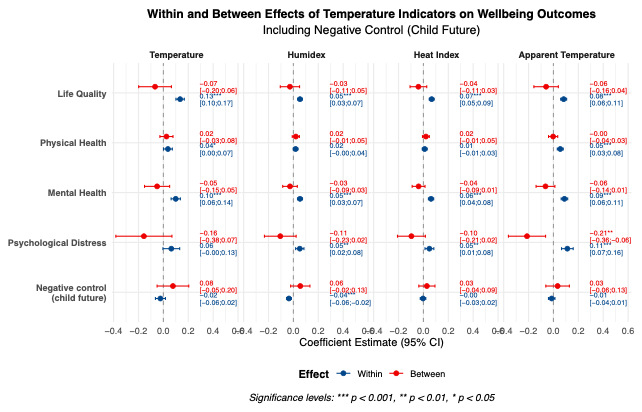
